# Looking beyond the obvious: a critical systematic review and meta-analyses of risk factors for fertility problems in a globalized world

**DOI:** 10.1101/2021.05.06.21256676

**Authors:** R.R. Bayoumi, J. Boivin, H.M. Fatemi, L. Hurt, G.I. Serour, S. van der Poel, C. Venetis

## Abstract

**Background:** Well-established risk factors for fertility problems such as smoking have been included in fertility awareness efforts globally. However, these efforts neglect risks that women in low and middle-income countries (LMIC) face.

**Objective:** To address this gap, we identified eight risk factors affecting women in LMIC and the aim of the current review was to estimate the impact of these risks on fertility.

**Methods:** We conducted systematic reviews and where data was available meta-analyses. We searched Medline, Embase, Cochrane library, regional databases and key organizational websites (1946-June 2016, updated January 2018, latest update taking place in 2021). Two researchers screened and extracted data independently. We included all study designs that assessed exposure to risk in clinical or community-based samples and excluded studies without control groups. The outcome of interest was fertility problems (inability to achieve pregnancy or live birth and neonatal death). We calculated pooled effect estimates from reported effect sizes or raw data. We assessed study quality using the Newcastle-Ottawa Scale. We registered the review with PROSPERO, registration number CRD42016048497.

**Results:** We identified 2,418 studies and included 61 (57 in meta-analyses). Results revealed a nine-fold increased risk of inability to become pregnant in genital tuberculosis (OR 8.91, CI 1.89-42.12) and almost threefold in HIV (OR 2.93, CI 1.95-4.42) and bacterial vaginosis (OR 2.81, CI 1.85-4.27). A twofold increased risk of tubal-factor infertility in Female Genital Mutilation/Cutting–Type II/III (OR 2.06, CI 1.03-4.15) and increased post-natal mortality in consanguinity (stillbirth, OR 1.28, CI 1.04-1.57; neonatal death, OR 1.57, CI 1.22-2.02).

**Strength and limitations:** Reliability of results was bolstered by a rigorous systematic review methodology that is replicable but limited by methodological shortcomings of the available primary studies and the small number of studies in each meta-analysis.

**Conclusions:** The risk factors investigated appeared to impact the reproductive process through multiple biological, behavioural, and clinical pathways. Additionally, infection and pelvic inflammatory disease seemed to be common pathways for several risk factors. The complex multifactorial risk profile can be addressed by LMIC using a global health framework to determine which risk factors are significant to their populations and how to tackle them. The subsequent health promotion encompassing these relevant health indicators could translate into more prevention and effective early detection of fertility problems in LMIC. Finally, the findings of multifactorial risk reinforced the need to put fertility as an agenda in global health initiatives.

## Introduction

### General introduction

“Improved reproductive health and reproductive rights via universal access to sexual and reproductive health care services…” was established as a Millennium Developmental Goal and continues as a target (3.7) within the Sustainable Development Goals^1^. The most recent World Health Organization (WHO) policy paper identifies four areas of reproductive health that “are of equal weight” (i.e., [1] antenatal, intra-partum and postnatal care, [2] contraception counseling and provision, [3] fertility care, [4] safe abortion care)^2^. Fertility care is defined as “interventions that include fertility awareness, support and fertility management with an intention to assist individuals and couples to realize their desires associated with reproduction and/or to build a family”^3^. Within this context, fertility awareness refers to knowledge of reproduction, fecundity, fecundability, related risk factors and reproductive options^3^, and is the least-addressed component of fertility care^4,5^. However, there is growing interest in awareness as an integral aspect of preventative healthcare^6, 7^, and the WHO has made a compelling case for the importance of understanding and addressing exposure to risk as a public health initiative^8^.

As health structures improve, economic and political stability increases and the capacity to address fertility care increases we now need to address the disparity between Low and Middle Income Countries (LMIC) and non-LMIC and within LMIC and we must work together to address the neglected area of fertility care including awareness to enhance the comprehensiveness of reproductive health. Current patterns of fertility in LMIC, declining fertility rates, higher contraceptive use, lower maternal and child mortality, achieved through sustained progress on millennium goals suggest there now is space for a broader reproductive agenda that incorporates fertility care and the complex LMIC risk profile related to communicable and non-communicable diseases, cultural practices and overburdened healthcare systems. The WHO emphasized that health promotion and communicating accurate information about risks has the potential to enhance people’s adoption of healthier behaviours and lifestyle choices^8^.

The impact of reducing burden of disease by targeting distal and proximal risk factors (RFs) through tailored prevention programs applied to communicable and non-communicable disease could potentially be applied to fertility problems. The WHO has made recommendations to assist countries improve health and wellbeing regarding non-communicable diseases, especially in LMIC^9^. These recommendations include the continued development of tools for effective community based education and referral^10^. A global perspective on health implies contextualization^11^. Therefore, integrating in the education and prevention programs health risks arising in different nations due to variation in socio-cultural, environmental, institutional, and economic determinants of health is essential^12^. The recommendations also endorse a global shift highlighting women’s health as an integral aspect of development and achieving millennium goals^13^.

Fertility problems can occur in all countries, but fertility problems present a more complex case in LMIC. Evidence from narrative reviews of risk profiles from the sub-Sahara, the Indian subcontinent and the Middle East suggest that socio-economic and cultural factors in these populations affect the risk profile for female fertility problems^14, 15^. Reproductive health experts concur and suggest that, owing to geographic variation in prevalence and limited quality reproductive health services, women in LMIC or in certain socioeconomic or cultural religious settings could be at greater risk from different factors. This complex multifactorial risk profile for fertility problems in LMIC, in addition to global risks (e.g., smoking) includes exposure to communicable disorders (e.g., tuberculosis, HIV), poorly managed infections owing to constrained healthcare systems (e.g., bacterial vaginosis [BV]) or reproductive events (e.g., birth), consequences of cultural practices (e.g., consanguineous marriages, Female Genital Mutilation/Cutting [FGM/C]) or dubious use of procedures (e.g., Dilatation and curettage [D & C])^16^. One or many of these risks could affect human reproduction with higher co-occurrence in LMIC. It has been suggested that these RFs could influence female fertility directly by compromising the integrity or function of reproductive organs (e.g., genital tuberculosis [GTB], BV) or indirectly through variations in patterns of help-seeking or healthcare provision (e.g., availability of screening programs for early detection of GTB, HIV)^17, 18^.

For this review, RFs affecting female fertility were selected based on a literature search, consultations with experts in the field^16^ and commonly used considerations for the selection of RFs^8, 19^, see Figure 1. RFs affecting male fertility are equally important and need to be examined in future research.

**Figure 1.**
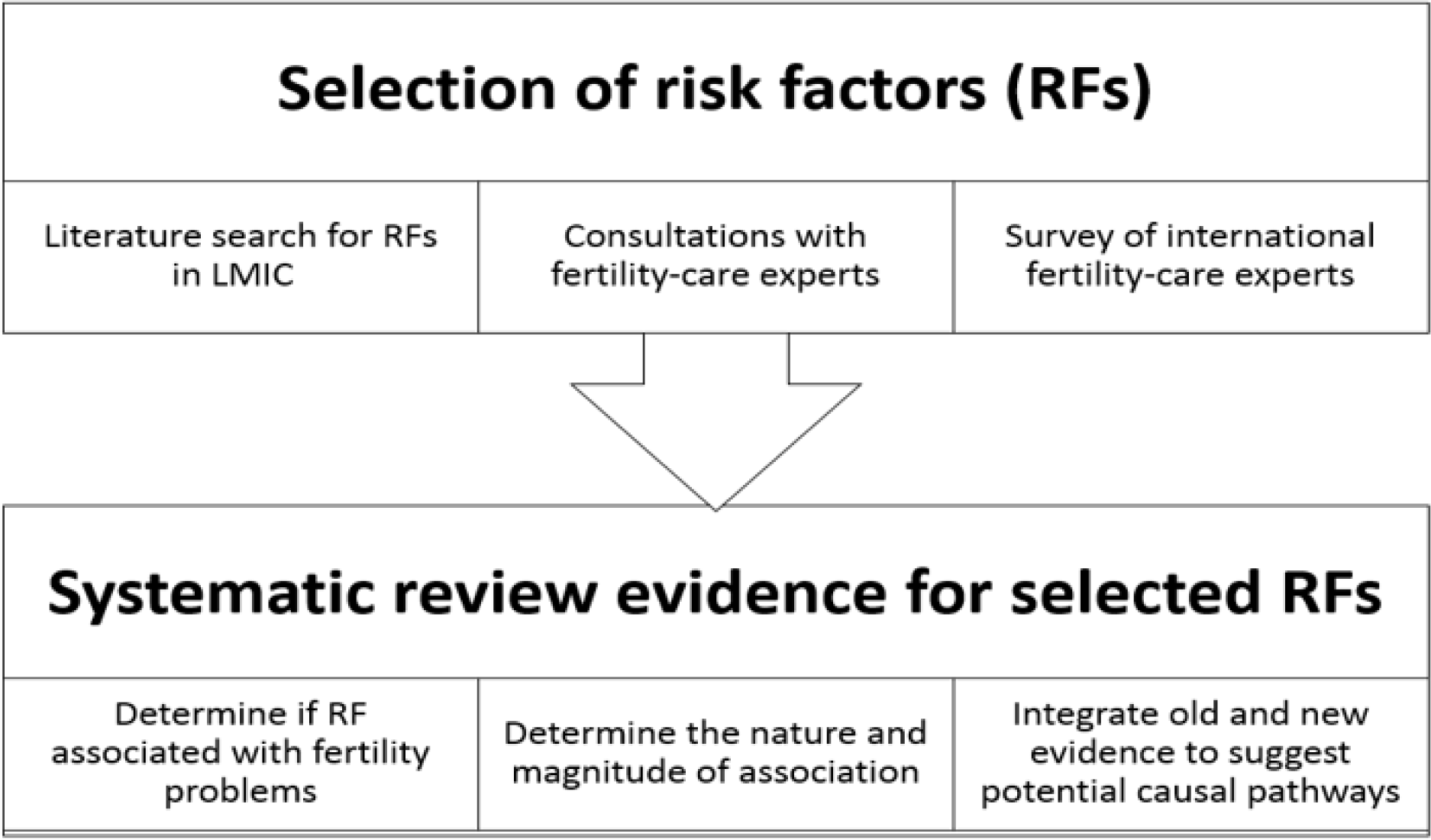
Process of selection of risk factors and systematic review. RF = risk factor, LMIC = low and middle income countries Adapted from Bayoumi et al, 2018

Figure 2 shows the multifactorial risk profile associated with fertility including global (e.g. age, smoking) and non-global factors that are bounded by geography, healthcare resources or culture e.g. HIV, FGM/C. There is systematic review evidence for the effect of many global RFs on fertility (e.g., smoking, alcohol consumption, see Supplemental Table 1). However, a systematic review of the selected RFs (SRFs) long suspected to also be important (especially in LMIC) is now possible and timely due to the emergence of more primary studies. If these SRFs are found to be associated with fertility problems, then they should be considered for inclusion in development or adaptation of educational programs designed to improve informed decision-making about modifiable RFs and timely help-seeking.

**Table 1.**
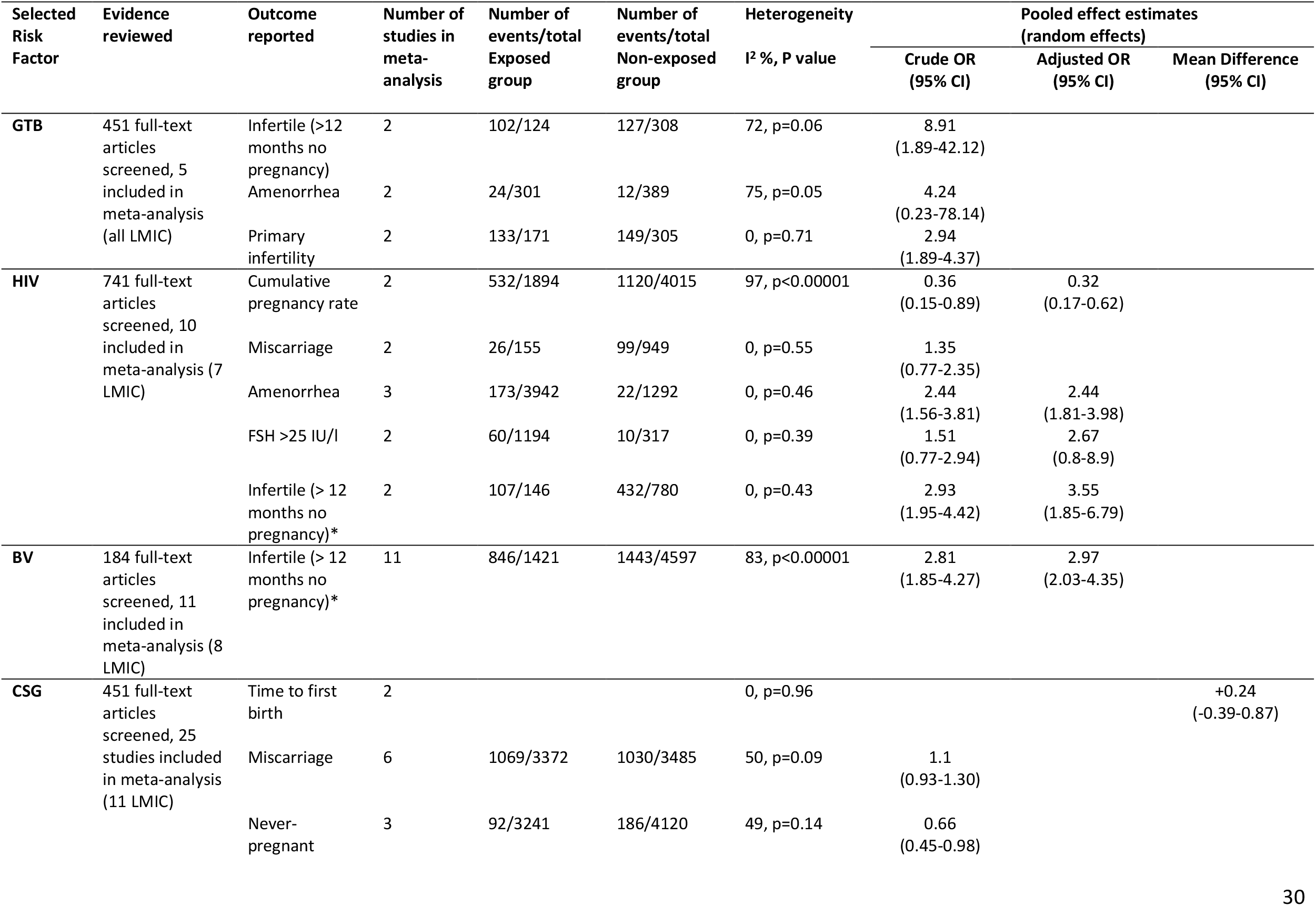
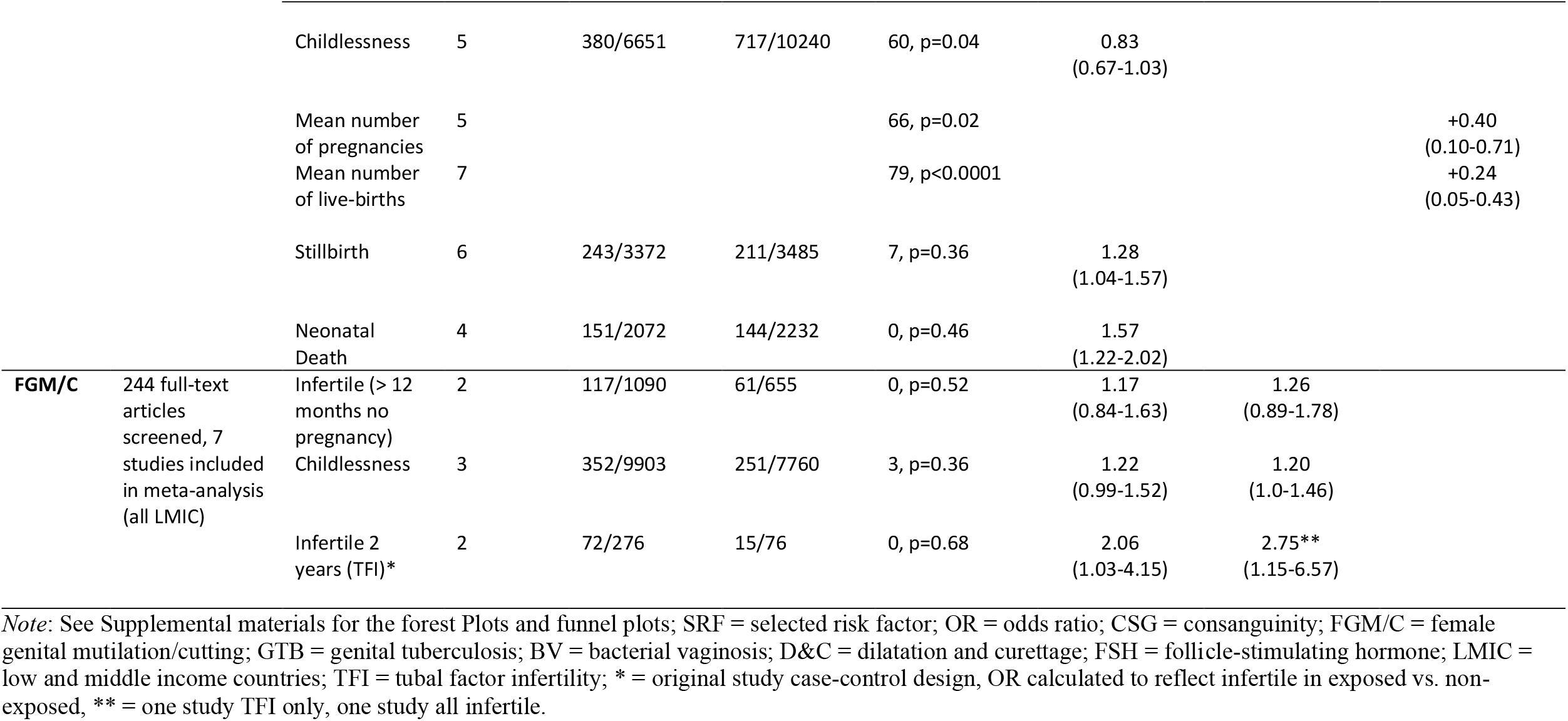
Results of meta-analysis for the five selected risk factors for which it was possible to calculate pooled estimates.

**Figure 2.**
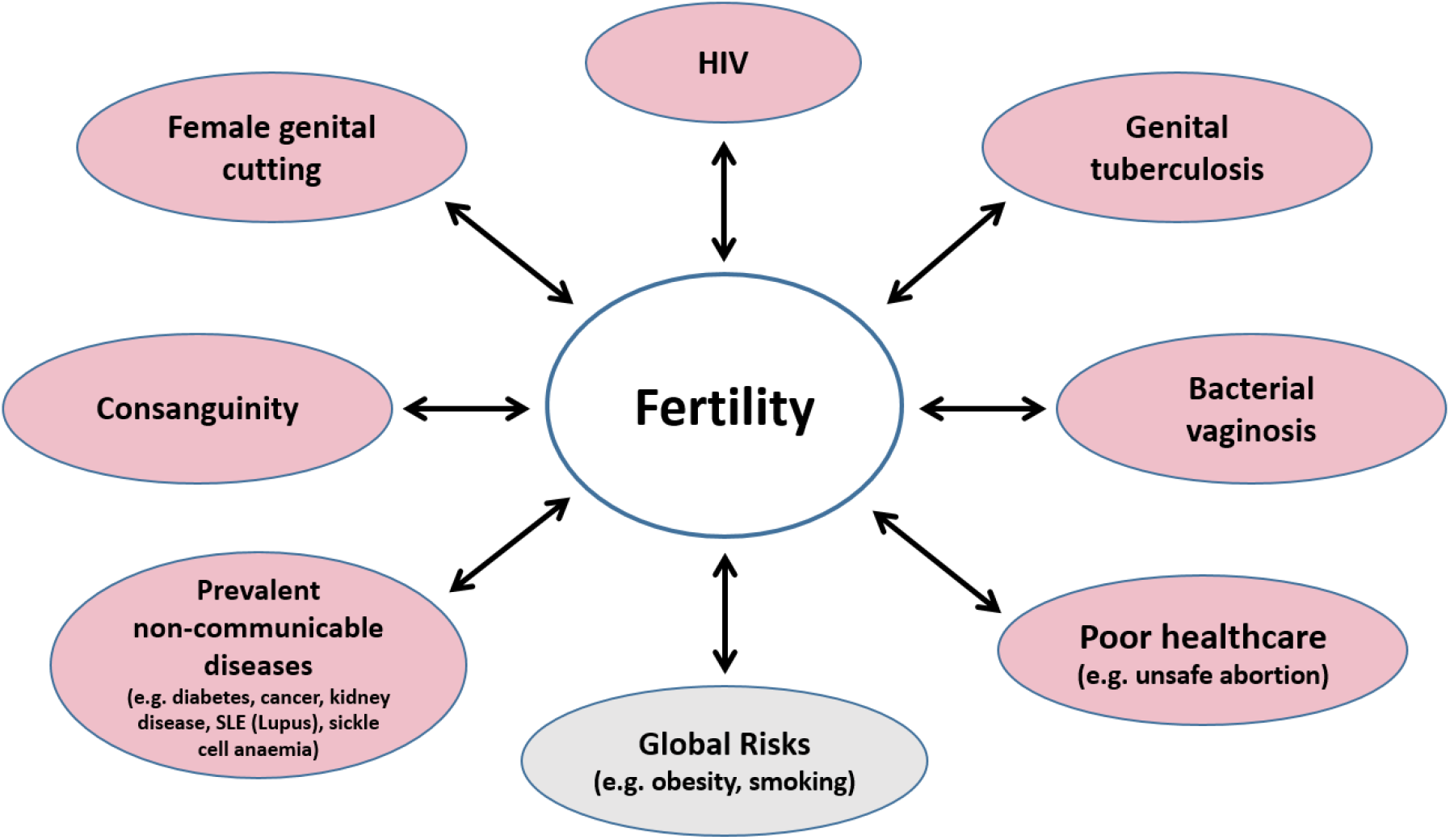
Factors impacting fertility. Some of these factors are included in fertility awareness programs and others were included as a result of the current review.

### The current review

The aim of the current review was to systematically identify and critically appraise the evidence on the impact of the SRFs on female fertility. For each SRF we systematically reviewed the literature and suggested plausible causal mechanisms for effects on fertility based on reproductive outcomes reported in the literature for that SRF. A systematic review of these SRFs will allow an in-depth understanding and translation of the risk profile of communities into fertility education and awareness tools, a necessary step to reduce the burden of fertility problems globally. The eight SRFs identified and included in the current study were: genital tuberculosis (GTB), HIV, bacterial vaginosis (BV), consanguinity (CSG), female genital mutilation/cutting (FGM/C), dilatation and curettage (D&C), vitamin D deficiency and water-pipe smoking.

#### Selected risk factors reviewed

GTB represents 15-20% of extrapulmonary TB and affects about 12% of women who have pulmonary TB^20^. GTB has been shown to cause lesions in the female reproductive tract and it is complications as a result of these lesions that are implicated in fertility problems^21, 22, 23, 24, 25, 26^. At the end of 2016 there were 36.7 million people living with HIV worldwide with 25.6 million people living with HIV in sub-Saharan Africa, which accounts for two thirds of the global total^27^.

HIV has been reported to be associated with reproductive problems, with consistent published evidence for menstrual irregularities, comorbid sexually transmitted infections (STI), tubal blockage, reduced pregnancy and birth rates and increased miscarriage^28, 29, 30, 31^ and less consistent evidence for reduced ovarian functioning and amenorrhea^28, 29, 30, 31^.

BV is an infection characterized by an overgrowth of anaerobic bacteria which causes an imbalance in the naturally occurring vaginal flora^32, 33, 34^. Prevalence of BV in India was reported to be 8.6% (pregnant women)^35^, 29% in America^36^ and 50.9% in Uganda^37^. BV has been reported to be associated with miscarriage, Pelvic Inflammatory Disease (PID), increased susceptibility to viral and other pathogenic bacterial infection, infertility and preterm labour^32, 33, 38, 39, 40^.

A consanguineous marriage is one between close biological relatives^41^. Prevalence is highest in North Africa, West, Central and South Asia (20-50%)^41, 42^. CSG has been suspected to increase pooling of recessive genes that could potentially reduce fertility^43, 44^ or increase gamete compatibility and maternal reproductive span that can enhance fertility^45, 46^.

FGM/C is defined as “all procedures that involve partial or total removal of the external female genitalia or other injury to the female genital organs for non-medical reasons”^47^. Prevalence is highest in North East Africa (Somalia 98%, Egypt 87%, Sudan 88%) and Northern West Africa (Guinea 97%, Mali 89%)^50, 51, 48^. FGM/C has been suspected of being associated with infections, tubal damage and obstetric complications^52, 53, 54, 55^.

D&C is a gynaecological procedure performed to remove tissue from the uterus after birth, miscarriage, and abortion, to treat abnormal uterine bleeding or for diagnosis and treatment of disease^56^. Negative reproductive outcomes such as intrauterine adhesions (IUAs), secondary infertility and recurrent miscarriages have been reported as complications of D&C performed after spontaneous miscarriage^57, 58^. Repeated D&C has been found to be associated with IUAs in meta-analysis^59^, and a single D&C as compared to hysteroscopy was associated with more IUAs^60, 61^. CE is a gynaecological procedure that uses electricity to destroy tissue in the cervix^62^. It is used to treat inflammations, cysts and cancerous or precancerous tissue. Anecdotal reports from Egyptian doctors suggested could cause infertility in LMIC^63^.

Evidence for the association of vitamin D deficiency on fertility comes from molecular level evidence in non-human animal and human studies suggestive of a role of vitamin D in supporting reproductive processes^64, 65^. Vitamin D deficiency is more prevalent in countries where a restrictive dress code is enforced due to social or religious customs (e.g., in the Middle East)^15^.

The methods for using tobacco differ worldwide (e.g., cigarettes, chewing tobacco, and water-pipe use), but according to the WHO, the impact on the human body is similar across methods^66^. The WHO advises that water-pipe smoking is as hazardous to human health as cigarette smoking^67^.

Figure 3 depicts a generic template of how SRFs could potentially impact fertility outcomes. In this figure, the ‘exposure’ column is for the different SRFs, the ‘mechanism’ column is for the potential pathways via which exposure is potentially associated with outcomes and the third column ‘outcome’ is the consequence of the exposure. See supplemental materials for diagrams of proposed pathways for each SRF.

**Figure 3.**
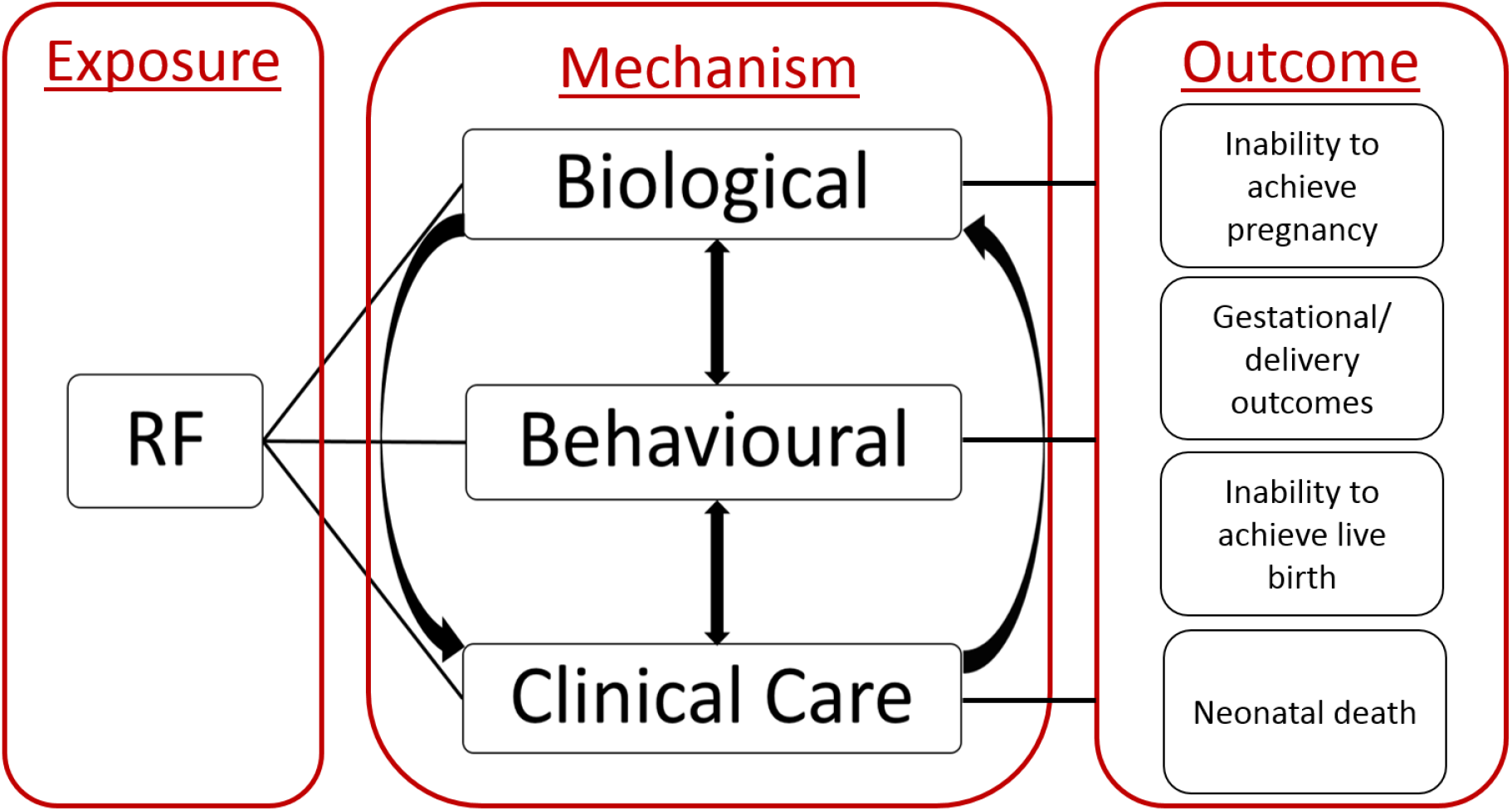
Proposed pathways describing potential impact of selected risk factors on fertility. Figure shows the exposure, the potential mechanisms and the potential outcomes affected. More distal risk factors, such as education or socio-economic status are not shown on the figure, as the overarching aim of this study was to understand the effects of the new risk factors identified for examination in this study. The potential mechanisms shown in the diagrams are informed by an aggregation of the information available in the best quality reviews in the literature. Biological mechanisms refer to changes or effects to physiology or anatomy (e.g., contracting an infection or the formation of scar tissue). Behavioural mechanisms refer to an effect on the actions people take as a result of the exposure (e.g., abstaining from sex after exposure to HIV). Clinical care mechanisms refer to the clinical care required due to the exposure (e.g., obstetric care will change for a woman with Female Genital Mutilation/Cutting [FGM/C]). Outcomes are markers of fertility problems as presented in available studies and can include an inability to achieve pregnancy, gestational or delivery problems and an inability to achieve live birth or neonatal death.

## Methods

We used the Meta-analysis Of Observational Studies in Epidemiology (MOOSE) checklist^68^ in the reporting of the current meta-analyses. The systematic review was registered with PROSPERO, registration number CRD42016048497, link: https://www.crd.york.ac.uk/prospero/display_record.php?RecordID=48497.

### Selection of RFs

RFs were selected based on preliminary literature search, face-to-face consultations and online survey with international infertility experts, as well as regional panel of infertility experts in the Middle East^9^. The following considerations for the selection of RFs that help ensure that risks submitted for deeper study and analysis are likely to be relevant for the disease of interest^2, 12^ were applied: a) determine if the RF is potentially among the primary causes of disease, globally and regionally. If this is unlikely then consider whether the risk can be prevalent and hazardous, or highly concentrated amongst a specific sector; b) assess whether there is a probability of causality based on aggregate of interdisciplinary scientific information; c) determine if data on risk levels and exposure is available or easily extrapolated; d) determine if the risk is potentially modifiable. Such considerations help ensure that risks submitted for deeper study and analysis are likely to be relevant for the disease of interest.

### PICO

For the intent of this review the influence of a risk factor on any dimension of fertility that leads to reduced pregnancy or reduced live births was included under the umbrella term ‘fertility problems’, and outcomes indicative of ‘fertility problems’ operationally defined for this review as inability to achieve pregnancy or live birth and neonatal death (e.g., being childless, episode of infertility). For the purposes of this review the terms ‘impaired fertility’ and ‘fertility problems’ were used synonymously. The Population, Intervention/Indicator, Comparison, Outcome (PICO) question for each SRF was to examine whether the SRF was associated with fertility problems in women, and at what point in the reproductive process the SRF might exert its effects. The population of interest for the reviews was women, the exposure was to the SRF and the outcome of interest was fertility problems. All study designs were included, and the study population consisted of both clinical (clinics, hospitals) and community based samples.

### Search Strategy

Ovid Medline was searched from 1946 to July 2016. All searches were updated in January 2018 to ensure newer studies were included (latest update taking place in 2021). Results reported in each review pertain to original and updated searches (see supplemental figures where the numbers are reported separately in review flow charts).

Fertility problems were searched and combined with ‘OR’ using the following MeSH terms: ‘female fertility’, ‘female infertility’, ‘fertility’ and ‘infertility’. All terms related to the potential SRF (e.g., consanguinity) were searched and combined with ‘OR’. Search terms for the SRF were combined with search terms for fertility problems using ‘AND’. No limits on language or date were used in the search.

The same search strategy was used to search Embase, the Cochrane library and other databases that might be relevant to low resource settings including LILACS, INDMED, Africana Periodical Literature and African Index Medicus. Key organizational websites were searched using the same search terms, including the WHO, United Nations Population Fund (UNFPA), as well as regional sites of these organizations such as the Eastern Mediterranean Regional Office (EMRO) and African Regional Office (AFRO) of the WHO. A search of the reference lists of the included articles was conducted to identify new studies and authors were contacted for missing information. Figure 1 in Supplemental materials shows flowchart of steps taken in the review process.

The exclusion criteria were: (1) use of non-human animal data only, (2) use of male data only, fertility related outcome not reported, (4) association between the SRF and fertility outcome not reported, (5) the SRF reported not of interest, (6) time to birth/duration of childlessness was (on average) less than 21 months because that would imply that pregnancy had occurred within the presumed fertile period of 12 months (i.e., 12 months trying plus 9 months gestation) and (7) study used secondary or qualitative data or was a publication or duplicate record of an included study.

To ensure the comprehensiveness of these search terms to determine if relevant studies that measured the SRF and specific fertility problems without mentioning the word fertility and infertility had been missed, we tested the robustness of our decision by searching for all SRFs combined with MeSH terms for specific indicators of fertility problems that did not include the words fertility and infertility (e.g., tubal occlusion, amenorrhea). These searches indicated that all relevant studies had been captured using the words ‘fertility’ and ‘infertility’, and the additional studies identified did not meet inclusion criteria.

### Data Extraction and Quality Assessment

A standard form was developed and used for extraction of data from included studies and two researchers independently extracted the information. Information was extracted on study design (case-controlled, cross-sectional), sample (location, size), definition of SRF (type of relative, coefficient of inbreeding), the primary outcome fertility problems (as indicated by different outcomes available for each SRF), confounders and data relevant to effect size calculation. Quality assessment of the included studies was based on an adapted version of the Newcastle-Ottawa Scale (NOS)^69^.

### Data Synthesis and Analysis

We used Review Manager (RevMan) [Computer program]. Version 5.3. to calculate effect sizes and meta-analysis and to generate forest plots. The primary outcome measure of association between an exposure and an outcome used was the odds ratio (OR). ORs were calculated from raw data presented in the primary studies as number of events and totals.

In all included analyses an OR of one implied no difference between the exposed (SRF) and non-exposed (no-SRF) groups, an OR greater than one indicated that the exposed group were more likely to have fertility problems (as indicated by the outcomes available in each search) than the non-exposed group, and an OR less than one indicated that the exposed group were less likely to have fertility problems than the non-exposed group.

When means and standard deviations were presented in the primary studies, the primary outcome measure was the mean difference (MD) between exposed and non-exposed groups and original units of measurement were used. Meta-analyses were computed separately for the different outcomes of fertility problems that were reported in the primary studies in each review.

Given that multiple mechanisms may influence how the SRFs affect fertility there may not be one true effect size, therefore a random effects model was deemed appropriate for the data analysis. Heterogeneity was tested using the *Q* statistic and I^2^ index, which specifies the proportion of variance in the effect size not due to chance. Where heterogeneity was statistically significant, subgroup/sensitivity analysis were conducted. The subgroup analyses were based on differences in methodological characteristics of the study e.g., type of control group, subcategories of infertility (tubal factor vs ovulatory).

When data in primary studies were not sufficient to calculate pooled estimates in meta-analyses a systematic review of the evidence was conducted. In such cases the available evidence from the search and from known sources was summarized narratively and conclusions on the potential impact of the SRF on fertility were reported.

### Assessment of Bias

We assessed different types of bias, for example, selection bias, recall bias and bias due to confounder using the modified version of the Newcastle-Ottawa Scale [NOS]^69^.

### Publication Bias

Bias that can affect the generalizability of the results of the review such as publication bias, was also examined. We included a search of grey literature to minimize publication bias. Funnel plots (where there were 10 or more studies), Egger’s test and trim and fill (to impute the number of “missing” studies in the meta-analysis, and to calculate the adjusted pooled effect estimate with the “missing” studies) procedures were used to evaluate publication bias using Comprehensive Meta-Analysis software (Comprehensive Meta-Analysis (Version 2) [Computer software]. A probability level of P<0.05 was used to determine the significance of change in the pooled effect size. Where there were only two studies in an analysis publication bias could not be assessed using funnel plots or trim and fill. These methods do not guarantee the validity of the results of the meta-analysis, but they allow an identification of a potential shortcoming of the review^70, 71^.

## Results

### Search outcome and identified studies

We screened 2,418 articles, and 156 full-text articles were assessed for eligibility. Sixty one primary studies were included in the review, of which 57 were included in meta-analyses and four reviewed systematically, see Figure 4. Data were available for meta-analysis for five of the eight new SRFs. Results for each factor are presented below and are summarized in Table 1. See supplemental materials for the forest plots (where applicable).

**Figure 4.**
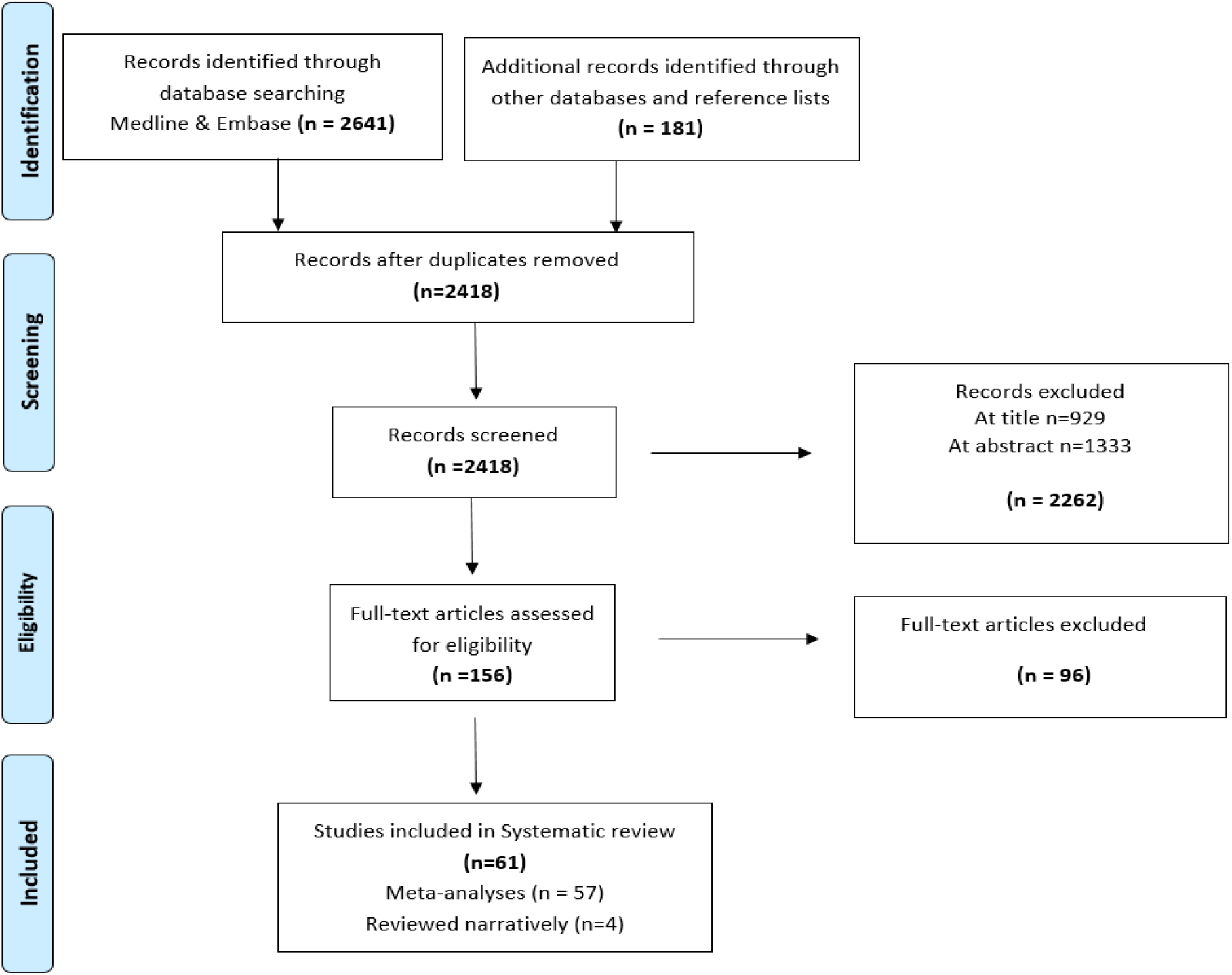
Summary PRISMA Flow Diagram showing the exclusion of articles at the different stages. Of the 57 studies included in meta-analyses, 5 were included in genital tuberculosis, 25 in consanguinity, 7 in Female Genital Mutilation/cutting, 9 in HIV and 11 in bacterial vaginosis analyses. The 4 reviewed narratively were for D&C. Reasons for exclusion at all levels are indicated in PRISMA diagrams for each SRFs in supplemental materials.

### Genital tuberculosis (GTB)

Five cross-sectional studies met inclusion criteria for this SRF. The outcomes they reported were infertility, amenorrhea and primary vs. secondary infertility. Three meta-analyses, each including two studies, were performed, see Table 1. In the first, women with GTB were more likely to be infertile (>12 months) than women without GTB. In the second, women with GTB were equally likely to report amenorrhea as women without GTB. In the third, women with GTB were more likely to have primary infertility than secondary infertility compared to women without GTB.

### HIV

Nine studies met inclusion criteria for this SRF: two case-control, one cohort study and three cross-sectional data embedded within cohort studies. The outcomes reported were cumulative pregnancy rate, amenorrhea, level of FSH greater than 25 IU/l, rate of miscarriage and rate of HIV in infertile and fertile controls. Five meta-analyses were performed, see Table 1. In the first, two studies were included; women with HIV (HIV+) were found to have fewer pregnancies than women without HIV (HIV-). In the second, two studies were included; the HIV+ group were equally likely to report miscarriages as the HIV-group. Three studies were included in the third analysis; the HIV+ group were more likely to have amenorrhea than the HIV-group. Two studies were included in the fourth analysis; the HIV+ group were equally likely to have FSH >25 IU/l as the HIV-group. Two studies were included in the fifth analysis; the HIV+ group were more likely to be infertile than HIV-group.

### Bacterial vaginosis (BV)

Eleven studies were included in this meta-analysis, 10 case-control and one cross-sectional study, see Table 1. The outcomes reported were cases of BV in infertile and fertile women. Women with BV were more likely to be infertile than women without BV. For this SRF, there were enough studies to also conduct pre-specified subgroup analysis, to determine impact on a specific type of infertility, Tubal Factor Infertility (TFI). This analysis showed that the subgroup comprising women with TFI only and the subgroup comprising women with multiple types of infertility were both significant and the difference between subgroups was significant.

### Consanguinity (CSG)

Twenty-six studies, 20 cross-sectional and four cohort studies were included in eight meta-analyses, see Table 1. The outcomes examined were time to first birth, never having been pregnant, childlessness, mean number of pregnancies and live-births, number of miscarriages, stillbirths and neonatal deaths. Eight meta-analyses were performed comparing CSG couples and those who were unrelated, see Table 1. There was no difference in average time to first birth and miscarriage. CSG couples were less likely to have never been pregnant but there was no association with childlessness. CSG couples had significantly more pregnancies and live births than unrelated couples. More still births and neonatal deaths were found in the CSG couples.

### Female genital mutilation/cutting (FGM/C)

Seven studies were included in the meta-analysis, five cross-sectional and two case-control studies. The outcomes they reported were infertility, childlessness and a comparison of cases with tubal infertility and pregnant controls. Three meta-analyses were performed, see Table 1. The first included two studies showing that women who had undergone FGM/C were not more likely to be infertile (>12 months) than women without FGM/C. Three studies were included in the second analysis and the odds of being childless were marginally higher in women with FGM/C than women without FGM/C (and significantly higher using adjusted ORs). The third analysis included two studies showing women with FGM/C Type II and III (severe types) were more likely to be diagnosed with tubal factor infertility (TFI) than women who had undergone Type I.

### Dilatation and curettage (D&C)

Four studies met the inclusion criteria, three cohort and one cross-sectional study. Pooled estimates could not be calculated because the studies all used different outcomes; therefore, results are summarized narratively. In a cohort study, women who had undergone D&C to remove retained products of conception (RPOC) experienced longer time to pregnancy and more ‘new infertility’ diagnoses compared to women who had undergone hysteroscopy^72^. In a cross-sectional study, women who had a history of D&C as part of infertility investigation had significantly more PID than women who had no such history^73^. In a cohort study, women who had undergone D&C developed more gynaecological disease (e.g., endometriosis, irregular uterine bleeding) and menstrual irregularity than women who had vacuum aspiration and/or received prostaglandins^74^. In a cohort study, there were no differences in the number of future pregnancies, normal deliveries, miscarriages, and infertility in women who had undergone D&C after miscarriage compared to women who had experienced expectant management^75^.

### Vitamin D deficiency and water-pipe smoking

A recent high-quality systematic review^76^ summarized the literature on the association between vitamin D deficiency and fertility, making an update unnecessary. This review^76^ indicated first that there was molecular and epidemiological evidence suggesting that vitamin D was involved in the physiologic processes of markers for ovarian reserve (e.g., anti-Mullerian hormone [AMH]), second, evidence from molecular, epidemiological and meta-analyses for a relationship between vitamin D deficiency and PCOS was not consistent and third, molecular evidence suggests that vitamin D could modulate inflammation and proliferation in endometriosis, but epidemiological evidence has been inconsistent^76^. The authors identified methodological shortcomings in the primary studies that may affect the interpretation of the results, such as small samples. We suggest that the inconsistency could also be due to the phenotypical expression of such a relationship (physiologic processes) that may be more complex and therefore difficult to measure, and there could be confounding effects (e.g., better nutrition and health overall) not consistently measured or reported.

A systematic search for water-pipe smoking was not necessary since water-pipe is only a different method to consuming tobacco and the WHO has recently conducted a review which concluded that the use of water-pipe smoking is as hazardous to human health as cigarette smoking. Specifically, a one hour water-pipe session is thought to be equivalent to inhaling 100-200 times the volume of smoke in a single cigarette^67^ and the impact of smoking cigarettes on fertility is well established^77^.

### Multifactorial Risk Model

Results of all meta-analyses were aggregated with extant evidence and used to construct a model that depicts how reviewed SRFs impact fertility using outcomes reported in the primary studies, see Figure 5.

**Figure 5.**
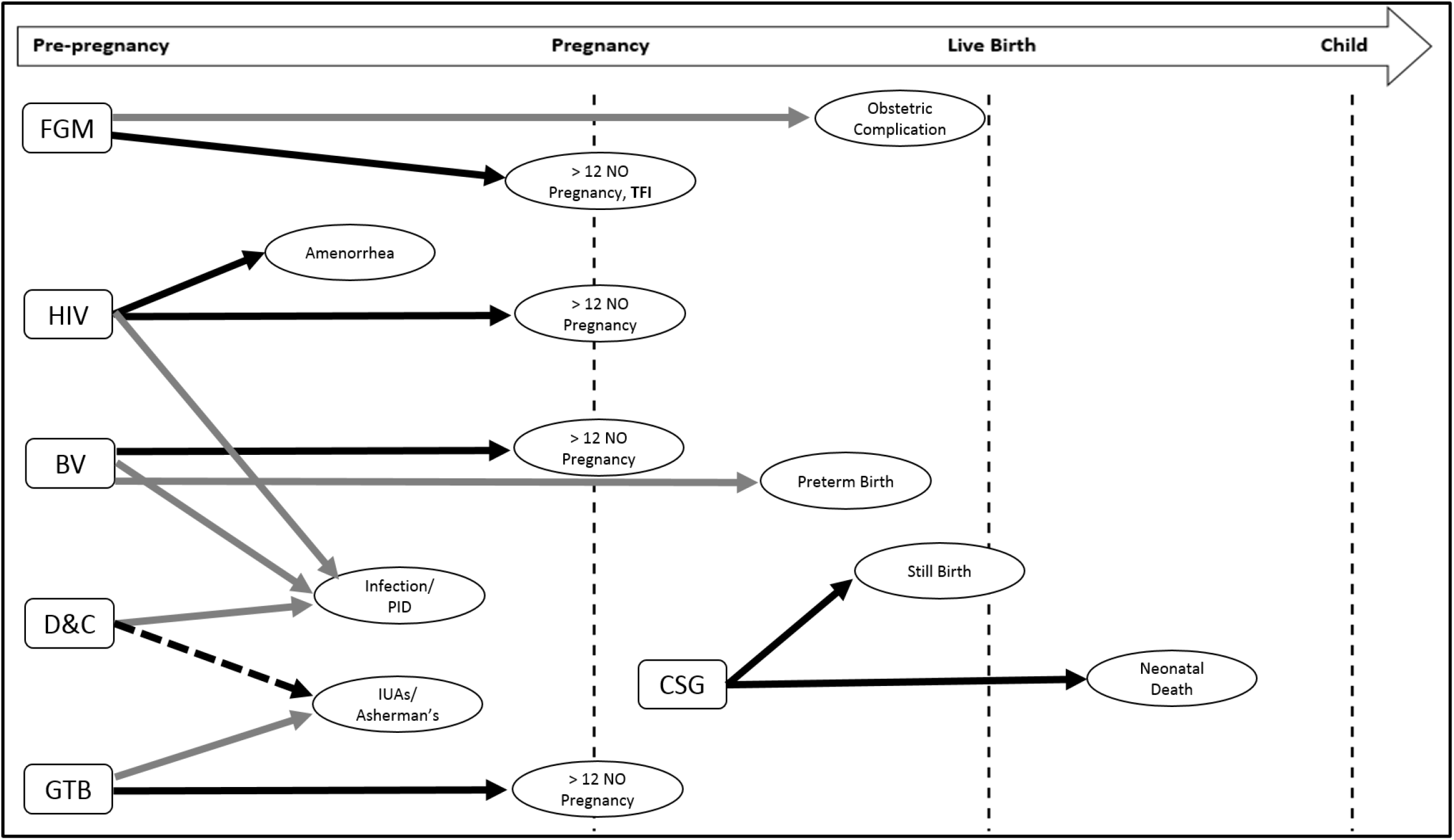
Association between risk factor and fertility problems according to type of evidence and proposed timing of effect in reproductive process. Type of evidence: evidence from current meta-analysis (solid black arrow), evidence from previous meta-analysis (dashed arrow), evidence from primary studies or narrative reviews (grey arrow); CSG = consanguinity; FGM/C = female genital mutilation/cutting; GTB = genital tuberculosis; BV = bacterial vaginosis; D&C = dilatation and curettage; PID = pelvic inflammatory disease; LMIC = low and middle income countries; TFI = tubal factor infertility

## Discussion

### Principal findings

The SRFs investigated were associated with fertility through multiple biological, behavioural and clinical pathways and meta-analytic results were consistent for the most part with past narrative reviews. GTB was shown to have a nine fold increased risk of inability to become pregnant, HIV and BV found to have an almost threefold increased risk of inability to become pregnant within 12 months. While other risks that are highly prevalent in some regions, such as FGM/C (Type II and III) a twofold increased risk of TFI (∼90% in some African nations)^51^ and CSG, detrimental effects such as post-natal mortality (50% of marriages in some nations)^42^.

#### Multiple global risks

A focus on prevalent risks in higher income countries or single risks could obscure the multifactorial risks to which people in LMIC could be exposed, see Figure 6. What can and should be done about risk exposure needs to be determined within countries and regions utilizing a global health framework. The findings of multifactorial risk also reinforced the need to put fertility as an agenda in global health initiatives.

**Figure 6.**
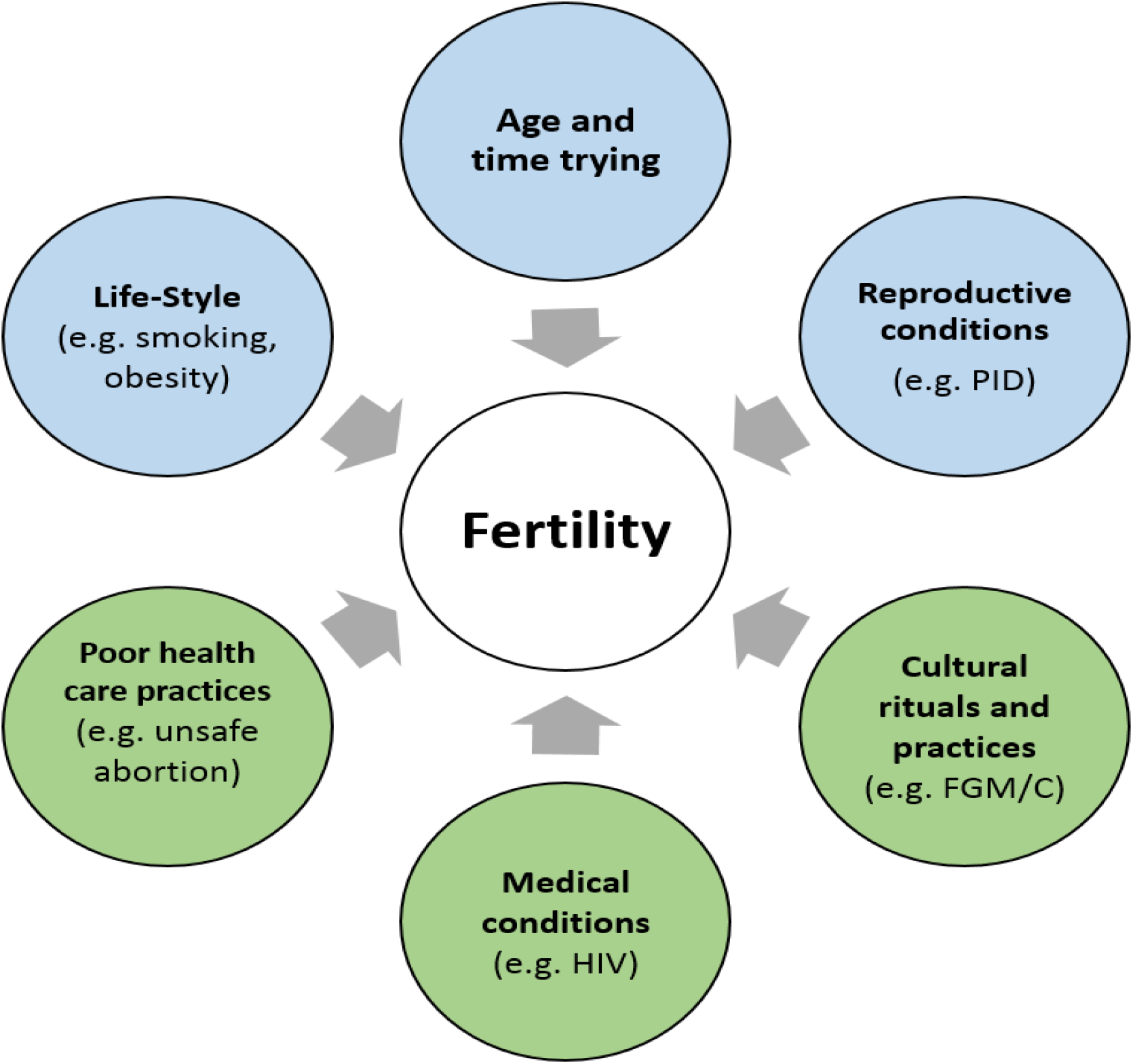
Factors impacting fertility. Some of these factors are included in fertility awareness programs and others were included as a result of the current review. Time trying refers to the time trying to achieve pregnancy. Reproductive and gynaecological refers to conditions or procedures affecting the reproductive tract. Medical conditions refer to both communicable and non-communicable diseases. Blue circles indicate risks that are relevant globally and green circles indicate risks that may not be relevant globally. PID = Pelvic Inflammatory Disease; FGM/C = Female Genital Mutilation/Cutting.

#### Where in the reproductive tract the impact appears to be

Some SRFs such as BV and FGM/C seem to have an impact at several stages in the reproductive process. In the case of BV this could be due to the fact that infection that occurs before pregnancy and reaches the tubes will compromise ability to achieve pregnancy, while infection that occurs during pregnancy could damage the amniotic sac and lead to preterm birth. In the case of FGM/C, it is likely that the TFI occurs secondary to infection arising from the more severe types of cutting where the anatomy is altered drastically. It should be noted that even if the cutting did not lead to infection, a woman could still be at risk of obstetric complications if the altered anatomy made delivery difficult as noted in the literature^52, 54, 55, 78, 79^. Therefore, it can be inferred that timing and extent of exposure to SRFs could affect fertility in different ways, consequently prevention and management should be informed by these mechanisms.

#### Common pathways (infection)

Some SRFs have common pathways, for example HIV, BV and D&C were all related to infection and PID. Although with FGM/C there was no data suggesting a direct link with infections and PID, it can be assumed that would be the case because of the association with TFI. All of these risk factors could affect fertility due to the progression of infection, namely that any infection to the reproductive tract if left untreated could lead to PID, ascend to the tubes, or lead to tubal damage and therefore inability to achieve pregnancy^81, 82^. However, there were no consistent findings to suggest that infections always led to inability to achieve pregnancy. This is probably because the impact would only appear if the infection remained untreated. Infections treated before they lead to PID would have no impact on the female reproductive tract and hence future ability to achieve pregnancy^80^. Furthermore, not all infections lead to PID and not all cases of PID lead to tubal damage^81^. Future research should ensure that data about treatment of infection is collected.

What can be clearly gleaned from Figure 5 is that the SRFs that included infection, PID or TFI in their pathways (e.g. BV, HIV and FGM/C) were found to be associated with an inability to achieve pregnancy, affirming historic reports in the literature about the association between infection and infertility in Africa and other LMIC^14, 82, 83, 84^. The available evidence would suggest that whilst infection is a shared pathway its potential causes are multiple and clinicians need to be mindful of all of the risks for infection and not just STIs and unsafe procedures (abortion, delivery) as has typically been the case^83, 85^.

### Strengths and limitations

The review process used rigorous systematic review methodology that is replicable. Two independent researchers duplicated screening/data extraction and used best-practice guidelines in the design, assessment and reporting of methodology which helped bolster the trustworthiness of the results.

The small number of studies in each meta-analysis limited the generalizability of results and potentially increased publication bias. However, assessment of publication bias for all meta-analyses using visual assessment of funnel plot asymmetry, trim and fill procedures and Egger’s tests did not alter the results, see supplemental materials.

Regardless of how rigorous the review process was, results could only be as strong as the primary studies included. Three limitations of the primary studies were similar across SRFs. First, in a majority of studies, recruitment occurred at fertility clinics, possibly limiting selection to women at higher risk of infertility (applicable to GTB, FGM/C, BV). Second, the definition of outcomes, period of exposure or type of infertility were often not reported (applicable to CSG, BV, HIV). Third, was the lack of inclusion of confounders potentially moderating the effect of the risk. For example, the type of circumciser in FGM/C could be linked to an increase in the likelihood of infection and comorbid STIs (applicable to HIV and BV). Only 10 of the 57 primary studies reported adjusted ORs. However, the use of adjusted ORs did not alter the results of the reviews except in one instance, where women with FGM/C reported more childlessness than women without FGM/C. It could be that had more primary studies reported adjusted ORs the results of the review may have changed but given that significance changed in only one of eight meta-analyses we can be reassured that the results might not have been impacted greatly by the lack of reporting of adjusted ORs. It is important to note that in six of the eight adjusted OR meta-analyses the magnitude of the effect size increased indicating that the association with fertility problems was related to the SRFs and not the confounders adjusted for. Additionally, heterogeneity remained the same or decreased in seven of the eight adjusted meta-analyses, indicating that the confounders were indeed increasing the heterogeneity.

### Implications of Findings

Targeting communicable and non-communicable diseases is not only a priority to reduce the effects of these conditions on health in general but also their impact on childbearing should be addressed. The findings strongly support the movement toward having a more global understanding of risk for disease, and by extension different settings can determine themselves which risk factors are key for their health providers and populations. This would allow health promotion to encompass culturally relevant health education and promotion. This understanding could ultimately translate into more effective early detection of fertility problems in LMIC.

Clinical implications of these findings include education about the impact of these SRFs that should be disseminated widely and in the most culturally appropriate manner. These results should be disseminated to clinicians who can have discussions with individuals about these SRFs that can lead to better choices to protect reproductive capacity and to ensure that there is informed decision-making about factors that impact fertility. The findings have specific implications for clinicians and women, and wider implications for the integration of fertility within the global reproductive health agenda. Awareness of the risks associated with reviewed SRFs should be communicated to couples, especially where the threat of the SRF is increased (e.g. high prevalence such as FGM/C in some countries, family member with TB). Appropriate education, awareness, support, and training initiatives are urgently needed to empower people to maintain or improve their fertility, quality of life, and productivity.

### Unanswered Questions and Future Research

Future research needs to determine what is the best method of selecting RFs, methods to systematically evaluate pathways leading to fertility problems, particularly more rigorous prospective designs or RCTs aimed at modifying risks (where possible). The methodological rigor of the systematic review process adopted enhanced reliability, however, small number of primary studies and inconsistencies in outcome measures were limitations.

The specific examples of what research needs to be conducted for each SRF were informed from the gaps in primary studies included using more rigorous methodology like RCTs where that is ethical and possible or longitudinal cohort studies. It also encompassed the inclusion of well-defined and consistent outcomes and the inclusion of confounders. Future research should also target an understanding of the causal pathways, for example more molecular level investigations. The implications of uncovering the exact causal pathways would be that more specific clinical recommendations and best practice guidelines could be established. Furthermore, research endeavours can be enhanced with the adoption of a more systematic approach to studying fertility globally.

Most importantly, the results highlighted the necessity of multinational cooperation between research teams to fill the gaps identified. To understand and address these gaps in LMIC requires a multidisciplinary approach involving public health, reproductive medicine, the emerging field of global health psychology and other relevant fields.

## Data Availability

The data that support the findings of this study are available in the supplementary file. Additional data are available on request from the corresponding author.

## Data availability

The study was registered with the PROSPERO registry, PROSPERO registration number CRD42016048497, https://www.crd.york.ac.uk/prospero/display_record.php?RecordID=48497 Detailed results available in the supplemental materials and additional data are available upon request from the corresponding author.

## Conflict of interest

The authors declare no conflicts of interest.

## Funding

This research was supported by the Economic and Social Research Council, Global Challenges Research Fund (Impact Acceleration Grant No. ES/M500422/1) and the World Health Organization/Human Reproduction Research Programme (WHO/HRP; UNDP/UNFPA/UNICEF/WHO/World Bank Special Programme of Research, Development and Research Training in Human Reproduction).

## Notes

### Competing Interest Statement

The authors have declared no competing interest.

### Author Declarations

School of Psychology Research Ethics Committee Cardiff University Tower Building 70 Park Place Cardiff CF10 3AT

