## Supplemental Materials for "Looking beyond the obvious: a critical systematic review and meta-analyses of risk factors for fertility problems in a globalized world"

### Operational definitions and Abbreviations:

**Amenorrhea**: an absence of menstruation
**Infertility**: A disease characterized by the failure to establish a clinical pregnancy after 12 months of regular, unprotected sexual intercourse or due to an impairment of a person's capacity to reproduce either as an individual or with his/her partner. Fertility interventions may be initiated in less than 1 year based on medical, sexual and reproductive history, age, physical findings and diagnostic testing. Infertility is a disease, which generates disability as an impairment of function.
**Fertility problems**: operationally defined for this review as inability to achieve pregnancy or live birth and neonatal death.
**Primary female infertility**: A woman who has never been diagnosed with a clinical pregnancy and meets the criteria of being classified as having infertility.
**Secondary female infertility**: A woman unable to establish a clinical pregnancy but who has previously been diagnosed with a clinical pregnancy.

BV: Bacterial vaginosis
CSG: Consanguinity
D&C: Dilatation and curettage
FGM/C: Female genital mutilation/cutting
GTB: Genital tuberculosis
LMIC: Low and middle income countries
PICO: Population, intervention/Indicator, comparison, outcome
PID: Pelvic inflammatory disease
RF: Risk factor
SRF: Selected risk factor
STI: Sexually transmitted infection
WHO: World Health Organization

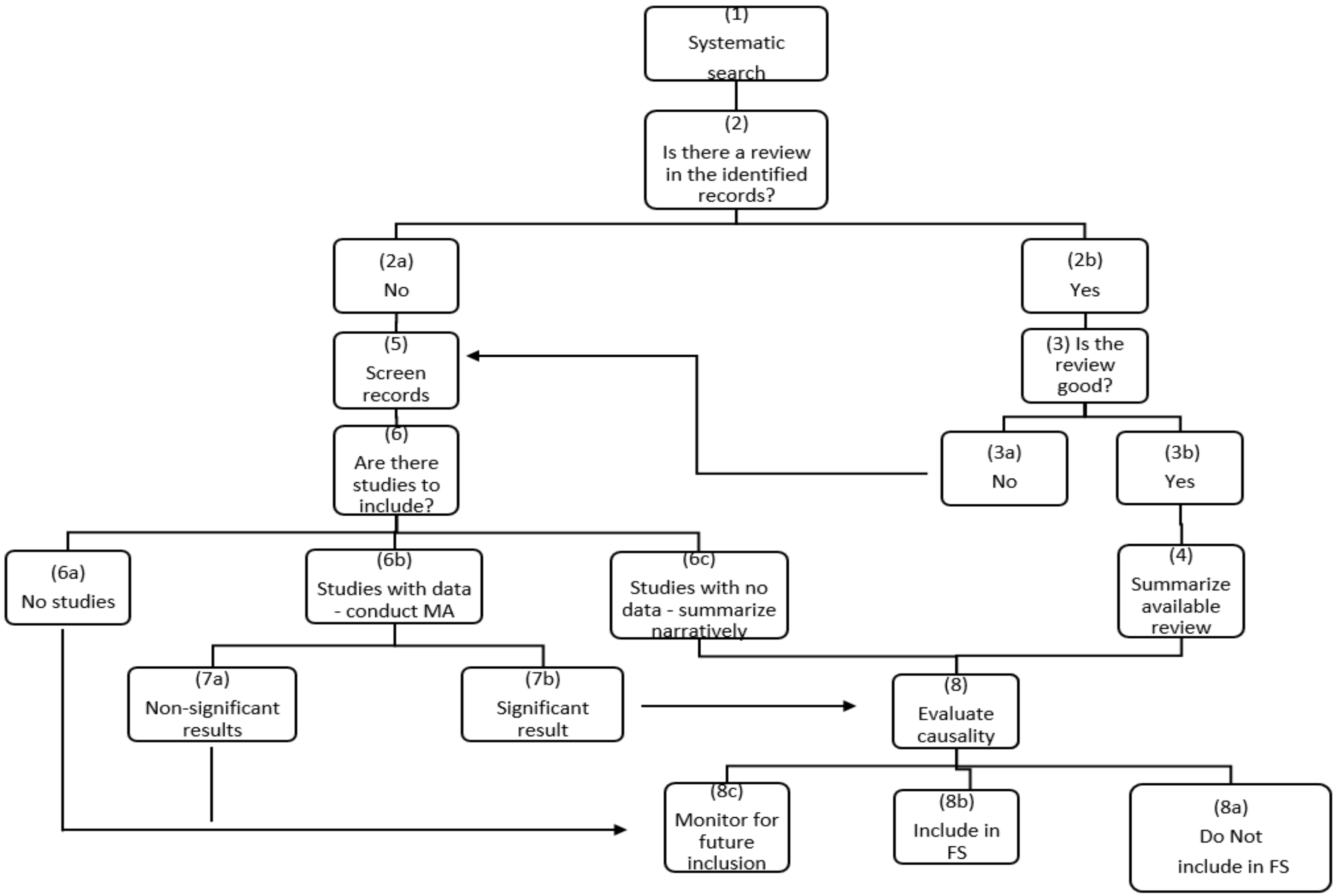


**Key:**
(1) Search Embase and Medline using Mesh terms (see pp XX for details about exact methodology)
(2) Is there a review found?
(a) No Review found → 5
(b) Review found → 3
(3) Evaluate the review using the "Critical Appraisal of Systematic Reviews" published by the WHO (Abalos, Carroli, Mackey & Bergel, 2001), and make sure is current (less than five years)
(a) Not Good → 5
(b) Good → 4
(4) Summarize results in the available review → 8
(5) Screen records as per inclusion and exclusion criteria (pp XX)
(6) Are there studies to include in the SR and are there data?
(a) No studies to include in SR → report on this
(b) Studies found and data available: conduct meta-analysis → 7
(c) Studies found but no data to be included in a meta-analysis: report results narratively → 8
(7) Results of MA
(a) Results not significant → report on this
(b) Results are significant → 8
(8) Evaluate causality and evidence using Bradford-hill criteria and potential pathways diagram, and make a decision about inclusion in FertiSTAT based on all info from results and literature search about potential pathways and quality of evidence available for all arms of the diagram
(a) Include in FertiSTAT (FS) - which version
(b) Do not include in FS - other recommendations
(c) Monitor for future inclusion in FS

*Figure 1* Flowchart showing the decision-making process used in the current systematic reviews. MA = meta-analysis; FS = FertiSTAT

Table 1.

Summary of Findings from Reviews on the Impact of Original FertiSTAT Risk Factors on Fertility

| Risk factor | Summary findings | Type of review | Source |
|---|---|---|---|
| **Age, lifestyle and reproductive** | | | |
| **Age** | Increasing parental age is a risk factor for reduced fertility. | Narrative Review | Schmidt, 2012 |
| **Age** | Birth rate starts to decrease when a woman reaches 35 years old. Young women conceive sooner than older women. Infertility increases as the age of the female increase. | Narrative Review | Liu, 2011 |
| **Appendectomy** | No statistical association between appendectomy and infertility | Systemic Review and meta-analysis of RCTs | Elraiyah, 2014 |
| **Pelvic surgery** | Adhesions are a common complication of gynaecological surgeries. Adhesions affect the interaction between the fallopian tube and ovaries consequently infertility can occur. | Narrative review | Hirschelmann, 2012 |
| **Chlamydia** | Inflammatory tissue destruction in response to infection leads to the development of tubal infertility and ectopic pregnancy | Narrative Review | Carey 2010 |
| **Endometriosis** | Dysfunction of pituitary-ovarian axis altering the feedback pathways, folliculogenesis, lower levels of estrogen and progestron, altered luteal function and the fact that they ovulate fewer oocytes are all accounted for infertility in women with endometritis | Narrative review | Stilley, 2012 |
| **Lifestyle** | Fertility is decreases by being overweight and underweight.<br>Folic acid and Vitamin B have been linked to infertility and spontaneous abortions.<br>High alcohol consumption can affect estrogen and progestron levels leading to anovulation, luteal phase dysfunction and impaired implantation. | Narrative Review (in some cases review of reviews e.g. in smoking several systematic reviews and meta- | Anderson, 2010 |

| | | | |
|---|---|---|---|
| | Consumption of caffeine in moderation has no effect on fertility however some evidence suggest that prolongs time to conception.<br>Smoking adversely effects fertility and pregnancy outcomes.<br>Recreational drugs are associated with decrease fertility, some prescription medications such as anti-hypertensives can affect the female reproduction on different levels.<br>Stress can supress the reproductive functions such as causing hypothalamic amenorrhea.<br>Environmental pollutant can cause a negative effect on fertility.<br>Evidence of oxidative stress has been found in women with PCOS, unexplained infertility and endometriosis. | analyses are reviewed here) | |
| **Lifestyle** | Increasing age of a women increases infertility and time to pregnancy.<br>Consuming more vitamins & proteins and less carbs & trans fats are recommended to preserve fertility.<br>Body weight has significant effect on infertility. Obesity increases the risk of miscarriages however being underweight is associated with ovarian dysfunction and infertility.<br>Vigorous exercise was found to have a negative effect on female reproduction by causing hypothalamic dysfunction and therefore menstrual abnormalities.<br>Physical stress can prolong the time to conceive, however psychological stress is more prominent among women attending the infertility treatment.<br>Smoking decrease the ovarian function and ovarian reserve.<br>Marijuana use increases the risk of primary infertility. Prescription medications such as anti-psychotics, anti-hypertensives and chemotherapy.<br>The amount of alcohol and caffeine consumed significantly affects the fertility of women.<br>Exposure to heavy metals such as lead is reported to alter hypothalamic-pituitary axis and overall fertility. | Narrative review | Sharma 2013 |

| | | | |
|---|---|---|---|
| **Obesity** | Obesity increases the risk of anovulatory infertility because of hyperandrogensim through granulosa cell apoptosis, peripheral conversion of androgens to estrogen leading to an increase negative feedback of gonadotropins and adverse effect on theca and granulosa because of increased leptin. PCOS is closely related to obesity but whether obesity causes PCOS is still undetermined | Narrative review of retrospective studies | Metwally, 2007 |
| **Smoking** | Smoking effects fertility by impairing folliculogenesis and steroidogenesis. The effect of cigarette toxins depends on the amount and duration of exposure. | Systemic review | Dechanet, 2011 |
| **Smoking** | There is a significant increased risk of infertility in women who smoked. Active cigarette smoking is associated with infertility. In some studies, smoking more than 20 cigarettes per day seem to effect fertility. | Systemic Review and metanalysis of observational studies (case-control and cohort) | Augood, 1998 |
| **STIs** | Adhesions cause by PID effects the tubes more than the uterus. Most of these pathogens lead to tubal infertility through an ascending infection.<br>M. genitalium cause salphingitis-PID which may account for infertility. Ascending infection from N. gonorrhoea, C. trachomatis, Gradenella vaginalis lead to tubal factor sterility. Genital amoebiasis can cause damage to the female reproductive system and sterility..<br>HIV adversely effects fertility but it is not understood whether the impact is from the virus or concomitant genital infection or the effect of treatment. | Narrative review | Pellati, 2007 |
| **Medical conditions** | | | |
| **Asthma** | The inflammatory immune response caused by asthma was found in the uterus and tubes of asthmatic women. It causes chronic peripheral inflammation that alters the whole body's inflammatory response. The link that metabolic response is a risk factor for asthma implies that PCOS is related to asthma as well. An imbalance of the adaptive immune system is associated with infertility. | Narrative review | Gade, 2014 |

| | | | |
|---|---|---|---|
| **Cancer** | Cancer-directed therapies reduces the ovarian reserve. Many chemotherapy agents have been linked to ovarian failure and radiation can lead to damage to the reproductive organs. | Narrative review | Levine, 2015 |
| **Chemotherapy** | Female infertility due to ovarian damage from chemotherapy is an inevitable consequence. Chemotherapy causes irreversible and progressive damage to the ovaries and germ cells. Radiotherapy impairs the development of the uterus in young women and increases the risk for ovarian failure. | Narrative review | Lmai, 2007 |
| **Celiac Disease** | Celiac Disease is relevant in women with unexplained infertility. Delayed menarche and amenorrhoea are also symptoms of Celiac Disease. Secondary amenorrhoea and spontaneous abortions were common in women with Celiac Disease. This can be attribute to deficiency of trace elements and vitamins due to malabsorption associated with Celiac Disease, this are responsible for a healthy reproductive life such as abnormal ovarian axis, p | Narrative review | Ozgor, 2010 |
| **Diabetes** | Type I diabetes impacts the reproduction in many ways. Women with Type I diabetes have hypogonadotropic hypogonadism which causes amenorrhoea. Disturbed insulin secretion whether high or low impacts ovarian development and function and can aid in the development of PCOS. Studies on young adult women show preserved ovulation however they found fewer pregnancies and live births. Hyperandrogenism has also been associated with diabetes, | Systemic review | Codner, 2012 |
| **Lupus** | POF in lupus patients can be due to autoimmunity or drug related. Patients with SLE can suffer from menstrual disturbances which has been associated with anti-corpus luteum antibodies which suggests autoimmunity as well | Narrative review | Hickman, 2011 |
| **Sickle cell disease** | Women with sickle cell disease have lower number of pregnancies and delayed menarche. | Narrative review | Smith-Whitley, 2014 |
| **Thyroid diseases** | Both hypothyroidism and hyperthyroidism are linked to menstrual abnormalities ranging from amenorrhoea to menorrhagia and subsequently leading to lower pregnancy rate and infertility. | Narrative review | Poppe, 2007 |

Note. STIs = sexually transmitted infections; PID = pelvic inflammatory disease; PCOS = polycystic ovarian syndrome

Table 2.

Application of Considerations for the Selection of Risk Factors*, as well as Identification and Endorsement Attained in Previous study**

| Risk Factor | Primary causes of disease | Prevalent or hazardous [a] | Potential causality | Data on exposure available | Potentially modifiable | Found in search in LMIC [b] | Endorsed by experts in survey [c] |
|---|---|---|---|---|---|---|---|
| **CSG** | No | Yes | Yes | Yes | Yes | Yes | Yes |
| **FGM/C** | No | Yes | Yes | Yes | Yes | Yes | Yes |
| **HIV** | Yes | Yes | Yes | Yes | Yes | Yes | Yes |
| **GTB** | Yes | Yes | Yes | Yes | Yes | Yes | Yes |
| **BV** | No | Yes | Yes | Yes | Yes | Yes | Yes |
| **D&C** | No | Unknown | Yes | No | Yes | Yes | Yes |
| **Vit D def** | Yes (musculoskeletal) | Yes | Yes | Yes | Yes | Yes | Yes |
| **Waterpipe smoking** | Yes (smoking in general) | Yes | Yes | Yes (smoking and equivalence to smoking) | Yes | No | Yes |

*Note*. *World Health Report, WHO, 2002; Ezzati et al., 2002; **Bayoumi et al., 2018.
[a]Ezzati et al., (2002) suggest that when the risk is not a primary cause of disease, consider the prevalence and or hazardous nature of the RF.
[b] Was the RF found in the preliminary search of the literature reported in Bayoumi et al., 2018.
[c] Was the RF endorsed by fertility experts in the survey reported in Bayoumi et al., 2018.
CSG = consanguinity; FGM/C = female genital mutilation/cutting; GTB = genital tuberculosis; BV = bacterial vaginosis; D&C = dilatation and curettage; Vit D def = vitamin D deficiency.

Table 3.

Bradford Hill Criteria and Definitions

| Criteria | Definition |
|---|---|
| **1. Strength** | A larger associations indicates that causality is more likely, but a small association doesn't mean there is no casual effect |
| **2. Consistency** | The consistency of findings across different studies in different populations and settings, but also molecular level studies bolster the epidemiological evidence from observational studies, decreasing the need for repetitions of observational studies |
| **3. Specificity** | A causal relationship is more likely if the association between a factor and the effect is more specific |
| **4. Temporality** | The cause has to occur before the effect |
| **5. Biological gradient** | The presence of a dose-response (more exposure-more effect) relationship increases the likelihood of a causal relationship |
| **6. Plausibility** | The biological evidence provides a model that helps explain the association of interest |
| **7. Coherence** | Consistency between laboratory and epidemiological findings increases likelihood of a causal relationship, similar to 'consistency' |
| **8. Experiment** | Evidence from experimental manipulation such that cessation of exposure leads to decrease in disease lends strong support to causal relationship |
| **9. Analogy** | Considering the effect of similar factors |

*Note*. Definitions derived from Hill, 1965; Fedak et al., 2015

### Genital Tuberculosis

Plausible mechanisms to explain how GTB could be associated with fertility problems

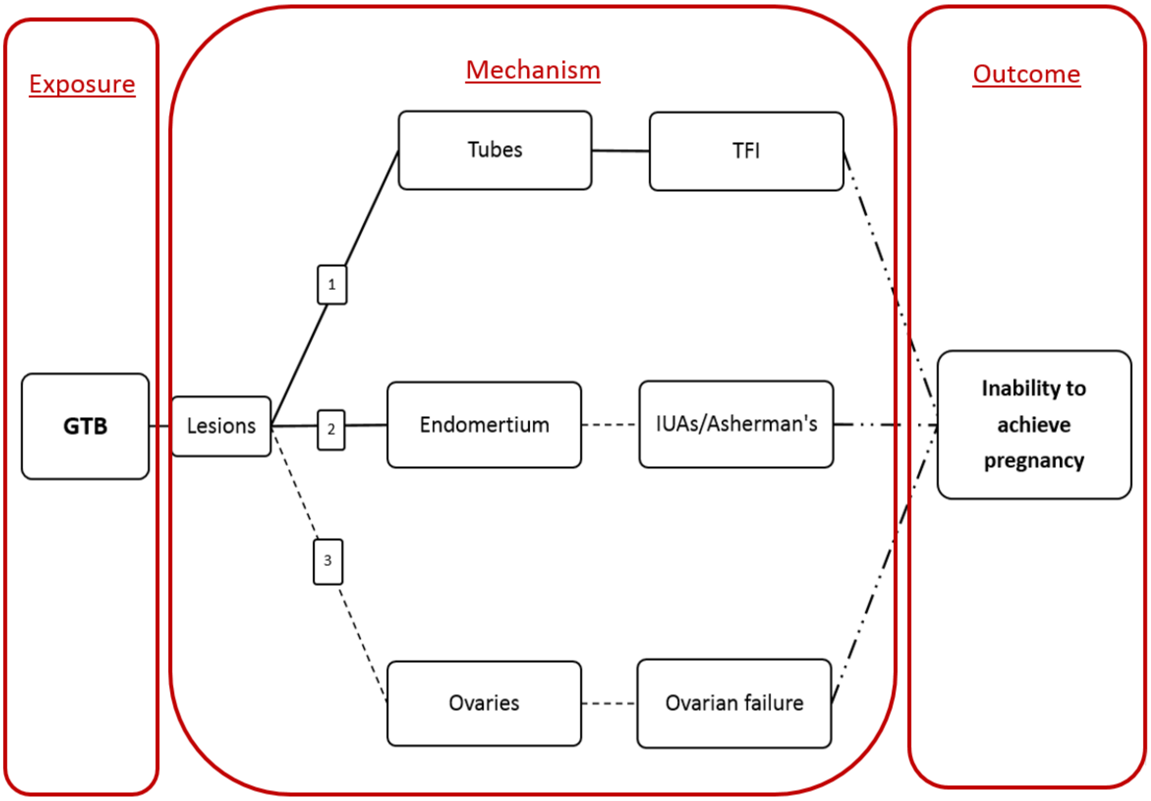


*Figure 2*. Proposed pathways for the impact of Genital Tuberculosis (GTB) on fertility. Solid line = Recent evidence (e.g. imaging); Dashed line = Proposed pathway/historic evidence; Dashed-Dotted line = Well established; TFI = tubal factor infertility; IUAs = intrauterine adhesion

Table 4.

Summary of Reproductive Health Consequences of Genital Tuberculosis (GTB) Reported in the Literature

| **Reproductive Outcome** | **Effect of GTB** | **Statistics reported** (percentage of GTB patients) | **Primary study** | **Review** |
|---|---|---|---|---|
| **Infertility** | Infertility is the presenting or most common complaint | 40 to 50 | Siegler, 1979; Sutherland, 1979; 1983, Bazax-Malik, 1983; Sivanesaratnam, 1986; Punnonen, 1983; Francis 1964; Govan, 1962; Russel, 1951 | Varma, 2008 |
| | Infertility | 64.2 vs. 22 control | Tripathy & Tripathy, 1987 | |
| | | 54.4 | Ojo & Unuigbe, 1987 | |
| | | 10 to 85 | Schaefer 1976, Krishna, 1977; Tripathy & Tripathy, 2002 | Ghosh, 2011 |
| | | NR | Arora, 2003; Choudhary, 1996; Bukulmez, 1999; Bapna, 2005; Varma, 1991; Sharma, 2008; Chavan, 2004; Dam, 2006 | Ghosh, 2011 |
| | | NR | Dhillon, 1990; de Vynck, 1990 | Varma, 2008 |
| | Infertility (primary and secondary) | 42.5 (78 and 22) | Qureshi et al., 2001 | Gatongi, 2005 |
| | Tubal blockage (Peritubal adhesions and tuboovarian masses) | 47.2 | deVynck et al, 1990 | Malik, 2003 |
| **Pelvic pain** | Is not usually severe and present for many months before presenting | 25 to 50 | Falk et al., 1980; Francis, 1964; Sutherland, 1979; Sutherland, 1983 | Varma, 2008 |
| | Progression of GTB increase severity of pelvic pain and is usually aggravated by coitus, exercise, and menses. | NR | Daly & Monif, 1982 | Varma, 2008 |
| | Chronic pelvic pain | 42.5 | Qureshi et al., 2001 | Gatongi, 2005 |
| | Chronic pelvic pain | 15.8 | Samal et al., 2000 | Gatongi, 2005 |
| **Menstrual dysfunction** | Abnormal uterine bleeding | 10 to 40 | Simon et al., 1977; Daly& Monif, 1982 | Varma, 2008 |
| | menorrhagia (very heavy) | 19 | Samal et al., 2000 | Varma, 2008, Ghosh, 2011; Gatongi, 2005 |
| | Oligohypomenorrhea | 54 | Samal et al., 2000 | Varma, 2008; Gatongi, 2005 |
| | Amenorrhea | NR | Sharma, 2008 | Ghosh, 2011 |

| Reproductive Outcome | Effect of GTB | Statistics reported (percentage of GTB patients) | Primary study | Review |
|---|---|---|---|---|
| | Amenorrhea | 14.3 | Samal et al., 2000 | Varma, 2008; Ghosh, 2011; Gatongi, 2005 |
| | | 15 | Qureshi et al., 2001 | Gatongi, 2005 |
| | Dyspareunia (painful sex) | 5 | Qureshi et al., 2001 | |
| | Dysmenorrhoea (painful period) | 12.5 | Qureshi et al., 2001 | |
| | Menstrual irregularities found cases of endometrial TB of which Amenorrhea was the most common | 85 and 43.6 | Tripathy & Tripathy, 1987 | Varma, 2008; Gatongi, 2005 |
| **Asherman's Syndrome** | Uterine adhesions can be the cause of infertility | NR | Sharma, 2008; Bukulmez, 1999 | Ghosh, 2011 |
| **TB in the neonate** | TB can be spread to fetus in utero/delivery from a mother who has GTB (referred to as congenital TB) | NR | Hamadeh,1992; Arora, 2003; Stark, 1997; Cantwell, 1994 | |

*Note*: NR=not reporte

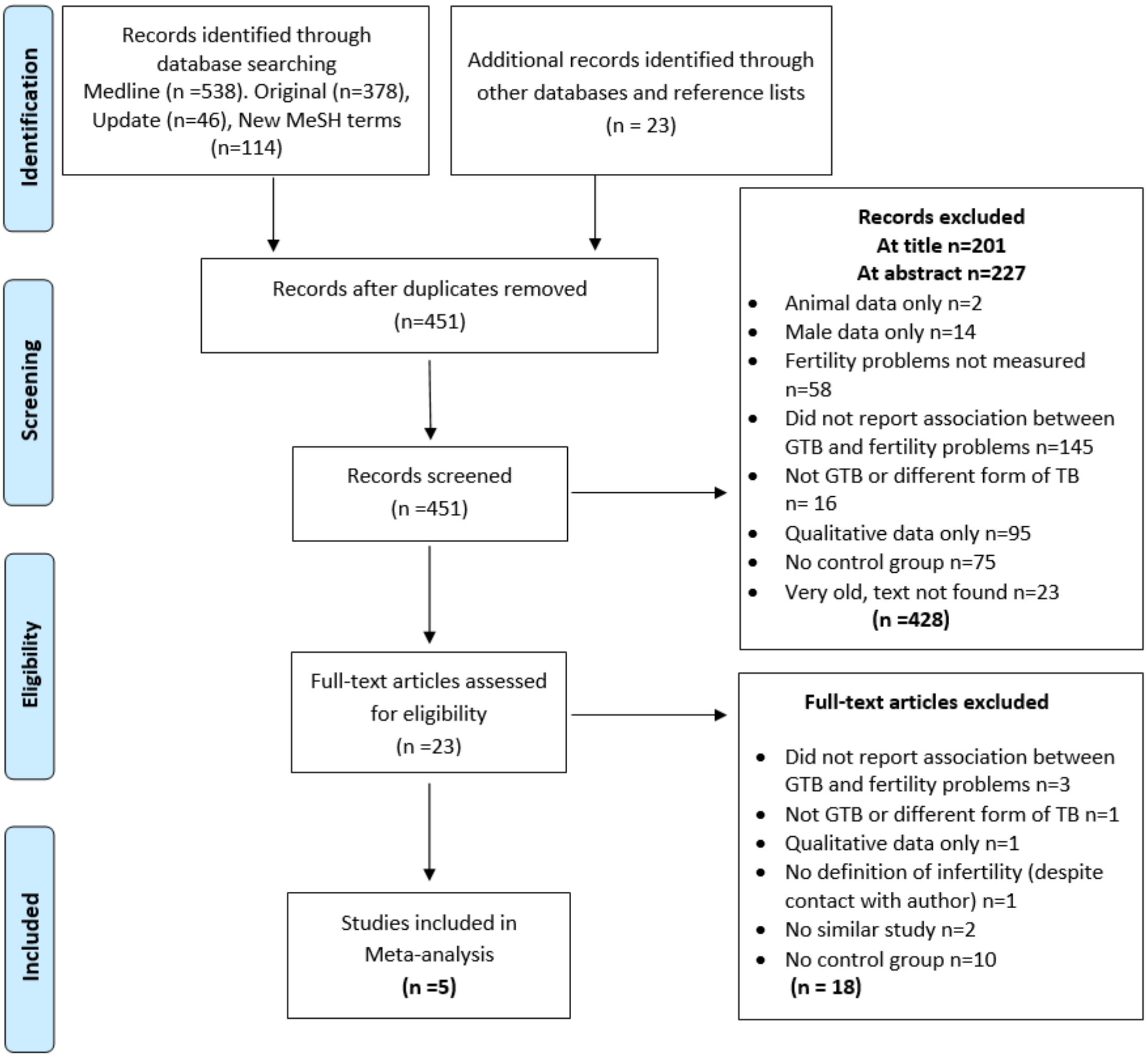


*Figure 3*. PRISMA Flow Diagram for GTB. Figure shows the exclusion of studies at different stages and the reasons for exclusion. Records identified through datbase searching of Medline and Embase includes original search, an update from the time of original search and a search using new MeSH terms. GTB = Genital Tuberculosis

Table 5.

Sample Characteristics of the Seven Included Studies

| Study | Location | Sample (n) | GTB (n) | No-GTB (n) | Age Women [a] | | |
|---|---|---|---|---|---|---|---|
| | | | | | | GTB | No-GTB |
| Ali, 2012 | Kassala, Sudan | 44 women | 25 | 19 | Mean (SD) | 34.8 (6.9) | 34.7 (7.7) |
| Bhanothu, 2014 | India (south) | 302 women | 202 | 100 | Mean (SD) | 28.54 (4.46) | 27.59 (4.62) |
| Sharma, 2011 | India | 388 women | 99 | 289 | Mean (SD) | 28.69 (4.83) | 29.72 (9.58) |
| Malhotra, 2012 | India | 208 women | 104 | 104 | Mean (SD) | 28.7 (3.9) | 28.2 (3.1) |
| Kitilla, 2002 | Ethiopia | 268 women | 67 | 201 | Range | Percentage (n) | Percentage (n) |
| | | | | | 15-19 | 0 | 0.5 (1) |
| | | | | | 20-24 | 19.4 (13) | 8.5 (17) |
| | | | | | 25-29 | 38.8 (26) | 31.3 (63) |
| | | | | | 30-34 | 29.9 (20) | 36.3 (73) |
| | | | | | 35-39 | 11.9 (8) | 19.9 (40) |
| | | | | | 40-44 | 0 | 3.0 (6) |
| | | | | | 45+ | 0 | 0.5 (1) |

*Note*..[a] Age for women at the beginning of the study; GTB = Genital Tuberculosis, SD=Standard deviation; NR= data not reported

Table 6.

Characteristics of the Design of the Seven Included Studies

| | **Study design** | **Data collection** | **Study period** | **GTB measure** | **Infertility outcome measure (duration)** |
|---|---|---|---|---|---|
| Ali, 2012 | Cross-sectional | Maternity Hospital | Jan-Dec 2010 | Clinical symptoms and Histology | Infertility defined as failure to become pregnant despite unprotected sexual practice after one year of marriage. |
| Bhanothu, 2014 | Cross-sectional | 2 Gynaecology clinics | 2006-2014 | Clinical symptoms and Histology | Amenorrhea (duration not specified) |
| Sharma, 2011 | Cross-sectional | University Hospital | 2007-2010 | PCR, Histology, culture, laparoscopy and hysteroscopy | Primary infertility (inability to conceive spontaneously despite one year of regular (3-4 times a week) unprotected intercourse) AND Amenorrhea (duration not specified) |
| Malhotra, 2012 | Cross-sectional | Outpatient Gynaecology clinic | 2007-2009 | PCR, Histology, culture, laparoscopy and hysteroscopy | Primary infertility Secondary infertility |
| Kitilla, 2002 | Cross-sectional | University Hospital | 1995-2000 | Surgical and Histology | TFI (tubo-peritoneal) Primary infertility Secondary infertility |

*Note*. PCR = polymerase chain reaction; TFI = tubal factor infertility

Table 7.

Quality Ratings for the Seven Included Studies on the Basis of an Adapted Newcastle-Ottawa Quality Assessment Scale

| | Quality Criterion | | | | | | |
|---|---|---|---|---|---|---|---|
| | **Adequacy of GTB (exposed) measure[a] Max 2 points** | **Adequacy of control (non-exposed), definition and selection[b] Max 2 points** | **Comparability of control[c] Max 2 points** | **Confounders adequately assessed Max 2 points[d]** | **Adequacy of outcome Infertility measure[e] Max 1 point** | **None response rate or loss to follow-up[f] Max 1 point** | **Overall rating[g]** |
| Ali, 2012 | 2 | 2 | 0 | 0 | 0 | NA | Average |
| Bhanothu, 2014 | 2 | 2 | 0 | 2 | 1 | NA | High |
| Sharma, 2011 | 2 | 1 | 1 | 1 | 0 | NA | Average |
| Malhotra, 2012 | 2 | 2 | 0 | 1 | 0 | NA | Average |
| Kitilla, 2002 | 2 | 1 | 0 | 2 | 0 | NA | Average |

*Note.* [a] GTB was adequately assessed when diagnosis was confirmed by medical testing or hospital records, and it was representative of the cohort i.e. drawn from the same population (up to 2 points); [b] Controls were adequately assessed when selection was comparable to cases, and GTB was excluded properly in the control population (up to 2 points); [c] Comparability of controls was achieved if exposed/non-exposed were matched or adjustment during analysis conducted. One point for rural-urban and one point for any other confounder (up to 2 points); [d] Confounders were adequately assessed if they were obtained from records or a blind interview, and one point was given if the same method was used for both groups (up to 2 points); [e] Infertility outcome was adequately assessed if independent or blind assessment was stated in the paper, or confirmation of the outcome by reference to secure records (medical records, etc.) (up to 1 point); [f] Point given if same rate for both groups and <20% loss to follow up reported; [g] The overall quality rating was low (0 to 3 points), average (4 to 6 points), or high (7 to 10 points).

Table 8.

Number and percentage of women with infertility or amenorrhea in the GTB and No-GTB groups in the included studies (k=5)

| Outcome | | Number of women (%) | |
|---|---|---|---|
| | | **GTB** | **No-GTB** |
| Infertility | | 102/124 (82.3) | 127/308 (41.2) |
| Amenorrhea | | 24/301 (8.0) | 12/389 (3.1) |
| Type of infertility | Primary | 133/171 (77.8) | 149/305 (48.9) |
| | Secondary | 38/171 (22.2) | 156/305 (51.1) |

*Note*. GTB = genital tuberculosis

### Results of Meta-analyses

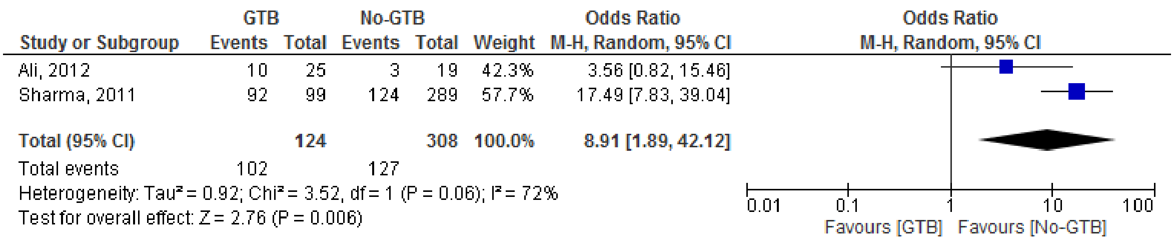


*Figure 4*. Odds ratio for the women who are infertile (>12 months) in the GTB and No-GTB groups

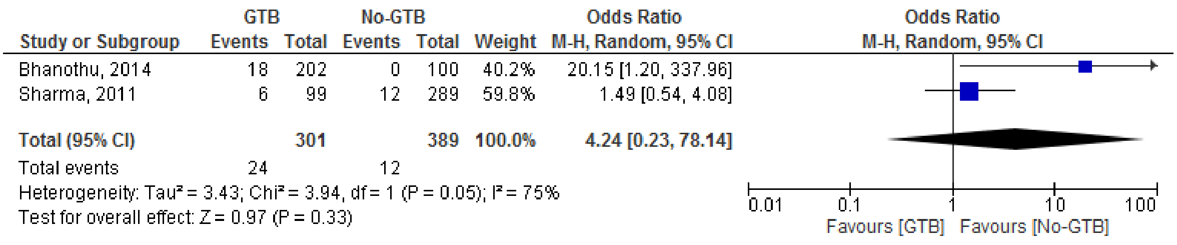


*Figure 5*. Odds ratio for the women reporting 'amenorrhea' (duration not specified) in the GTB and No-GTB groups

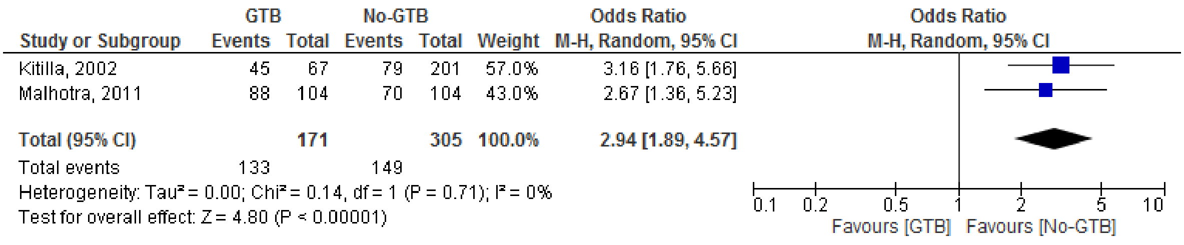


*Figure 6*. Odds ratio for 'primary infertility' amongst infertile women in the GTB and No-GTB groups

### HIV

Table 9.

WHO Clinical Staging of HIV/AIDS for Adults and Adolescents

| Clinical Stage | Clinical Conditions or Symptoms |
|---|---|
| **Primary HIV Infection** | Asymptomatic<br>Acute retroviral syndrome |
| **Clinical Stage 1** | Asymptomatic<br>Persistent generalized lymphadenopathy |
| **Clinical Stage 2** | Moderate unexplained weight loss (<10% of presumed or measured body weight)<br>Recurrent infections (respiratory, Herpes, oral ulceration, Seborrheic dermatitis<br>Fungal nail infections) |
| **Clinical Stage 3** | Unexplained severe weight loss (>10% of presumed or measured body weight)<br>Unexplained chronic diarrhea for >1 month<br>Unexplained persistent fever for >1 month (>37.6°C, intermittent or constant)<br>Persistent oral candidiasis (thrush), Oral hairy leukoplakia<br>Pulmonary tuberculosis (current)<br>Severe presumed bacterial infections (e.g., pneumonia, empyema, pyomyositis, bone or joint infection, meningitis, bacteremia)<br>Unexplained anemia (hemoglobin <8 g/dL)<br>Neutropenia (neutrophils <500 cells/µL)<br>Chronic thrombocytopenia (platelets <50,000 cells/µL) |
| **Clinical Stage 4** | HIV wasting syndrome<br>Recurrent infections (severe bacterial pneumonia, Chronic herpes, Esophageal candidiasis, Extrapulmonary tuberculosis, Cytomegalovirus infection)<br>Cancer (Kaposi sarcoma, Lymphoma, Invasive cervical carcinoma)<br>Central nervous system toxoplasmosis<br>Other severe infections and cancers |

*Note*. Table adapted from Consolidated Guidelines on the Use of Antiretroviral Drugs for Treating and Preventing HIV Infection: Recommendations for a Public Health Approach. 2nd edition. WHO, 2016. https://www.ncbi.nlm.nih.gov/books/NBK374293/ Copyright by WHO [2016]. Reprinted by permission.

**Plausible mechanisms to explain how HIV could be associated with fertility problems**

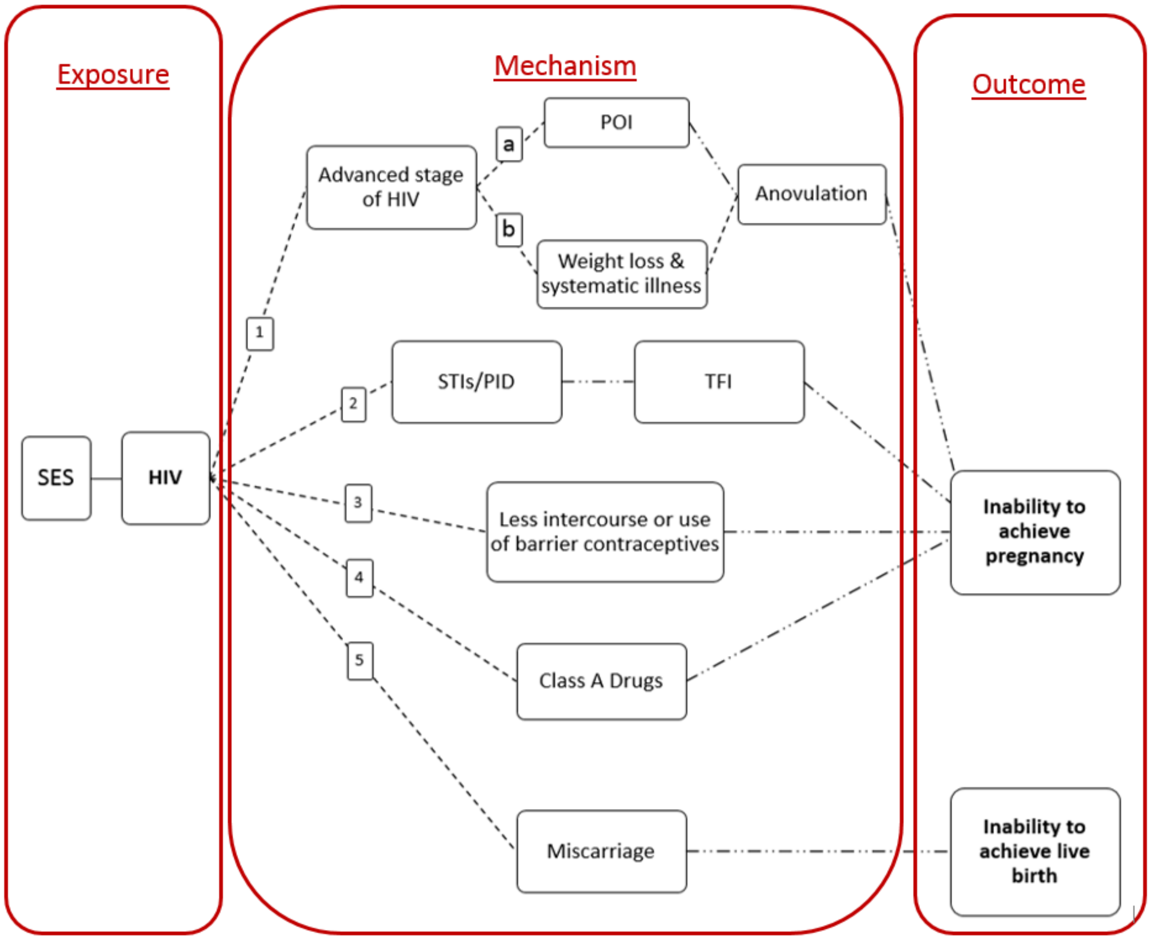


*Figure 7*. Proposed pathways for the impact of HIV on fertility. Solid line = Recent evidence (imaging); Dashed line =Proposd pathway/historic evidence; Dashed-Dotted line = Well established; SES = socioeconomic status; POI = primary; ovarian insufficiency; STIs = sexually transmitted infections; PID = pelvic inflammatory disease; TFI = tubal factor infertility, IUAs = intrauterine adhesion

Table 10.

Summary of Reproductive Health Consequences of HIV Reported in the Literature

| **Reproductive outcome** | **Effect of HIV** | **Primary study** | **Statistics reported (where available)** | **Review** |
|---|---|---|---|---|
| **Ovarian function** | Change in ovarian reserve in HIV+ women Mixed results | Schoenbaum et al. (2005); Martinet et al. (2006) reported normal ovarian reserve | NR | van Leeuwen et al. (2007) |
| | | Clark et al. (2001); Englert et al. (2004) reported dramatically reduced ovarian function i.e. Primary ovarian insufficiency (POI) | NR | |

| **Reproductive outcome** | **Effect of HIV** | **Primary study** | **Statistics reported (where available)** | **Review** |
|---|---|---|---|---|
| | FSH level | Clark et al. (2001) report higher rates of elevated FSH | 8% of HIV+ women (20-42yrs) had FSH level indicative of menopause | Kushnir and Lewis (2011) |
| | | Cejtin et al. (2006) reported no difference in FSH in women with amenorrhea | NR | Kushnir and Lewis (2011) |
| | | Seifer et al. (2007) found no evidence that HIV infection influences ovarian aging (FSH and AMH levels) | NR | Kushnir and Lewis (2011) |
| | Ovaries susceptible to HIV and secondary infections | Not well studied but hypothetically i.e. no specific evidence | NR | Lo and Schambelan (2001) |
| **Menstrual cycle** | Menstrual irregularities (very short and very long) in HIV+ women without AIDS | Chirgwin et al. (1996) | NR | van Leeuwen et al. (2007); Lo and Schambelan (2001); Waters et al. (2007) |
| | Increased rate of menstrual irregularities in HIV infected women with AIDS (and the associated wasting). | Harlow et al. (2000) | NR | van Leeuwen et al. (2007) |
| | | Grinspoon et al. (1997) | NR | Lo and Schambelan (2001) |
| | HIV+ had little effect on menstrual irregularities (cycle length/ menstrual duration) | Harlow et al. (2000) | NR | Lo and Schambelan (2001); Waters et al. (2007) |
| | | Harlow et al. (2000); Chirgwin et al. (1996) | NR | Waters, et al. (2007); van Leeuwen et al. (2007) |
| | Among HIV+ women, increased cycle variability was associated with higher viral loads and lower CD4 cell counts | Harlow et al. (2000) | NR | Lo and Schambelan (2001); Waters et al. (2007) |
| | | Clark et al. (2001) | NR | Waters et al. (2007) |
| **Amenorrhea** | Prolonged amenorrhea without ovarian failure | Cejtin et al. (2006) | HIV+ women 3 times more likely to have prolonged amenorrhea without ovarian failure | Kushnir and Lewis (2011) |
| | Increased rate of amenorrhea | Chirgwin et al. (1996) | NR | Lo and Schambelan (2001); Kushnir and |

| Reproductive outcome | Effect of HIV | Primary study | Statistics reported (where available) | Review |
|---|---|---|---|---|
| | | | | Lewis (2011); Waters et al. (2007) |
| | Being HIV+ had little overall impact on amenorrhea | Harlow et al. (2000) | NR | Lo and Schambelan (2001); Waters et al. (2007) |
| | | Ellerbrock et al. (2007); Harlow et al. (2000) | NR | Kushnir and Lewis (2011) |
| **Comorbid STIs** | A high incidence of comorbid STIs in HIV+ | Paxton et al. (1998); Gray et al. (1998); Wawer et al. (1998) | NR | Kushnir and Lewis (2011) |
| | | Frankel et al. (1997); Sobel (2000) | NR | van Leeuwen et al. (2007) |
| **Tubal blockage** | Higher rates of tubal blockage | Frodsham et al. (2006) | NR | Waters et al. (2007) |
| | Higher STIs suggesting that women who are HIV+ may be at increased risk of tubal damage. | Frankel et al. (1997); Sobel (2000) | NR | van Leeuwen et al. (2007) |
| | Tubal occlusion | Coll et al. (2007) | 27.8% among HIV+ women | Kushnir and Lewis (2011) |
| **Pregnancy rate** | Lower pregnancy rate in HIV+ women | Zaba et al. (1998) [Africa] | fertility was 25% to 40% lower in HIV+ women in Sub-Saharan Africa | Kushnir and Lewis (2011) |
| | | Massad et al. (2004) [USA] | NR | |
| | | Stephenson et al. (1996); Thackway et al. (1997); De Vincenzi et al. (1997) | NR | Lo and Schambelan (2001) |
| | | Zaba and Gregson (1998) (Regardless of STIs) | NR | van Leeuwen et al. (2007) |
| | Dramatic decline in pregnancy rate in HIV+ women with increased progression of the disease | Sedgh et al. (2005) | NR | van Leeuwen et al. (2007) |
| **Birth rate** | Lower birth rate in HIV+ women | Stephenson et al. (1996); Thackway et al. (1997); De Vincenzi et al. (1997) | NR | Lo and Schambelan (2001) |
| **Abortions/miscarriage** | Pregnancy loss was more common among HIV+ women | Gray et al. (1998) | HIV+ vs. HIV- (18.5% vs.12.2%) | Kushnir and Lewis (2011) |
| | Before HAART pregnancy loss was much more common among HIV+ women | D'Ulbaldo et al. (1998) | 67% higher among HIV+ | Kushnir and Lewis (2011) |

| Reproductive outcome | Effect of HIV | Primary study | Statistics reported (where available) | Review |
|---|---|---|---|---|
| | Miscarriage rate remained constant from 1990 through 2006 despite evolution of therapy during this period | Townsend et al. (2008) | Miscarriage rate of 4% | Kushnir and Lewis (2011) |
| | Higher rates of abortion | Stephenson et al. (1996); Thackway et al. (1997); De Vincenzi et al. (1997) | NR | Lo and Schambelan (2001) |

*Note***.** NR = not reported; POI = Primary Ovarian Insufficiency; FSH = Follicle-Stimulating Hormone; AMH = Antimullerian hormone; CD4 = Type of white blood cell; STIs = Sexually Transmitted Infections; PID = Pelvic Inflammatory Disease

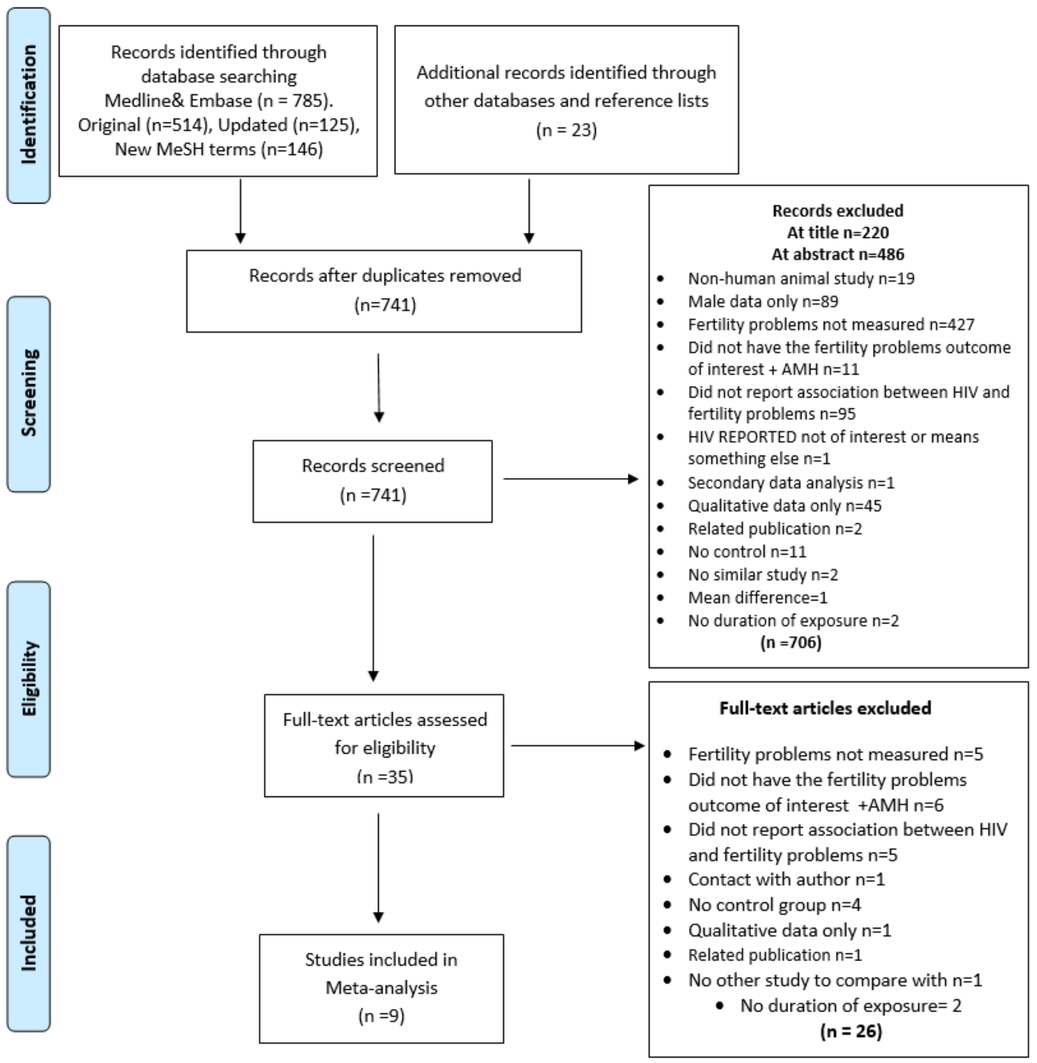


*Figure 8*. PRISMA Flow Diagram for HIV. Figure shows the exclusion of articles at the different stages and the reasons for exclusion. Records identified through datbase searching of Medline and Embase includes original search, an update from the time of original search and a search using new MeSH terms.

Table 11.

Sample Characteristics Reported in the Ten Included Studies

| Study | Location | Sample (n) | N | N | Age [a] Women | | |
|---|---|---|---|---|---|---|---|
| **Cohort/cross-sectional studies** | | | **HIV** | **No-HIV** | | **HIV** | **No-HIV** |
| Cejtin, 2006 | USA | 1431 women | 1145 women | 286 women | Range | Percentage (n) | Percentage (n) |
| | | | | | 16–39 | 59.1 (677) | 63.6 (182) |
| | | | | | 40–44 | 25.2 (288) | 25.5 (73) |
| | | | | | 45–49 | 11.4 (131) | 8 (23) |
| | | | | | 50–55 | 4.3 (49) | 2.8 (8) |
| Chirgwin, 1996 | USA | 330 women | 248 women | 82 women | Mean (SD) | 32.7 (6.2) | 34.5 (6.9) |
| Gray, 1998 | Uganda | 4497 women | 953 women | 3544 women | Range | Percentage (n) | Percentage (n) |
| | | | | | 15–19 | 23·9 (847) | 7·1 (68) |
| | | | | | 20–24 | 26·5 (938) | 30·7 (293) |
| | | | | | 25–29 | 16·3 (578) | 30·6 (292) |
| | | | | | 30–39 | 21·9 (775) | 26·1 (249) |
| | | | | | >40 | 11·5 (406) | 5·4 (51) |
| Linas, 2011 | USA | 1412 women | 941 women | 471 women | NR | NR | NR |
| Willems, 2013 | Burkina Faso | 93 women | 54 women | 39 women | Mean (SD) | 35 (5) | 29 (6.5) |
| Ross 2003 | Uganda | 216 women | 81 women | 135 women | NR | NR | NR |
| Yaro 2001 | Burkina Faso | 912 women | 63 women | 849 women | Mean (SD) | 16.7 ±2 | 16.9±2 |
| Ezechi 2010 | Markurdi, Nigeria | 3473 women | 2549 women | 924 women | Mean age | 32.7± 4.9 | 33.2±5.7 |
| **Case-control studies** | | | **Infertile [b]** | **Fertile (control)** | | **Infertile** | **Fertile** |
| De Muylder, 1990 | Zimbabwe | 331 women | 227 | 104 | Mean (SD) | 28.4 (4.8) tubal<br>27.1 (4.9) non-tubal | NR |
| Dhont, 2010 | Rwanda | 595 women | 312 | 283 | Median (IQR) | 30 (27–35) | 27 (24–31) |

*Note*. [a] Age for women at the beginning of the study; [b] Unable to become pregnant after at least 12 months of unprotected intercourse; NR= data not reported; SD=Standard deviation; IQR=inter-quartile range

Table 12.

Characteristics of the Design of the Ten Included Studies

| Study | Study design | Data collection | Study period | HIV self-report or Blood test | Fertility Problems outcome measure (duration) |
|---|---|---|---|---|---|
| Cejtin, 2006 | Cross-sectional data embedded in a Cohort study | Interagency HIV Study Hospital/clinic based | 1994-1997 | Blood test (type not specified) | Amenorrhea > 12 months And/or FSH > 25 (mUI/ml) |
| Chirgwin, 1996 | Cross-sectional data embedded in a Cohort study | Hospital/clinic based | 1991-1994 | Blood test (type not specified) | Amenorrhea > 3 months |
| Gray, 1998 | Cross-sectional data embedded in a Cohort study | Community based | 1994-1995 | Blood test (Western-blot) | Pregnancy rate per woman (we converted to no-pregnancy) |
| Linas, 2011 | Cohort | Interagency HIV Study Hospital/clinic based | 2002-2009 | Blood test (HIV RNA, CD4 count and Serology) | Pregnancy rate per woman (we converted to no-pregnancy) |
| Willems, 2013 | Cross-sectional data | Hospital/clinic based | 2008 | Blood test (ELISA and Western-blot) | FSH > 40 (mUI/ml) |
| De Muylder, 1990 | Case-control | Hospital based | 1985-1987 | Blood test (ELISA and Western-blot) | More than 18 months unprotected sex |
| Dhont, 2010 | Case-control | Hospital based & community | 2007-2009 | Blood test (Rapid test) | More than 12 months unprotected sex |
| Ross 2003 | Cohort study | Clinic based | 1990-2001 | Records (CD4 count & WHO staging) | Foetal loss- spontaneous abortion and still birth |
| Yaro 2001 | Cross-sectional | Clinic based | 1988 | Blood test (type not specified) | Live birth, still birth, abortion |
| Ezechi 2010 | Cross-sectional | Research institute & medical centre | 2005-2007 | Blood test (ELISA, Western bolt, CD4 count and viral load) | Amenorrhoea > 90 days |

*Note*. HIV = Human Immunodeficiency Virus; CD4 = cluster of differentiation 4; RNA = Ribonucleic Acid ; FSH = Follicle Stimulating Hormone; ELISA = Enzyme-linked Immunosorbent Assay

Table 13.

Quality Ratings for the Ten Included Studies on the Basis of an Adapted Newcastle-Ottawa Quality Assessment Scale

| Study | Quality Criterion | | | | | | Overall rating [g] |
|---|---|---|---|---|---|---|---|
| | Adequacy of HIV (exposed) measure [a] Max 2 points | Adequacy of control (non-exposed), definition and selection [b] Max 2 points | Comparability of control [c] Max 2 points | Confounders adequately assessed Max 2 points [d] | Adequacy of outcome Fertility Problems measure [e] Max 1 point | None response rate or loss to follow-up [f] Max 1 point | |
| **Cejtin, 2006** | **2** | **2** | **1** | **2** | **1** | **0** | **High** |
| **Chirgwin, 1996** | 2 | 2 | 1 | 1 | 1 | NA | High |
| **Gray, 1998** | 2 | 2 | 1 | 2 | 1 | NA | High |
| **Linas, 2011** | 2 | 2 | 1 | 2 | 0 | NR | High |
| **Willems, 2013** | 2 | 2 | 1 | 2 | 1 | NA | High |
| **De Muylder, 1990** | 2 | 2 | 0 | 1 | 1 | NA | Average |
| **Dhont, 2010** | 2 | 2 | 1 | 2 | 1 | NA | High |
| **Ross 2003** | 2 | 1 | 1 | 2 | 0 | NR | Average |
| **Yaro 2001** | 2 | 2 | 1 | 1 | 0 | NA | Average |
| **Ezechi 2010** | 2 | 2 | 1 | 1 | 1 | NA | High |

*Note***.** NA = not applicable; NR = not reported; [a] HIV was adequately assessed when independent validation of the diagnosis (e.g. blood testing and/or hospital/medical records) and it was representative of the cohort i.e. drawn from the same population (up to 2 points); [b] Controls were adequately assessed when selection was comparable to cases, and HIV was excluded properly in the control population (up to 2 points); [c] Comparability of controls was achieved if exposed/non-exposed were matched or adjustment during analysis conducted. One point for *age* and one point for any other confounder (up to 2 points); [d] Confounders were adequately assessed if they were obtained from records or a blind interview, and one point was given if the same method was used for both groups (up to 2 points); [e] Fertility problems outcome was adequately assessed if independent or blind assessment was stated in the paper, or confirmation of the outcome by reference to secure records (medical records, etc.) (up to 1 point); [f] Point given if same rate for both groups and <20% loss to follow up reported; [g] The overall quality rating was low (0 to 3 points), average (4 to 6 points), or high (7 to 10 points).

Table 14.

Number and Percentage of Women with a Specific Outcome in the HIV+ and HIV- Groups in the Included Studies (k=9)

| Outcome | Number of women (%) | |
|---|---|---|
| | HIV+ | HIV- |
| Pregnancy | 532 of 1894 (28.1) | 1120 of 4015 (27.9) |
| Amenorrhoea | 173 of 3942 (4.4) | 22 of 1292 (1.7) |
| FSH >25 IU/l | 60 of 1194 (5.0) | 10 of 317 (3.2) |
| Infertile > 12 months | 107 of 146 (73.3) | 432 of 780 (55.4) |
| Miscarriage | 26 of 155 (16.8) | 99 of 948 (10.4) |

*Note*. FSH = follicle-stimulating hormone

### Results of Meta-analyses

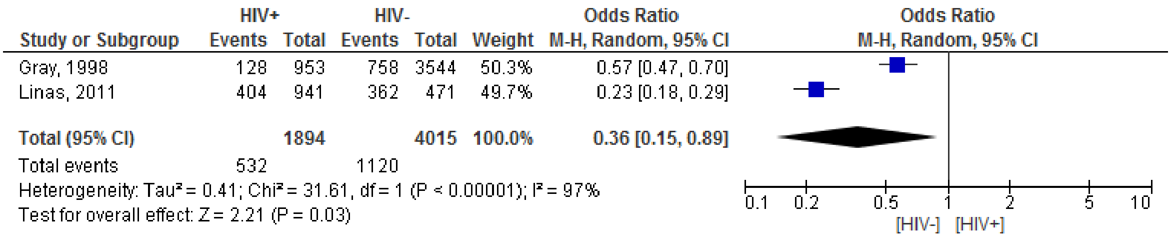


*Figure 9*. Odds ratio for women reporting 'pregnancy' in the HIV+ and the HIV- groups

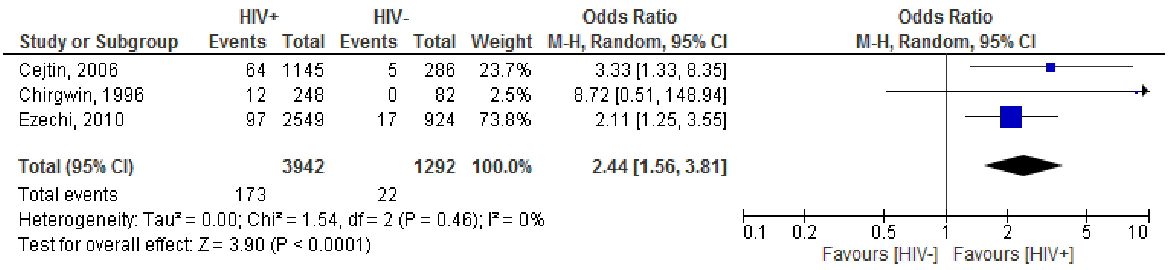


*Figure 10*. Odds ratio for the proportion of women who have amenorrhea (>3 months) in HIV+ vs HIV- women

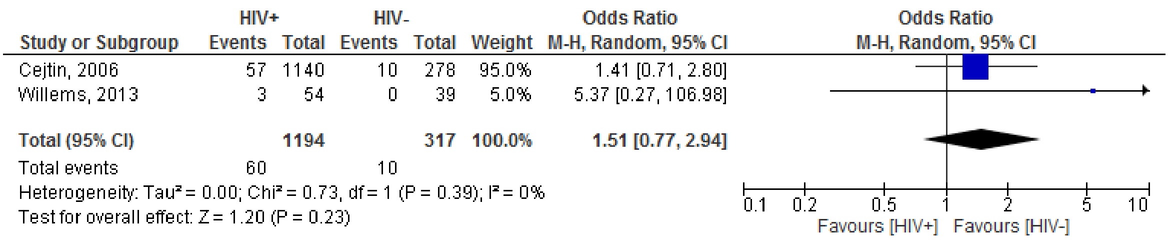


*Figure 11*. Odds ratio for proportion of women who have 'Level of FSH >25 IU/l' (indicative of POI) in the HIV+ and HIV- groups

| Study or Subgroup | HIV+ Events | HIV+ Total | HIV- Events | HIV- Total | Weight | Odds Ratio M-H, Random, 95% CI |
|---|---|---|---|---|---|---|
| De Muylder, 1990 | 9 | 9 | 218 | 322 | 2.1% | 9.09 [0.52, 157.62] |
| Dhont, 2010 | 98 | 137 | 214 | 458 | 97.9% | 2.87 [1.89, 4.33] |
| **Total (95% CI)** | | **146** | | **780** | **100.0%** | **2.93 [1.95, 4.42]** |
| Total events | 107 | | 432 | | | |

Heterogeneity: Tau² = 0.00; Chi² = 0.63, df = 1 (P = 0.43); I² = 0%
Test for overall effect: Z = 5.15 (P < 0.00001)

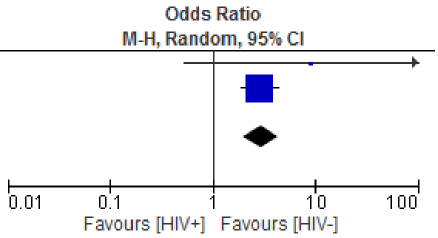


*Figure 12*. Odds ratio proportion of women who are 'infertile > 12 months' in the HIV+ and HIV- women

| Study or Subgroup | HIV+ Events | HIV+ Total | HIV- Events | HIV- Total | Weight | Odds Ratio M-H, Random, 95% CI |
|---|---|---|---|---|---|---|
| Ross, 2003 | 19 | 92 | 14 | 99 | 53.8% | 1.58 [0.74, 3.37] |
| Yaro, 2001 | 7 | 63 | 85 | 849 | 46.2% | 1.12 [0.50, 2.54] |
| **Total (95% CI)** | | **155** | | **948** | **100.0%** | **1.35 [0.77, 2.35]** |
| Total events | 26 | | 99 | | | |

Heterogeneity: Tau² = 0.00; Chi² = 0.36, df = 1 (P = 0.55); I² = 0%
Test for overall effect: Z = 1.06 (P = 0.29)

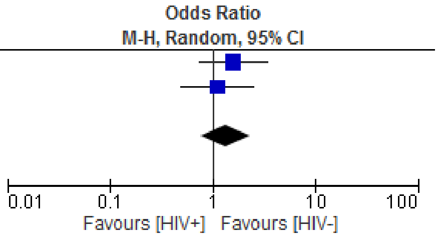


*Figure 13*. Odds ratio for the proportion of women who had a 'miscarriage' in the HIV+ and HIV- women

**Bacterial Vaginosis**

Table 15.

Clinical and Laboratory Approaches, Criteria and Evaluation for the Diagnosis of Bacterial Vaginosis

| Approach | Criteria | Evaluation |
| --- | --- | --- |
| **Amsel criteria (clinical)** | (1) Thin, white, homogeneous discharge<br>(2) Clue cells on microscopy of wet mount 5<br>(3) pH of vaginal fluid >4.5<br>(4) Release of a fishy odour on adding alkali (10% KOH) | At least three of the four criteria are present for the diagnosis to be confirmed |
| **Gram stained vaginal smear (laboratory)** | Grade 1 (Normal): Lactobacillus morphotypes predominate<br>Grade 2 (Intermediate): Mixed flora with some Lactobacilli present, but Gardnerella or Mobiluncus morphotypes also present<br>Grade 3 (BV): Predominantly Gardnerella and/or Mobiluncus morphotypes. Few or absent Lactobacilli | To be evaluated with the Nugent criteria or the Hay/Ison criteria |

*Note*. BV = bacterial vaginosis; UK guidelines for the management of BV (Hay, Patel & Daniels, 2012)

Plausible mechanisms to explain how BV could be associated with fertility problems

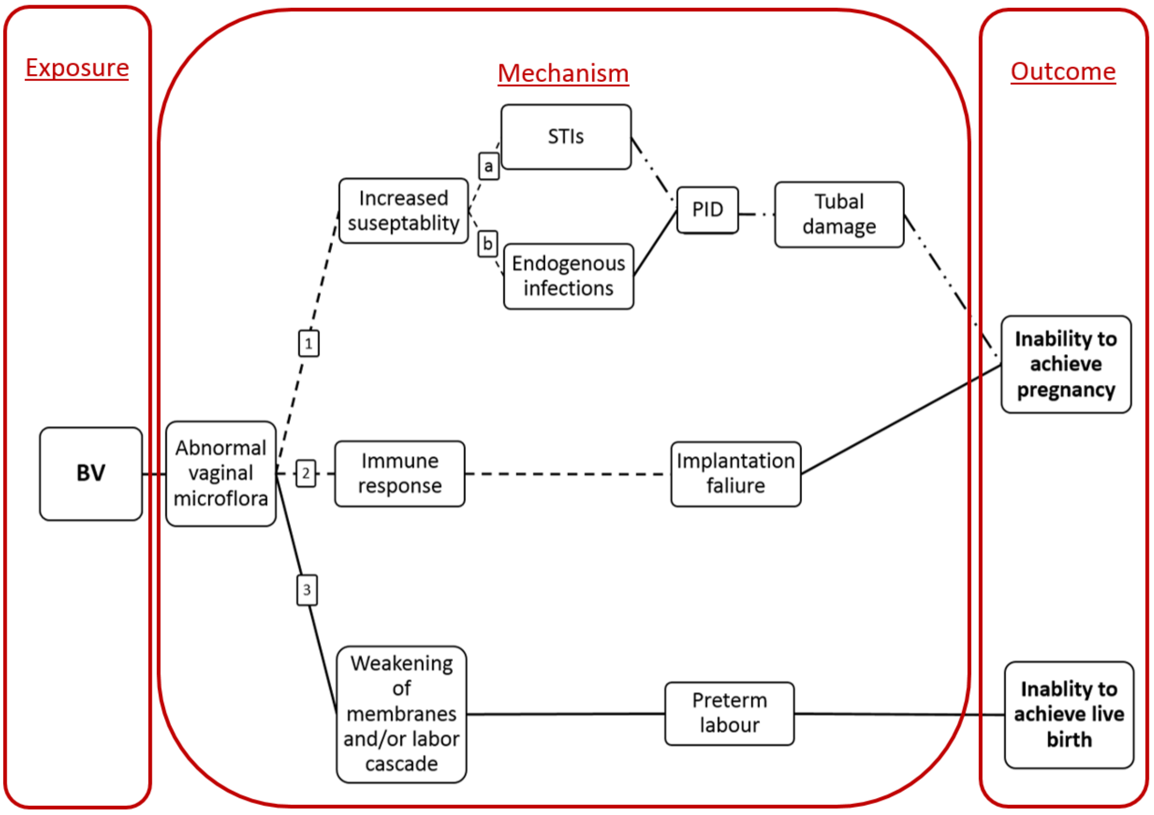


*Figure 14*. Proposed pathways describing potential impact of Bacterial Vaginosis (BV) on fertility. Solid line = Recent evidence (molecular level laboratory); Dashed line = Proposed pathway/historic evidence; Dashed and dotted = Well established

Table 16.

Summary of Reproductive Health Consequences of Bacterial Vaginosis Reported in the Literature

| Reproductive outcome | Effect of BV | Primary study | Statistics reported | Review |
|---|---|---|---|---|
| **Preterm labour/ delivery** | Women with BV at increased risk of preterm birth | Hillier, et al., 1995<br><br>McGregor, et al., 1995<br>Hay, et al., 1994<br>Hillier, et al. 1995<br>Hauth, Goldenberg, Andrews, DuBard & Copper, 2001 | ORs between 1.8 and 6.9<br><br><br><br>Attributable risks between 2-10 for BV in pregnancy leading to preterm delivery (women with no previous history) and over 30 (women with a history of a previous preterm birth) | Hay, (2004)<br><br>Hay, (2004)<br>Hay, et al., (2012)<br>Morris et al., (2001) |
| | The strong association between BV and loss before 20 weeks was confirmed in women examined at less than 14 weeks' gestation (Belgium) | Donders, et al., 2000 | RR= 5.4 | Hay, (2004) |
| | The overall risk of preterm birth for women with BV was determined in meta-analysis of 20 232 pregnancies | Leitich, et al., 2003 | Studies that screened before 16 weeks' gestation OR= 7.55,<br>Studies that screened before 20 weeks gestation OR= 4.20 | Hay, (2004) |
| | Preterm labour due to chorioamnionitis found to be related to organisms associated with BV | Hillier, et al., 1988<br>Heller, Moorehouse-Moore, Skurnick & Baergen, 2003<br>Sebire, 2001<br>Goldenberg, Hauth & Andrews, 2000 | NR | Hay, (2004)<br>Hay, et al., (2012) |
| | Release of enzymes by bacteria, allowing penetration of mucus and weakening of the membranes, leading to preterm labour | Howe, et al., 1999 | NR | Hay, (2004) |
| | Alterations in vaginal microbiology associated with late miscarriage and premature birth | Koumans, Markowitz & Hogan, 2002 | NR | Mastromarino, et al., (2014) |
| **Miscarriage** | Higher risk for preclinical pregnancy loss in women who had BV than those who didn't (conceived by IVF) | van Oostrum, et al., (2013) Meta-analysis | (OR 2.36, 95% CI 1.24 to 4.51). | van Oostrum, et al., (2013) |
| | Even if BV resolves during pregnancy, that doesn't reduce risk of miscarriage and preterm labour | Riduan, et al., 1993<br>Lamont, Duncan, Mandal & Bassett, 2003 | NR | Hay, (2004) |
| | More first trimester miscarriage in women with BV in a sample of women who conceived with IVF treatment, even after adjusting for factors known to increase risk of miscarriage | Ralph, Rutherford & Wilson, 1999 | First trimester miscarriage was 31.6% for those with BV compared with 18.5% for those with normal vaginal flora (crude odds ratio 2.49, 1.21 to 5.12) | Hay, (2004)<br>Morris et al., (2001) |
| | In study on natural conception BV was associated with miscarriage early in the second trimester 13-15 weeks, but not at 10-12 weeks | Oakeshott, et al., 2002 | 13–15 weeks' gestation (OR 3.5; 1.2–10.3)<br>10 and 12 weeks gestation (OR 1.32; 0.67–2.62) | Hay, (2004) |

| Reproductive outcome | Effect of BV | Primary study | Statistics reported | Review |
|---|---|---|---|---|
| **PID** | BV found to be more common in women with PID. | Moi, 1990<br>Taylor, 1997<br>Soper, Brockwell, Dalton & Johnson, 1994<br>Larsson, Platz-Christensen, Thejls, Forsum & Pahlson, 1992 | NR | Morris, et al., (2001)<br>Hay, et al., (2012) |
| | BV related organisms have been isolated from the endometrium and fallopian tubes of women with PID | Sweet, 1987 | NR | Hay, (2004) |
| | Increased risk of PID in women with BV (using only clinical diagnosis for PID) | Eschenbach, et al., 1988 | Nine-fold | Morris, et al., (2001) |
| | Increased risk of PID in women with BV (using gold standard laparoscopy to diagnose PID) | Peipert, Montagno, Cooper & Sung, 1997 | Three-fold | Morris, et al., (2001) |
| | BV associated with a markedly increased risk for development of PID | Ness, et al., 2005 | NR | Hay, et al., (2012) |
| **Endometritis** | Endometritis more in women with BV than without | Hillier, et al., 1996 | (OR 15, 95% CI 2-686) | Morris, et al., (2001) |
| | Microorganisms associated with BV were isolated more from the endometria of women with than without plasma cell endometritis | Korn, et al., 1995 | (OR 12.4) | Morris, et al., (2001)<br>Hay, (2004) |
| | BV associated with post-partum endometritis | Watts, Krohn, Hillier & Eschenbach, 1990 | NR | Hay, et al., (2012) |
| **Infertility** | Significantly moreBV in women attending infertility clinic than attending antenatal clinic | van Oostrum, 2013<br>Meta-analysis | (OR 3.32, 95% CI 1.53 to 7.20) | van Oostrum, et al., (2013) |
| | In women undergoing IVF more BV in women with TFI than those with non-TFI | Gaudoin, Rekha, Morris, Lynch & Acharya, 1999<br>Liversedge, et al., 1999 | NR | Morris, et al., (2001) |
| | Preclinical pregnancy loss following IVF higher in infertility patients with BV than those with no BV | van Oostrum, et al., (2013) | (OR 2.36, 95% CI 1.24 to 4.51). | van Oostrum, et al., (2013) |
| | BV more common in women with TFI than other types of infertility in sample of women undergoing IVF | Liversedge, et al., 1999 | BV more common in women with TFI (31.5%) than non-TFI (19.7%) infertility (OR 1.87) | Hay, (2004) |
| | | Wilson, Ralph & Rutherford, 2000. | Compared with endometriosis (OR 3.63, 95% CI 1.52–8.67), male factor (OR 2.98, 95% CI 1.80–4.90), and unexplained infertility (OR 2.20, 95% CI 1.35–3.59) [adjusted ORs] | Morris, et al., (2001)<br>Hay, (2004) |
| | Significantly more BV in women with TFI as compared to other causes of infertility in sample of women undergoing IVF | van Oostrum, et al., (2013) | (OR 2.77, 95% CI 1.62 to 4.75) | van Oostrum, et al., (2013) |
| | Significantly more BV in women with anovulation than other types of infertility (but less than TFI) in sample of women undergoing IVF | Wilson, Ralph & Rutherford, 2000. | Compared with endometriosis (OR 3.77, 95% CI 1.28–11.08), male factor (OR 3.09, 95% CI 1.37–6.96), and unexplained infertility (OR 2.29, 95% CI 1.02–5.12) [adjusted ORs] | Morris, et al., (2001)<br>Hay, (2004) |
| | A correlation between bacterial vaginosis, immune response and idiopathic infertility demonstrated in sample of women undergoing IVF | Spandorfer, Neuer, Giraldo, Rosenwaks & Witkin, 2001 | NR | Mastromariano, et al., (2014) |

| Reproductive outcome | Effect of BV | Primary study | Statistics reported | Review |
|---|---|---|---|---|
| **Increased susceptibility to infections** | More HIV+ in women with severe BV (score of 9-10 on a Gram stain) than those with normal vaginal flora in Uganda | Wawer, et al., 1999<br>Sewankambo, et al., 1997 | (OR 2.08, 95% CI 1.48-2.94) | Morris, et al., (2001) |
| | Women with BV significantly more likely to seroconvert before giving birth and after giving birth (Malawi) | Taha et al., 1998 | (OR 3.7, P = 0.03) before giving birth<br>(OR 2.3, P = 0.04) after giving birth | Morris, et al., (2001)<br>Hay, et al., (2012) |
| | Women with abnormal flora on Gram's stain at increased risk of HIV acquisition (Kenya) | Martin, et al., 1999 | (HR = 1.9, 95% CI 1.1-3.1) | Morris, et al., (2001) |
| | Absence of lactobacilli, characteristic of BV and associated with an increased risk of HIV | Martin, et al., 1999 | (HR = 2.0; 95% CI 1.2-3.5) | Morris, et al., (2001)<br>Mastromariano, et al., (2014) |
| | Pregnant women with abnormal vaginal flora at increased risk of HIV seroconversion (North Carolina, USA) | Royce, Thorp, Granados & Savitz, 1999 | (RR 4.0, 95% CI 1.1-14.9) | Morris, et al., (2001) |
| | BV is associated with a markedly increased risk for acquisition of HIV | Cu-Uvin, et al., 2001<br>Schwebke, 2003<br>Atashili, Poole, Ndumbe, Adimora & Smith, 2008 | NR | Hay, et al., (2012) |
| | BV risk factor for female to male HIV transmission | Cohen, et al., 2012 | adjusted OR (3.06, 1.35-6.95) | Hay, et al., (2012) |
| | BV associated with a markedly increased risk for acquisition STIs | Martin, et al., 1999<br>Cherpes, Meyn, Krohn, Lurie & Hillier, 2003<br>Harmanli, Cheng, Nyirjesy, Chatwani & Gaughan, 2000 | NR | Mastromariano, et al., (2014) |
| | Abnormal vaginal flora lacking lactobacilli facilitates infection by parasites e.g. Trichomonas vaginalis and bacteria e.g. Neisseria gonorrhoea and Chlamydia trachomatis | Wiesenfeld, Hillier, Krohn, Landers &<br>Sweet, 2003 | NR | Mastromariano, et al., (2014) |
| | Absence of vaginal lactobacilli, is an independent risk factor for acquisition of herpes simplex virus | Cherpes, Meyn, Krohn, Lurie & Hillier, 2003 | NR | Mastromariano, et al., (2014) |
| **Cervical intraepithelial neoplasia** (changes in the squamous cells of the cervix.) | Association between BV and CIN (suggested to be caused by nitrosamines produced by the abnormal vaginal microflora) | Hudson, Tidy, McCulloch & Rogstad, 1997<br>Pavic, 1984 | NR | Morris, et al., (2001) |
| | Significantly more BV in women with CIN | Uthayakumar, Boyle, Barton, Nayagam & Smith, 1998 | NR | Morris, et al., (2001) |

*Note:* BV = bacterial vaginosis; OR = odds ratio; RR = risk ratio; NR = not reported; IVF = in vitro fertilization; PID = pelvic inflammatory disease; TFI = tubal factor infertility; HIV = human immunodeficiency virus; STIs=sexually transmitted infections; CIN = Cervical intraepithelial neoplasia

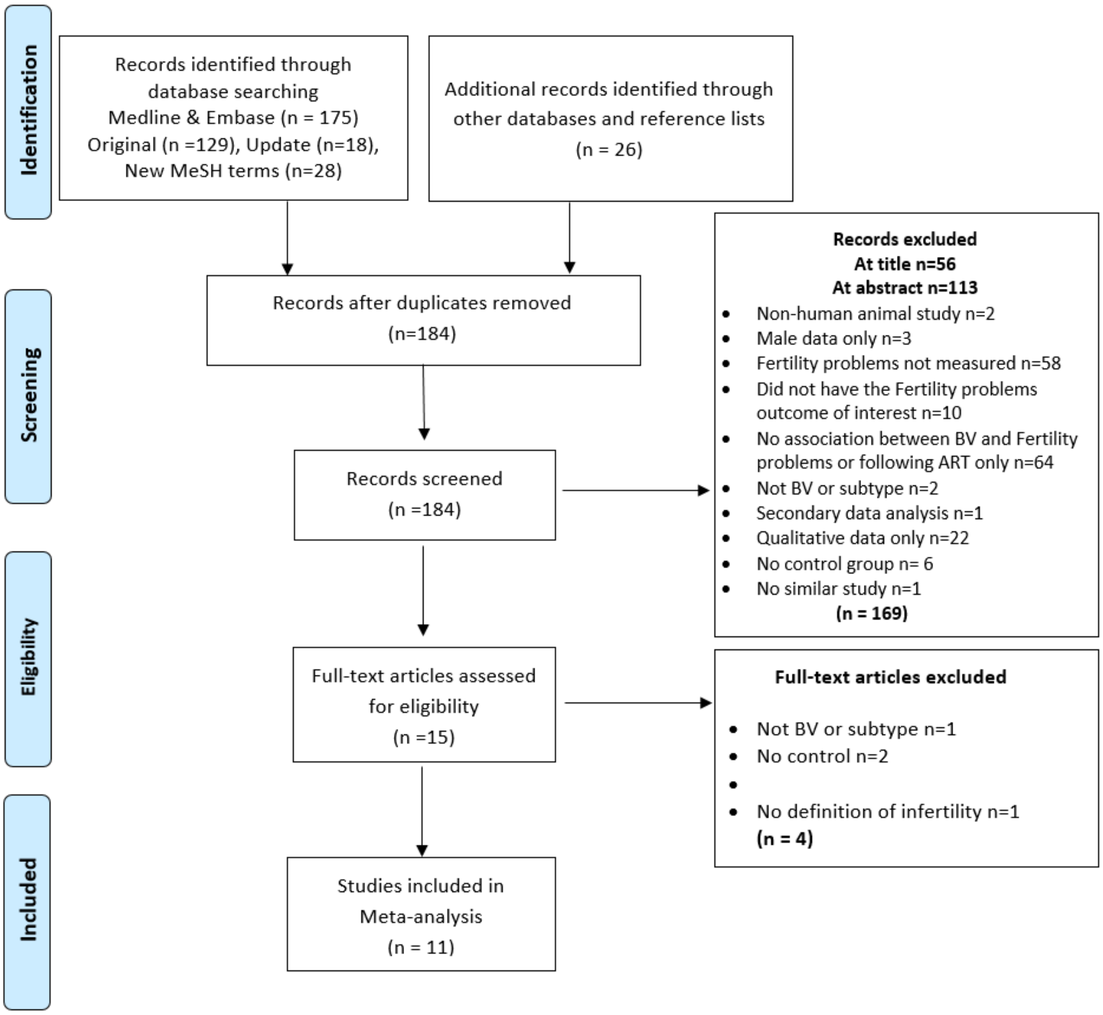


*Figure 15.* PRISMA Flow Diagram for BV. Figure shows the exclusions of articles at the different stages and the reasons for exclusion. Records identified through datbase searching of Medline and Embase includes original search, an update from the time of original search and a search using new MeSH terms. BV = Bacterial Vaginosis

Table 17.

Sample Characteristics Reported in the Eleven Included Studies

| Study | Location | Sample (n) | N | N | | Age [a] Women | |
|---|---|---|---|---|---|---|---|
| **Case-control studies** | | | **Infertile [b]** | **Fertile (control)** | | **Infertile** | **Fertile** |
| Aboul Enien, 2005 | Egypt | 60 women | 40 | 20 | Mean (SD) | NR | NR |
| Adamson, 2011 | India | 897 women | 113 | 784 | Mean (SD) | 24.0 (3.4) | 26.1 (3.0) |
| Almanza, 2011 | Cuba | 189 women | 89 | 100 | Mean | 30.4 | 24.3 |
| Dhont, 2010 | Rwanda | 571 women | 307 | 264 | Median (IQR) | 30 (27–35) | 27 (24–31) |
| Dhont, 2011 | Rwanda | 396 women | 177 | 219 | Median (IQR) | 32 (28-37) | 28 (25-32) |
| Durugbo, 2015 | Nigeria | 356 women | 178 | 178 | Mean (SD) | 28 (5) | NR |
| | | | | | <20 | 0 | 6 (3.4) |
| | | | | | 20-24 | 20 (11.2) | 20 (11.2) |
| | | | | | 25-29 | 77 (43.3) | 66 (37.1) |
| | | | | | 30-35 | 60 (33.7) | 60 (33.7) |
| | | | | | >35 | 21 (11.8) | 26 (14.6) |
| Kildea, 2000 | Australia (Indigenous Women) | 342 women | 241 | 101 | Mean (CI) | 30.4 (95% CI, 29.7-31.1) | |
| Mania-Pramanik, 2009 | India | 214 women | 112 | 102 | Mean (SD) | In BV+ women 27.7 (5.2) | |
| Morgan, 1997 | UK | 1578 women | 199 | 1379 | | NR | NR |
| Salah, 2013 | Egypt | 1256 women | 874 | 382 | Mean (SD) | 27.1 (2.2) | 25.8 (3.1) |
| Tomusiak, 2013 | Poland | 161 women | 101 | 60 | Range | 20-40 | |

*Note*. [a] Age for women at the beginning of the study; [b] Unable to become pregnant after 1 or 2 years of unprotected intercourse, a specific diagnosis e.g. idiopathic, female factor; NR = not reported; SD = Standard deviation; IQR = inter-quartile range

Table 18.

Characteristics of the Design of the Eleven Included Studies

| Study | Study design | Recruitment and data collection | Study period | BV self-report or lab test | Fertility Problems outcome measure (duration) | Control |
|---|---|---|---|---|---|---|
| Aboul Enien, 2005 | Case-control | Hospital based | | Gram staining for the presence of BV using Nugent's scoring system | Diagnosed idiopathic infertility | Fertile women |
| Adamson, 2011 | Case-control | Hospital based | 2005-2006 | Gram staining for the presence of BV using Nugent's scoring system | Primary infertility: married (or partnered) for more than two years, sexually active, not using modern contraception, and without children | Sexually active, not using modern contraception fertile women (not explicitly stated that they have a child, but only that they are fertile) |
| Almanza, 2011 | Case-control | Hospital based | 2009 | Bacteriological culture techniques | Diagnosed tubal obstruction | Currently pregnant women about to deliver |
| Dhont, 2010 | Case-control | Hospital based | 2007-2009 | Gram staining for the presence of BV using Nugent's scoring system and Amsel criteria | Infertility: having regular unprotected intercourse for 1 year or more without conception with at least one regular partner, and included both primary and secondary infertility. TFI subcategory | Non-pregnant women recently delivered (within past 6 to 18 months) |
| Dhont, 2011 | Case-control | Hospital based | 2007-2009 | Gram staining for the presence of BV using Nugent's scoring system and Amsel criteria | Secondary infertility: having regular unprotected intercourse for one year or more with at least one regular partner without conception in women who conceived at least once before | Non-pregnant women recently delivered (between 6 and 18 months ago) |
| Durugbo, 2015 | Case-control | Hospital based | 2014 | Visual assessment of discharge, then pH test, then 'whiff test' then microscopic examination ('fourth Amsel criteria') | TFI previously diagnosed by hysterosalpingography | Fertile women attending the family planning clinic |
| Kildea, 2000 | Cross-sectional | Medical records | 1996 | Culture or microscopy | Primary infertility: never given birth to a live child despite 36 months of unprotected sexual intercourse. Secondary infertility: given birth to one or more live children in the past but now unable to become pregnant after 36 months of unprotected intercourse | Women who had been able to conceive within 36 months of unprotected intercourse |
| Mania-Pramanik, 2009 | Case-control | Hospital based | NR | Gram staining for the presence of BV using Nugent's scoring system | Women who did not conceive within two years of marriage but were trying to conceive | Currently pregnant antenatal cases (first trimester, 2-3 months) |
| Morgan, 1997 | Case-control | Clinic based | | Gram staining for the presence of BV using Nugent's scoring system | Women attending at a specialist infertility clinic (trying to conceive for at least one year) | Currently pregnant (antenatal clinic) |
| Salah, 2013 | Case-control | Hospital based | 2009-2011 | Gram staining for the presence of BV using Spiegel's criteria | Women diagnosed with female factor infertility | Attending family planning |
| Tomusiak, 2013 | Case-control | Hospital/clinic based | NR | Gram staining for the presence of BV confirmed based on pH, Nugent score and quantitative culture results | Women in the infertile group had been treated for infertility for at least one year. Anatomical, hormonal abnormalities, endometriosis and abnormal sperm parameters ruled out | Women who had no history of fertility problems and at least one child |

*Note*. BV = Bacterial vaginosis; TFI = tubal factor infertility; NR = not reported

Table 19.

Quality Ratings for the Eleven Included Studies on the Basis of an Adapted Newcastle-Ottawa Quality Assessment Scale

| Study | Quality Criterion | | | | | | Overall rating [g] |
|---|---|---|---|---|---|---|---|
| | **Adequacy of infertility (exposed) measure[a] Max 2 points** | **Adequacy of control (non-exposed), definition and selection [b] Max 2 points** | **Comparability of control [c] Max 2 points** | **Confounders adequately assessed Max 2 points [d]** | **Adequacy of outcome BV measure [e] Max 1 point** | **Loss to follow-up [f] Max 1 point** | |
| Aboul Enien, 2005 | 2 | 1 | 0 | 0 | 1 | NA | Average |
| Adamson, 2011 | 1 | 1 | 1 | 2 | 1 | NA | Average |
| Almanza, 2011 | 1 | 1 | 1 | 1 | 1 | NA | Average |
| Dhont, 2010 | 1 | 2 | 1 | 2 | 1 | NA | High |
| Dhont, 2011 | 1 | 2 | 1 | 1 | 1 | NA | Average |
| Durugbo, 2015 | 2 | 1 | 2 | 1 | 0 | NA | Average |
| Kildea, 2000 | 2 | 2 | 2 | 2 | 0 | NA | High |
| Mania-Pramanik,2009 | 1 | 2 | 0 | 0 | 1 | NA | Average |
| Morgan, 1997 | 0 | 2 | 0 | 0 | 1 | NA | Low |
| Salah, 2013 | 2 | 1 | 0 | 0 | 1 | NA | Average |
| Tomusiak, 2013 | 2 | 1 | 0 | 0 | 1 | NA | Average |

*Note*. NA= not applicable; **[a]** Infertility was adequately assessed when independent validation of (e.g. laboratory testing and/or hospital/medical records) and it was representative of the cohort i.e. drawn from the same population (up to 2 points); **[b]** Controls were adequately assessed when selection was comparable to cases, and infertility was excluded properly in the control population (up to 2 points); **[c]** Comparability of controls was achieved if exposed/non-exposed were matched or adjustment during analysis conducted. One point for *STIs* and one point for any other confounder (up to 2 points); **[d]** Confounders were adequately assessed if they were obtained from records or a blind interview, and one point was given if the same method was used for both groups (up to 2 points); **[e]**Fertility problems outcome was adequately assessed if independent or blind assessment was stated in the paper, or confirmation of the outcome by reference to secure records (medical records, etc.) (up to 1 point); **[f]** Point given if same rate for both groups and <20% loss to follow up reported; **[g]** The overall quality rating was low (0 to 3 points), average (4 to 6 points), or high (7 to 10 points).

Table 20.

Number and Percentage of Infertile Women in BV and No-BV Groups in the Included Studies (k=11)

| Studies included | Number of women (%) | |
|---|---|---|
| | BV | No-BV |
| All studies<br>k=11 | 846 of 1421 (59.5) | 1443 of 4597 (31.4) |
| Exclusively TFI (subgroup)<br>k=2 | 114 of 159 (71.7) | 153 of 386 (39.6) |
| Not only TFI (subgroup)<br>k=9 | 732 of 1262 (58.0) | 1290 of 4211 (30.6) |

*Note*. BV = bacterial vaginosis; TFI = tubal factor infertility

### Results of Meta-analyses

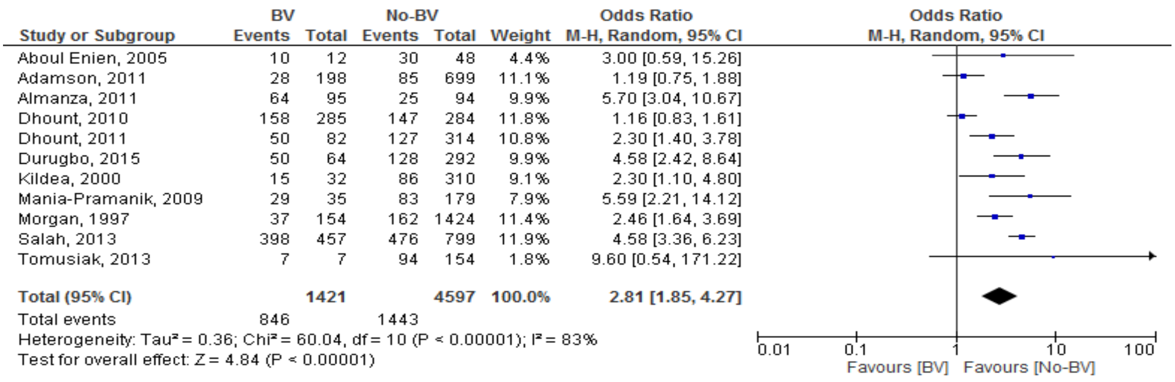


*Figure 16*. Odds ratio for women who are infertile in the BV and No-BV groups

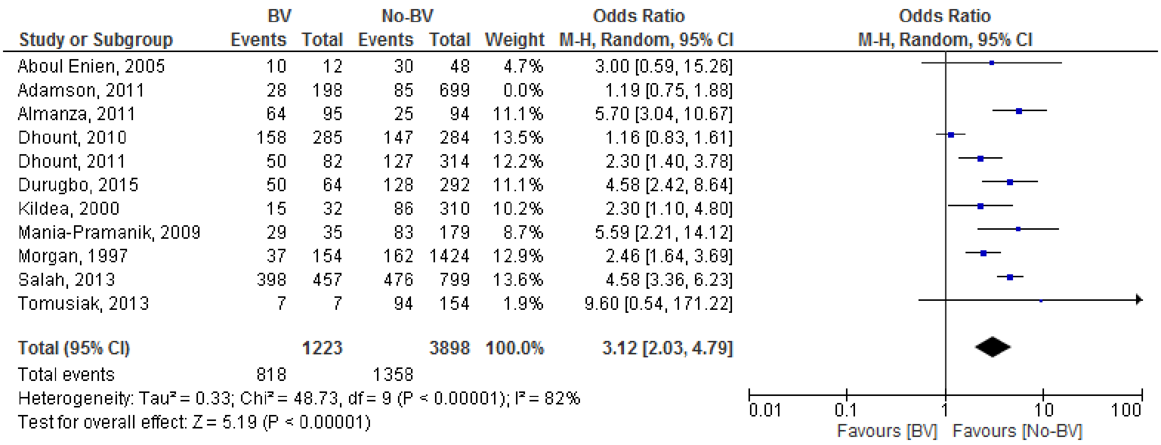


*Figure 17*. Sensitivity analysis by outcome (removed one study reporting childlessness and compared only studies retorting inability to become pregnant) for the comparison ‘Odds ratio for women who are infertile in the BV and No-BV groups’

| Study or Subgroup | BV Events | BV Total | No-BV Events | No-BV Total | Weight | Odds Ratio M-H, Random, 95% CI |
|---|---|---|---|---|---|---|
| **1.3.1 Only tubal infertility** | | | | | | |
| Almanza, 2011 | 64 | 95 | 25 | 94 | 9.9% | 5.70 [3.04, 10.67] |
| Durugbo, 2015 | 50 | 64 | 128 | 292 | 9.9% | 4.58 [2.42, 8.64] |
| **Subtotal (95% CI)** | | **159** | | **386** | **19.8%** | **5.11 [3.27, 7.99]** |
| Total events | 114 | | 153 | | | |
| Heterogeneity: Tau² = 0.00; Chi² = 0.23, df = 1 (P = 0.63); I² = 0% | | | | | | |
| Test for overall effect: Z = 7.16 (P < 0.00001) | | | | | | |
| **1.3.2 Not only tubal infertility** | | | | | | |
| Aboul Enien, 2005 | 10 | 12 | 30 | 48 | 4.4% | 3.00 [0.59, 15.26] |
| Adamson, 2011 | 28 | 198 | 85 | 699 | 11.1% | 1.19 [0.75, 1.88] |
| Dhount, 2010 | 158 | 285 | 147 | 284 | 11.8% | 1.16 [0.83, 1.61] |
| Dhount, 2011 | 50 | 82 | 127 | 314 | 10.8% | 2.30 [1.40, 3.78] |
| Kildea, 2000 | 15 | 32 | 86 | 310 | 9.1% | 2.30 [1.10, 4.80] |
| Mania-Pramanik, 2009 | 29 | 35 | 83 | 179 | 7.9% | 5.59 [2.21, 14.12] |
| Morgan, 1997 | 37 | 154 | 162 | 1424 | 11.4% | 2.46 [1.64, 3.69] |
| Salah, 2013 | 398 | 457 | 476 | 799 | 11.9% | 4.58 [3.36, 6.23] |
| Tomusiak, 2013 | 7 | 7 | 94 | 154 | 1.8% | 9.60 [0.54, 171.22] |
| **Subtotal (95% CI)** | | **1262** | | **4211** | **80.2%** | **2.42 [1.53, 3.84]** |
| Total events | 732 | | 1290 | | | |
| Heterogeneity: Tau² = 0.34; Chi² = 48.65, df = 8 (P < 0.00001); I² = 84% | | | | | | |
| Test for overall effect: Z = 3.77 (P = 0.0002) | | | | | | |
| **Total (95% CI)** | | **1421** | | **4597** | **100.0%** | **2.81 [1.85, 4.27]** |
| Total events | 846 | | 1443 | | | |
| Heterogeneity: Tau² = 0.36; Chi² = 60.04, df = 10 (P < 0.00001); I² = 83% | | | | | | |
| Test for overall effect: Z = 4.84 (P < 0.00001) | | | | | | |
| Test for subgroup differences: Chi² = 5.22, df = 1 (P = 0.02), I² = 80.9% | | | | | | |

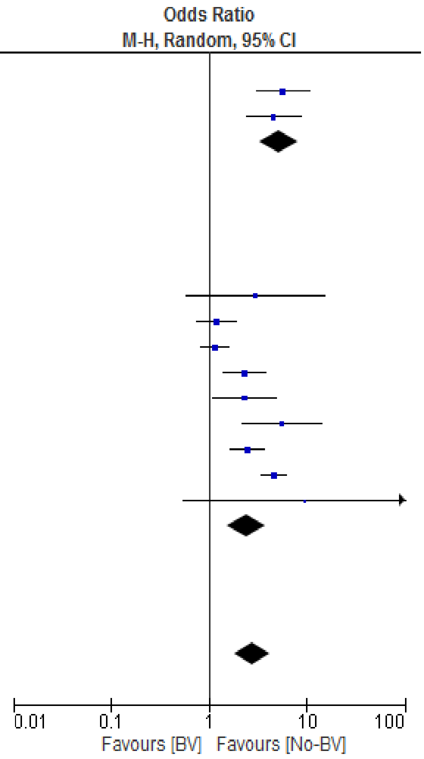


*Figure 18*. Subgroup analysis by outcome (with studies that are exclusively TFI, and studies that are not only TFI) for the comparison 'Odds ratio for women who are infertile in the BV and No-BV groups'

### Publication bias assessment.

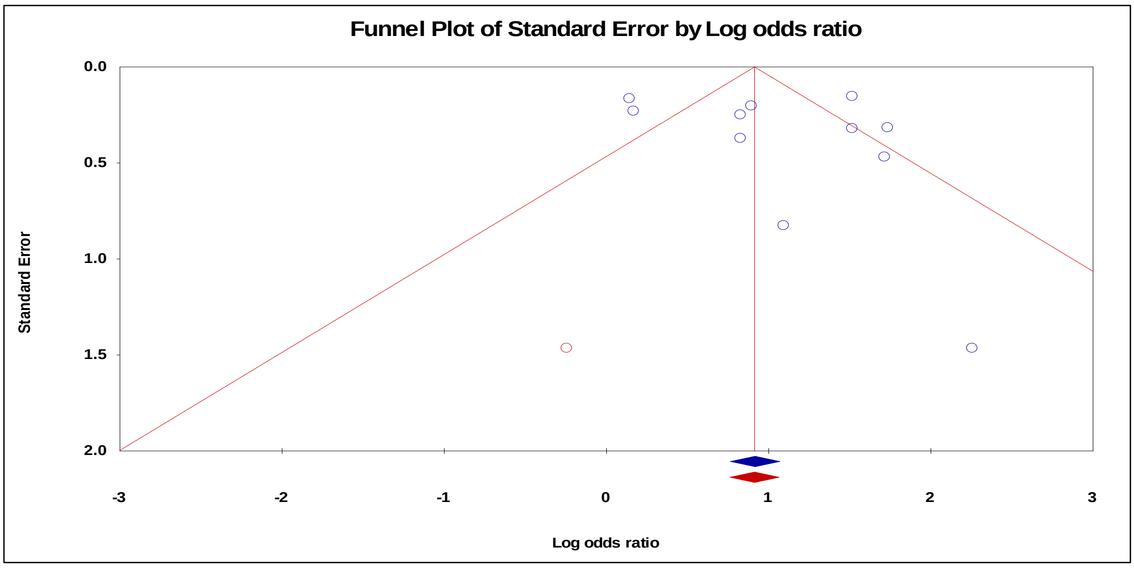


*Figure 19*. Funnel plot with trim and fill procedure to impute 'missing' studies (missing studies in red) for the 'odds ratio for women who are infertile in the BV and No-BV groups'

### STIs and Sexual History Reported in the Included Studies

Data were not available to enable a subgroup analysis of women with STIs and those without. Only a summary of percentages of women with STIs in the BV and No-BV groups was possible, see Table 3.5.7.

Table 21.

Percentage of Women with Comorbid STIs or a History of STIs in Infertile Versus Fertile Women in Eight of the Eleven Included Studies (k=8)

<table>
<tr><th>Study</th><th>Type of infection</th><th colspan="4">Percentage of STIs</th></tr>
<tr><th></th><th></th><th colspan="2">Infertile (%)</th><th colspan="2">Fertile (control) (%)</th></tr>
<tr><td>Adamson, 2011</td><td>HSV</td><td colspan="2">22/113 (19.5)</td><td colspan="2">81/784 (10.3)</td></tr>
<tr><td>Almanza, 2011</td><td>Chlamydia</td><td colspan="2">41/89 (46)</td><td colspan="2">2/100 (2)</td></tr>
<tr><td></td><td>Mycoplasma hominis</td><td colspan="2">15/89 (16.9)</td><td colspan="2">10/100 (10)</td></tr>
<tr><td></td><td>Ureaplasma urealyticum</td><td colspan="2">38/89 (42.7)</td><td colspan="2">2/100 (2)</td></tr>
<tr><td>Dhont, 2010</td><td>HIV</td><td colspan="2">98/312 (32)</td><td colspan="2">39/283 (14)</td></tr>
<tr><td></td><td>HSV</td><td colspan="2">180/312 (59)</td><td colspan="2">115/283 (41)</td></tr>
<tr><td></td><td>Chlamydia</td><td colspan="2">57/312 (19)</td><td colspan="2">44/283 (16)</td></tr>
<tr><td>Dhont, 2011</td><td>HIV</td><td colspan="2">74/177 (42)</td><td colspan="2">35/219 (16)</td></tr>
<tr><td></td><td>HSV</td><td colspan="2">121/177 (70)</td><td colspan="2">99/219 (45)</td></tr>
<tr><td></td><td>Chlamydia</td><td colspan="2">31/177 (18)</td><td colspan="2">33/219 (15)</td></tr>
<tr><td>Kildea, 2000</td><td>Chlamydia</td><td colspan="2">36/101 (36)</td><td colspan="2">68/241 (28)</td></tr>
<tr><td></td><td>Gonorrhoeae</td><td colspan="2">42/101 (42)</td><td colspan="2">51/241 (21)</td></tr>
<tr><td></td><td>Trichomonas vaginalis</td><td colspan="2">64/101 (63)</td><td colspan="2">95/241 (39)</td></tr>
<tr><td>Tomusiak, 2013</td><td>Chlamydia</td><td colspan="2">0/101 (0)</td><td colspan="2">2/60 (3)</td></tr>
<tr><td></td><td>Mycoplasma hominis</td><td colspan="2">4/101 (4)</td><td colspan="2">0/60 (0)</td></tr>
<tr><td></td><td>Ureaplasma urealyticum</td><td colspan="2">9/101 (9)</td><td colspan="2">5/60 (8)</td></tr>
<tr><td>Durugbo, 2015</td><td>History of STIs</td><td colspan="2">64/178 (36)</td><td colspan="2">35/178 (19.7)</td></tr>
<tr><td></td><td></td><td>Infertile/BV</td><td>Infertile/no-BV</td><td>Fertile/BV</td><td>Fertile/no-BV</td></tr>
<tr><td></td><td></td><td>38/50 (74)</td><td>26/128 (20.3)</td><td>11/14 (79)</td><td>24/164 (14.6)</td></tr>
<tr><td>Mania-Pramanik,2009</td><td>Chlamydia and HPV</td><td>5/29 (17.2)</td><td>NR</td><td>0/6 (0)</td><td>NR</td></tr>
</table>

*Note:* HSV = herpes simplex virus; HIV = human immune deficiency virus; HPV = human papilloma virus; STIs = sexually transmitted infections; NR = not reported

### Consanguinity

Plausible Mechanisms to explain how consanguinity could be associated with fertility problems

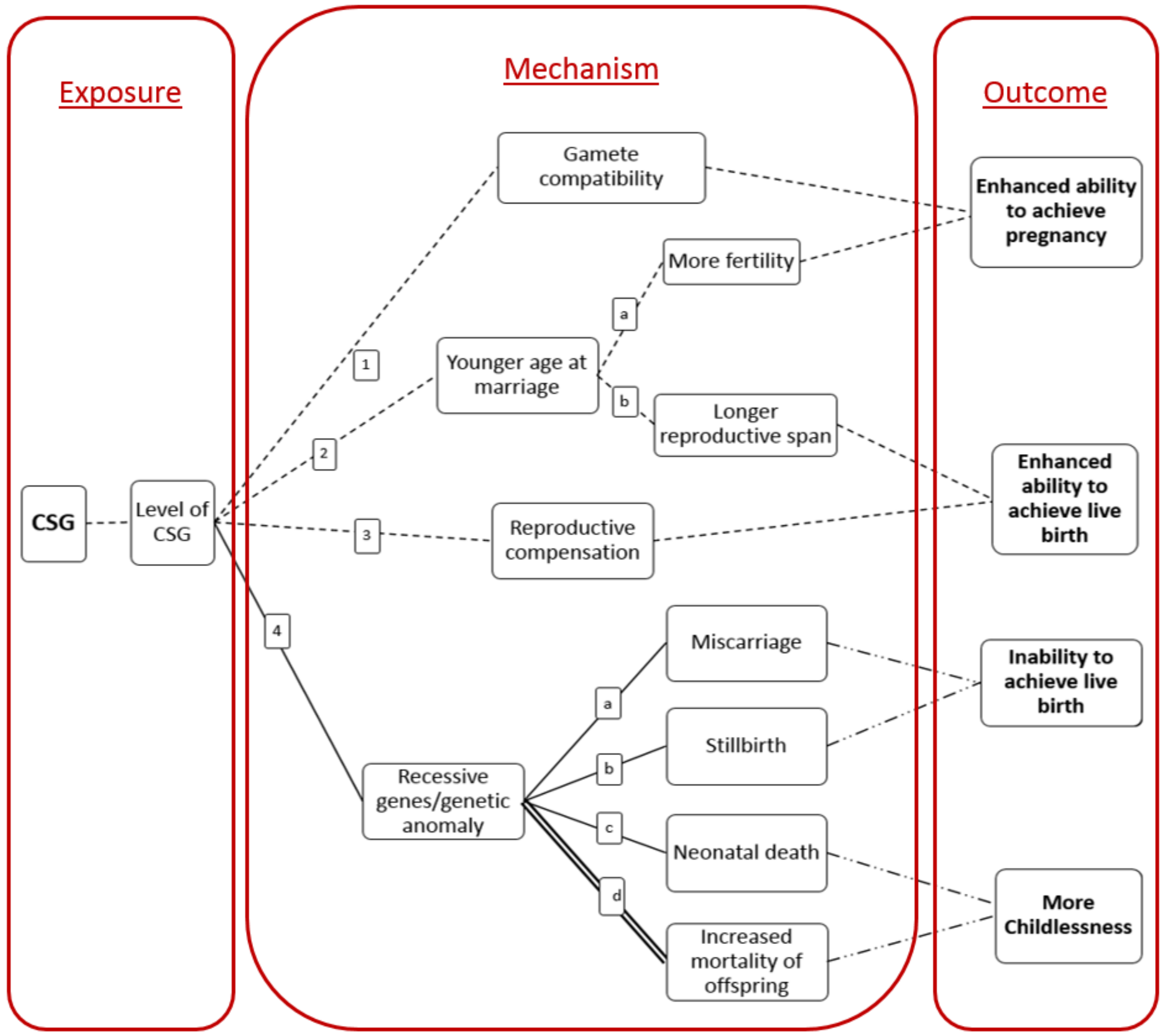


*Figure 20*. Proposed pathways for the impact of Consanguinity (CSG) on fertility. Solid line = Recent evidence (primary studies); Double solid line = meta-analytic evidence; Dashed line =Proposed pathway/historic evidence; Dashed-Dotted line = Well established

Table 22.

Summary of reproductive health consequences of CSG reported in the literature

| **Reproductive outcome** | **Effect of CSG** | **Statistics reported (where available)** | **Review** |
|---|---|---|---|
| | **Positive effect** | | |
| Live birth rate | Statistically significant in the first cousin only but not in other categories of CSG | First cousins had 0.26 more children | Bittles et al., 2002 |
| | Higher live birth rate in first cousin marriages compared to non-CSG marriages | Mean live births range in first cousins (2.26-7.48) in non-CSG (2.14-5.83) | Hussain and Bittles, 2004 |
| | **Negative effect** | | |
| Mortality of offspring | More mortality in progeny of first cousins compared to non-CSG progeny | Meta-analysis showed significant mean excess mortality of 3.5% in the CSG progeny ($r2 = 0.70$; $P < 0.00001$) | Bittles & Black, 2010 |
| Mortality and morbidity of offspring | Higher rates of infant morbidity and mortality in offspring of CSG couples than non-CSG couples where reported | Range of infant morbidity 1.34-42% in CSG and 0.81-25% in non-CSG, mortality 0.95-8.6% in the CSG and 0.63-5.3% in non-CSG | Bhasin & Shampa, 2012 |
| Recessive genes in offspring | Probability of inheriting recessive gene increases with the increase in the proximity of the relationship between parents | NR | Hamamy, 2012 |

*Note*. NR = not reported; CSG = consanguinity/consanguineous

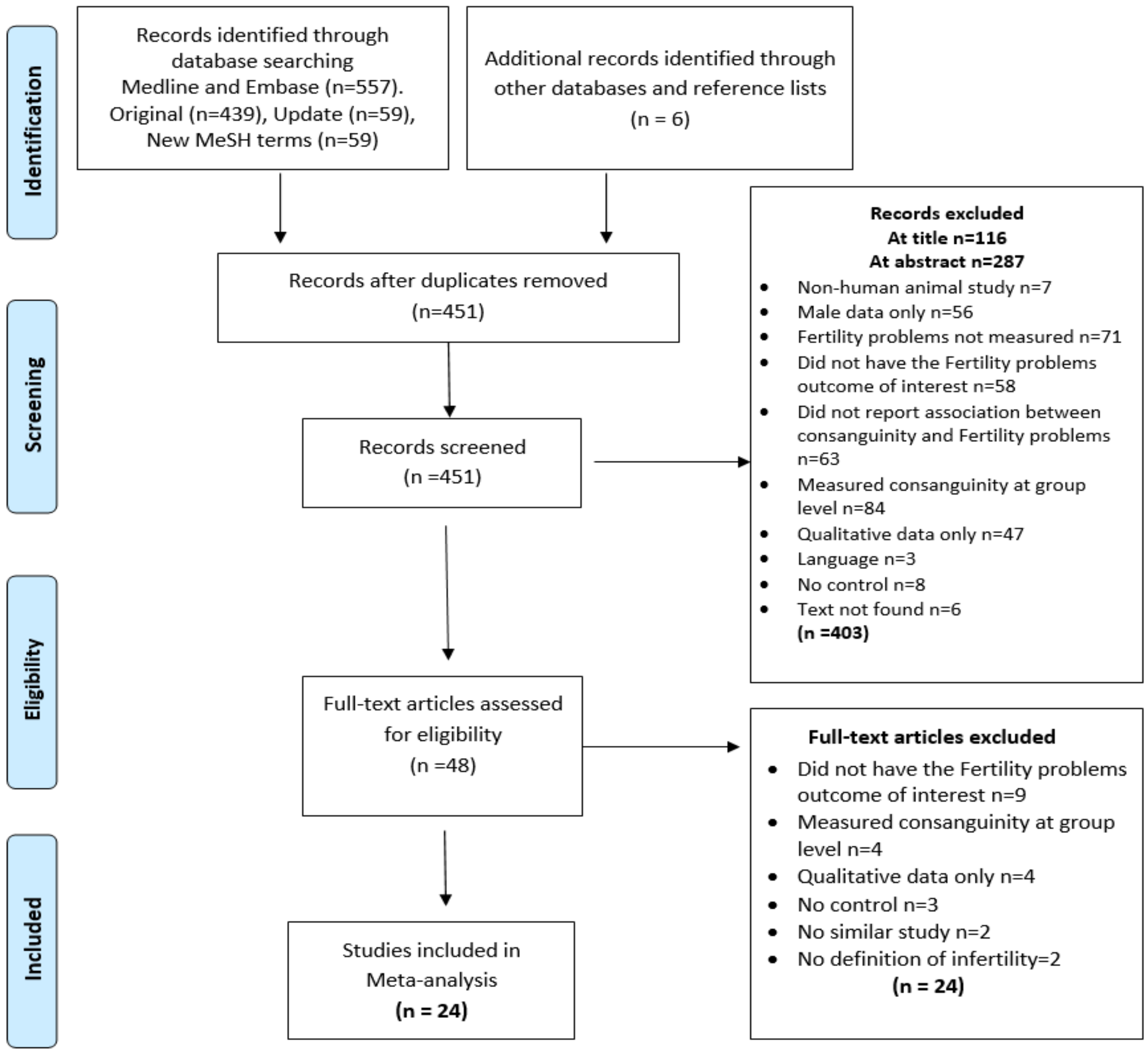


*Figure 21*. PRISMA Flow Diagram for consanguinity. Figure shows the exclusion of articles at the different stages and the reasons for exclusion. Records identified through datbase searching of Medline and Embase includes original search, an update from the time of original search and a search using new MeSH terms.

Table 23.

Sample characteristics of the 24 included Studies

| Study | Location | Sample (n) | CSG (n) | Non- CSG (n) | Mean age at marriage | | | |
|---|---|---|---|---|---|---|---|---|
| | | | | | Women | | Men | |
| | | | | | CSG | Non-CSG | CSG | Non-CSG |
| Edo, 1985 | Spain | 965 couples | 272 | 693 | 25.74 | 26.02 | 28.9 | 29.2 |
| Hann, 1984 | Karnataka State in South India | 1885 women | 722 | 1163 | NR | | | |
| Tanaka, 1977 | Fukuoka, Japan | 1450 couples | 346 | 1104 | NR | | | |
| Yamaguchi, 1975 | Fukuoka, Japan | 4026 couples | 2173 | 1853 | NR | | | |
| Bittles, 1993 | Punjabi Provence of Pakistan | 9520 women | 4784 | 4736 | 18.97 | 19.74 | 23.81 | 25.97 |
| Rao, 1979 | Southern India District of Tamil Nadu | 15, 926 women | 6379 | 9547 | NR | | | |
| Shami, 1990 | Punjabi Provence of Pakistan | 3329 women | 2227 | 1102 | 18.95 | 19.93 | 23.7 | 26 |
| Al-Kandari 2007 | Kuwait | 7315 women | 4009 | 3306 | NR | | | |
| Bener 2006 | Qatar | 1515 women | 818 | 687 | NR | | | |
| Blanco 2006 | Leon, Spain | 2670 women | 474 | 2196 | 25.63 | 26.70 | 28.81 | 30.39 |
| Ciceklioglu 2013 | Bayrakli, suburb of Izmir, Turkey | 170 women | 85 | 85 | NR | | | |
| Devi 1981 | Karnataka, South India | 3254 women | 920 | 2301 | NR | | | |

| Study | Location | Sample (n) | CSG (n) | Non- CSG (n) | Mean age at marriage | | | |
|---|---|---|---|---|---|---|---|---|
| | | | | | Women | | Men | |
| | | | | | CSG | Non-CSG | CSG | Non-CSG |
| Fuster 2003 | Los Nogales, Galicia, Spain | 1581 | 132 | 1449 | 24.58 | | | |
| Khlat 1988 | Beirut, Lebanon | 2801 | 705 | 2096 | NR | | | |
| Khoury 2000 | Jordan | 1867 | 947 | 920 | 24.6 | 25.8 | | |
| Luna 1990 | La Alpujarra, Andalusia, Spain | 647 | 75 | 572 | NR | | | |
| Abdulrazzaq 1997 | Alain & Dubai, UAE | 2033 | 1026 | 100 | NR | | | |
| Al Husain 1996 | Riyadh, KSA | 2001 couples | 1022 | 979 | NR | | | |
| Asha 1981 | South India | 377 women | 156 | 211 | NR | | | |
| Gharyeb 2014 | Yatta, Palestine | 500 women | 305 | 195 | NR | | | |
| Islam 2013 | Oman | 2037 women | 1052 | 985 | NR | | | |
| Saha 1990 | Khartoum, Sudan | 926 women | 586 | 340 | NR | | | |
| Verma 1992 | Pondicherry, India | 1000 women | 308 | 692 | NR | | | |
| Yuksel 2009 | Malatya, Turkey | 409 women | 116 | 293 | NR | | | |

*Note*: CSG = consanguineous/consanguinity; [a] Mean age for women at the beginning of the study; NR= data not reported

Table 24.

Characteristics of the design of the 24 included studies

| Study | Study design | Data collection | Study period | CSG measure | Fertility Problems outcome measure (duration) |
|---|---|---|---|---|---|
| Edo, 1985 | Retrospective cohort | Extracted from parish records and civil registries | 1900-1974 | 1st and 2nd degree cousins | Childless marriages at the end of reproductive life (age 45) |
| Hann, 1984 | Cross-sectional | Household interviews | Not reported | 1st degree cousin and Uncle-niece | Primary sterility defined as never having conceived in (1) women who have completed reproduction (over 40, menopausal or widowed) or (2) after 10 years without contraception in women of reproductive age |
| Tanaka, 1977 | Retrospective cohort | Household interviews in 2 rural villages and cross-checked with records | Not reported | CSG between spouse, between husband's parents and between wife's parents | Infertility defined as never been pregnant after living with husband for more than 5 years |
| Yamaguchi, 1975 | Retrospective cohort | Household interviews in rural villages and cross-checked with records | Not reported | CSG between spouse, between husband's parents and between wife's parents | Sterility defined as no pregnancy after more than 5 years of marriage |
| Bittles, 1993 (not in 71) | Cross-sectional | Household & hospital interviews in 11 cities | 1979-1985 | Mixed, double 1st cousin, 1st cousin, double second cousin, second cousin, | Time to first delivery from start of marriage in years |
| Rao, 1979 | Cross-sectional | Household interviews in In 14 rural and urban districts | 1969-1975 | Mixed, uncle-niece, first cousin, beyond first cousin | Primary sterility defined as a married woman who has not had a live-born baby after consummation of marriage and unprotected sexual activity (duration in 5 year intervals) |
| Shami, 1990 | Cross-sectional | from general hospital and labour wards, as well as door-to-door interviews | 1980-1983 | Mixed, double first cousin, first cousin, first cousin once removed, second Cousin. | Time to first birth from start of marriage in years |
| Al-Kandari 2007 | Cross-sectional | Questionnaires filled by women attending 10 different PHC | 2002 | Double cousin, first cousin, second cousin, third cousin | Number of births per women |

| Study | Study design | Data collection | Study period | CSG measure | Fertility Problems outcome measure (duration) |
|---|---|---|---|---|---|
| Bener 2006 | Cross-sectional | Questionnaires filled by face-to-face interviews from 10 health centres mostly visited and women’s hospital | 2004 | Double cousin, first cousin, first cousin once removed, second cousin, less than second cousin | Number of pregnancies and live births |
| Blanco 2006 | Cross-sectional | La Cabrera parish registers | 1880-1959 | Up to third degree | Live births |
| Ciceklioglu 2013 | Cross-sectional | Community based in-person interviews from 3 neighbourhoods in Bayraklu | 2009 | First and second degree cousins | Number of pregnancies and deliveries |
| Devi 1981 | Cross-sectional | 17 hospitals, maternity homes and health centres from records or interviews by staff | 1971 | Beyond second cousin, second cousin, first cousin, uncle-niece | Mean number of live born |
| Fuster 2003 | Cross-sectional | Biodemographic data from parish and Lugo bisphoric records | 1871-1977 | Uncle-niece, first cousin, first cousin once removed, second cousin, second cousin once removed, third cousin | Mean birth |
| Khlat 1988 | Cross-sectional | 2752 household were interviewed | 1983-1984 | First cousin and more distant than first cousin | Mean number of pregnancies, live births |
| Khoury 2000 | Cross-sectional | Community based, 7200 households | 1980 | Double first cousins, first cousin 1,2.3 and 4, first cousins once removed, from the family | Number of pregnancies |
| Luna 1990 | Cross-sectional | Community based. 8 villages in an isolated mountain population | NR | Level of CSG NR | Average number of pregnancies, live births |
| Abdulrazzaq 1997 | Cross-sectional | Antenatal, postnatal and immunization centres based interviews and questionnaires | 1994-1995 | Double first degree, first cousin, first cousin once removed, second cousin, less than second cousin | Number of abortions and still births |

| Study | Study design | Data collection | Study period | CSG measure | Fertility Problems outcome measure (duration) |
|---|---|---|---|---|---|
| Al Husain 1996 | Cross-sectional | PHC and antenatal care clinic interviews | 1993 | Double first cousin, first cousin, second cousin, more distant relative | Abortion, still birth and neonatal death |
| Asha 1981 | Prospective cohort study | NR | NR | Uncle-niece, first cousin, first cousin once removed, second cousin, second cousin once removed, third cousin | Abortion (termination =<28 weeks), still birth (born with no heart beat), neonatal death (within first 28 days of life) |
| Gharyeb 2014 | Cross-sectional | Community based, personally interviewed by structured questionnaires | NR | First degree, second degree, third degree | Abortion (at or before 28 weeks), still births |
| Islam 2013 | Cross-sectional | ONHS data, 2013 household were interviewed | 2000 | First cousin; father's side, first cousin; mother's side, other; second cousin and beyond | Mean number of pregnancies, live births, number of miscarriage, number of still birth |
| Saha 1990 | Cross-sectional | ANC clinic in the OBGYN department, faculty of Medicine, U of K | NR | First cousins; mother's brother & sister, father's brother & sister, Other type of CSG marriages | Abortion, Still birth, neonatal deaths |
| Verma 1992 | Cross-sectional | Interview in maternity ward in JIPMER hospital | 1978 | First cousin; MBD or FSD, uncle-niece, other; beyond first cousin | Neonatal death |
| Yuksel 2009 | Cross-sectional | Household interviews, face to face questionnaires | NR | First cousin, others; half first cousin and second degree cousin, distant CSG marriages | Spontaneous abortions, still births |

*Note*. CSG = consanguineous/consanguinity; NR = not reported; PHC = primary health care

Table 25.

Quality ratings for the 24 included studies on the basis of an adapted Newcastle-Ottawa quality assessment scale

| Study | Quality Criterion | | | | | | Overall rating [g] |
|---|---|---|---|---|---|---|---|
| | Adequacy of CSG(exposed) measure[a] Max 2 points | Adequacy of control (non-exposed), definition and selection [b] Max 2 points | Comparability of control [c] Max 2 points | Confounders adequately assessed Max 2 points [d] | Adequacy of outcome Fertility Problems measure [e] Max 1 point | None response rate or loss to follow-up [f] Max 1 point | |
| Edo, 1985 | 1 | 2 | 1 | 1 | 1 | NR | Average |
| Hann, 1984 | 2 | 2 | 0 | 0 | 0 | NA | Average |
| Tanaka, 1977 | 2 | 2 | 1 | 0 | 1 | NR | Average |
| Yamaguchi, 1975 | 2 | 2 | 1 | 0 | 1 | NR | Average |
| Bittles, 1993 | 2 | 2 | 1 | 1 | 0 | NA | Average |
| Rao, 1979 | 2 | 2 | 1 | 1 | 0 | NA | Average |
| Shami, 1990 | 2 | 2 | 0 | 1 | 0 | NA | Average |
| Al-Kandari 2007 | 1 | 2 | 1 | 1 | 0 | NA | Average |
| Bener 2006 | 1 | 2 | 0 | 1 | 0 | NA | Low |
| Blanco 2006 | 1 | 2 | 1 | 2 | 1 | NA | High |
| Cicekliogl 2013 | 2 | 2 | 1 | 2 | 1 | NA | High |
| Devi 1981 | 2 | 2 | 0 | 2 | 1 | NA | High |
| Fuster 2003 | 1 | 2 | 0 | 2 | 1 | NA | Average |
| Khlat 1988 | 1 | 2 | 1 | 2 | 0 | NA | Average |
| Khoury 2000 | 2 | 1 | 1 | 2 | 0 | NA | Average |

| Study | Quality Criterion: Adequacy of CSG(exposed) measure[a] Max 2 points | Adequacy of control (non-exposed), definition and selection [b] Max 2 points | Comparability of control [c] Max 2 points | Confounders adequately assessed Max 2 points [d] | Adequacy of outcome Fertility Problems measure [e] Max 1 point | None response rate or loss to follow-up [f] Max 1 point | Overall rating [g] |
|---|---|---|---|---|---|---|---|
| Luna 1990 | 1 | 1 | 2 | 0 | 0 | NA | Low |
| Abdulrazzaq 1997 | 1 | 2 | 1 | 2 | 0 | NA | Average |
| Al Husain 1996 | 2 | 2 | 1 | 1 | 0 | NA | Average |
| Asha 1981 | 2 | 1 | 1 | 2 | 0 | NR | Average |
| Gharyeb 2014 | 1 | 2 | 1 | 1 | 0 | NA | Average |
| Islam 2013 | 1 | 2 | 0 | 2 | 0 | NA | Average |
| Saha 1990 | 1 | 2 | 1 | 2 | 0 | NA | Average |
| Verma 1992 | 2 | 2 | 1 | 2 | 0 | NA | High |
| Yuksel 2009 | 2 | 2 | 1 | 2 | 0 | NA | High |

*Note***.** CSG = consanguineous/consanguinity; NR = not reported; NA = not applicable
[a] CSG was adequately assessed when independent validation of the degree of relatedness was assessed or coefficient of CSG(F) calculated, (e.g. >1 person/record/time/process to extract information, or reference to primary record source such as medical/hospital records) and it was representative of the cohort i.e. drawn from the same population (up to 2 points); [b] Controls were adequately assessed when selection was comparable to cases, and CSG was excluded properly in the control population (up to 2 points); [c] Comparability of controls was achieved if exposed/non-exposed were matched or adjustment during analysis conducted. One point for age at marriage and one point for any other confounder (e.g. education) (up to 2 points); [d] Confounders were adequately assessed if they were obtained from records or a blind interview, and one point was given if the same method was used for both groups (up to 2 points); [e] Fertility problems outcome was adequately assessed if independent or blind assessment was stated in the paper, or confirmation of the outcome by reference to secure records (medical records, etc.) (up to 1 point); [f] Point given if same rate for both groups and <20% loss to follow up reported; [g] The overall quality rating was low (0 to 3 points), average (4 to 6 points), or high (7 to 10 points).

Table 26.

Proportion of specific outcome in CSG and non-CSG couples in the included studies, (k=24)

| **Outcome** | **CSG** | **Non-CSG** |
|---|---|---|
| **Outcome (number of studies)** | **Number (%)** | **Number (%)** |
| Never pregnant (k=3) | 92 of 3241 (2.8) | 186 of 4120 (4.5) |
| Childless (K=5) | 380 of 6651 (5.7) | 717 of 10,240 (7.0) |
| Miscarriages (k=7) | 1069 of 3372 (31.7) | 1030 of 3485 (29.6) |
| Stillbirths (k=7) | 243 of 3372 (7.2) | 211 of 3485 (6.1) |
| Neonatal death (k=7) | 151 of 2072 (7.3) | 144 of 2232 (6.5) |
| **Outcome (number of studies)** | **Mean (SD), total** | **Mean (SD), total** |
| Mean time to first birth in years (k=2) | 1.8 (24.8), 7011 | 1.6 (9.4), 2608 |
| Mean number of pregnancies (k=5) | 5.0 (3.0), 2735 | 4.6 (2.9), 4435 |
| Mean number of live births (k=7) | 3.9 (2.5), 7433 | 3.7 (2.3), 10142 |

*Note*. CSG = Consanguineous; Non-CSG = none consanguineous

### Results of Meta-analyses

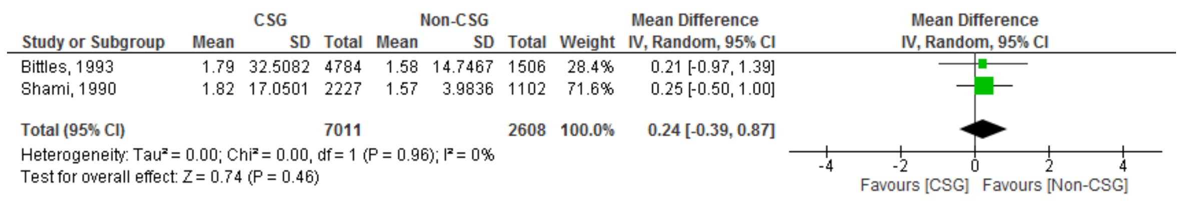


*Figure 22*. Odds ratio for 'time-to-first-birth' (in years) in the CSG and non-CSG groups

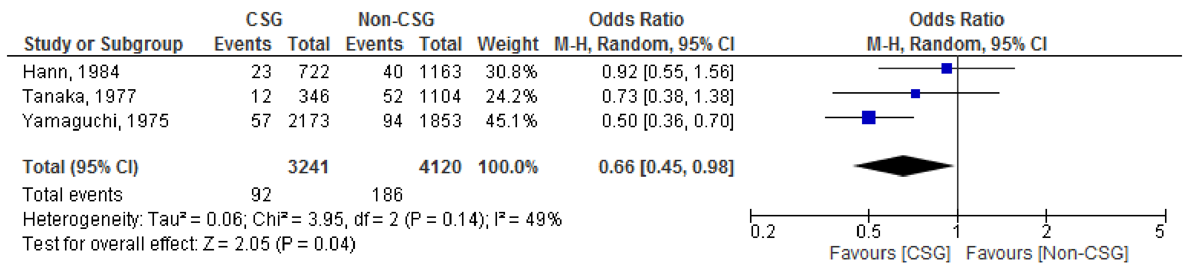


*Figure 23*. Odds ratio for proportion of couples who were 'never-pregnant' in the CSG and non-CSG groups

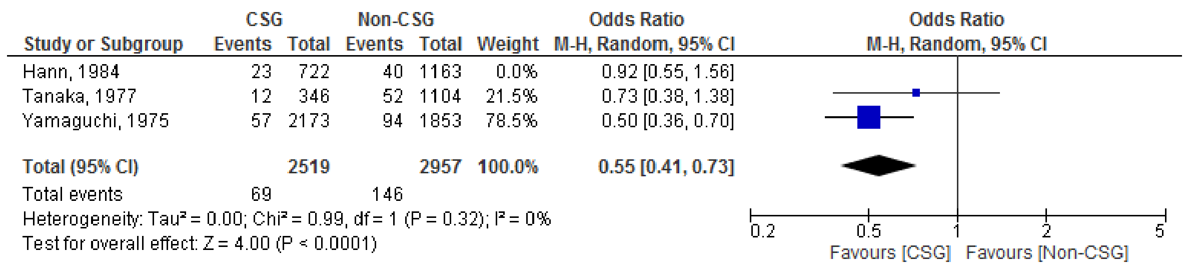


*Figure 24*. Sensitivity analysis by duration for the comparison odds ratio for proportion of couples who were 'never-pregnant' in the CSG and non-CSG groups

| Study or Subgroup | CSG Events | CSG Total | Non-CSG Events | Non-CSG Total | Weight | Odds Ratio M-H, Random, 95% CI |
|---|---|---|---|---|---|---|
| Edo, 1985 | 30 | 272 | 70 | 693 | 13.8% | 1.10 [0.70, 1.74] |
| Rao, grp 1 (5-9 yrs) | 164 | 1456 | 268 | 2353 | 26.3% | 0.99 [0.80, 1.21] |
| Rao, grp 2 (10-14 yrs) | 59 | 1367 | 157 | 2156 | 20.4% | 0.57 [0.42, 0.78] |
| Rao, grp 3 (15-19 yrs) | 49 | 1141 | 78 | 1575 | 17.5% | 0.86 [0.60, 1.24] |
| Rao, grp 4 (20 + yrs) | 78 | 2415 | 144 | 3463 | 21.9% | 0.77 [0.58, 1.02] |
| **Total (95% CI)** | | **6651** | | **10240** | **100.0%** | **0.83 [0.67, 1.03]** |
| Total events | 380 | | 717 | | | |

Heterogeneity: Tau² = 0.04; Chi² = 10.04, df = 4 (P = 0.04); I² = 60%
Test for overall effect: Z = 1.68 (P = 0.09)

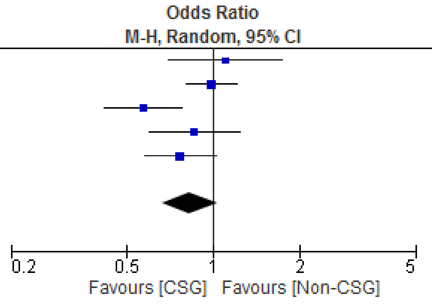


*Figure 25*. Odds ratio for proportion of ‘childless’ couples in the CSG and non-CSG groups

| Study or Subgroup | CSG Events | CSG Total | Non-CSG Events | Non-CSG Total | Weight | Odds Ratio M-H, Random, 95% CI |
|---|---|---|---|---|---|---|
| **15.2.1 >20 years** | | | | | | |
| Edo, 1985 | 30 | 272 | 70 | 693 | 13.8% | 1.10 [0.70, 1.74] |
| Rao, grp 4 (20 + yrs) | 78 | 2415 | 144 | 3463 | 21.9% | 0.77 [0.58, 1.02] |
| **Subtotal (95% CI)** | | **2687** | | **4156** | **35.8%** | **0.88 [0.63, 1.24]** |
| Total events | 108 | | 214 | | | |
| Heterogeneity: Tau² = 0.03; Chi² = 1.76, df = 1 (P = 0.18); I² = 43% | | | | | | |
| Test for overall effect: Z = 0.73 (P = 0.46) | | | | | | |
| **15.2.2 <20 years** | | | | | | |
| Rao, grp 1 (5-9 yrs) | 164 | 1456 | 268 | 2353 | 26.3% | 0.99 [0.80, 1.21] |
| Rao, grp 2 (10-14 yrs) | 59 | 1367 | 157 | 2156 | 20.4% | 0.57 [0.42, 0.78] |
| Rao, grp 3 (15-19 yrs) | 49 | 1141 | 78 | 1575 | 17.5% | 0.86 [0.60, 1.24] |
| **Subtotal (95% CI)** | | **3964** | | **6084** | **64.2%** | **0.80 [0.57, 1.12]** |
| Total events | 272 | | 503 | | | |
| Heterogeneity: Tau² = 0.07; Chi² = 8.27, df = 2 (P = 0.02); I² = 76% | | | | | | |
| Test for overall effect: Z = 1.32 (P = 0.19) | | | | | | |
| **Total (95% CI)** | | **6651** | | **10240** | **100.0%** | **0.83 [0.67, 1.03]** |
| Total events | 380 | | 717 | | | |

Heterogeneity: Tau² = 0.04; Chi² = 10.04, df = 4 (P = 0.04); I² = 60%
Test for overall effect: Z = 1.68 (P = 0.09)
Test for subgroup differences: Chi² = 0.17, df = 1 (P = 0.68), I² = 0%

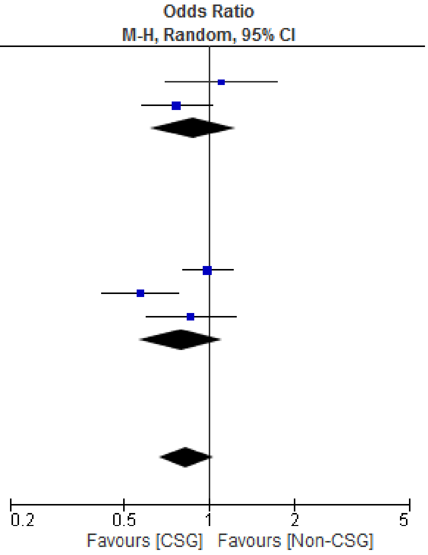


*Figure 26*. Subgroup analysis by duration (>20 years vs <20 years) for the comparison odds ratio for the proportion of ‘childless’ couples in the CSG and non-CSG groups

| Study or Subgroup | CSG Mean | CSG SD | CSG Total | Non-CSG Mean | Non-CSG SD | Non-CSG Total | Weight | Mean Difference IV, Random, 95% CI |
|---|---|---|---|---|---|---|---|---|
| Bener, 2006 | 6.33 | 2.87 | 818 | 5.88 | 3.1 | 697 | 24.4% | 0.45 [0.15, 0.75] |
| Ciceklioglu, 2013 | 2.77 | 1.86 | 85 | 2.72 | 1.79 | 85 | 15.7% | 0.05 [-0.50, 0.60] |
| Islam, 2013 | 5.5 | 4 | 1052 | 5.4 | 3.9 | 985 | 22.8% | 0.10 [-0.24, 0.44] |
| Khlat, 1988 | 5.69 | 3.475 | 705 | 4.88 | 3.41 | 2096 | 24.7% | 0.81 [0.51, 1.11] |
| Luna and Fuster, 1990 | 4.83 | 2.85 | 75 | 4.32 | 2.44 | 572 | 12.4% | 0.51 [-0.17, 1.19] |
| **Total (95% CI)** | | | **2735** | | | **4435** | **100.0%** | **0.40 [0.10, 0.71]** |

Heterogeneity: Tau² = 0.07; Chi² = 11.78, df = 4 (P = 0.02); I² = 66%
Test for overall effect: Z = 2.60 (P = 0.009)

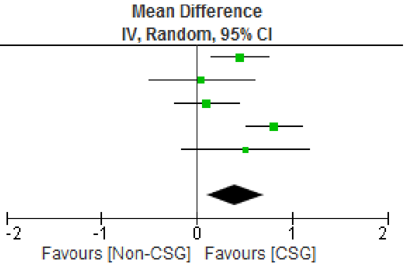


*Figure 27*. Mean difference for ‘Number of pregnancies’ in the CSG and non-CSG groups

| Study or Subgroup | CSG Mean | CSG SD | CSG Total | Non-CSG Mean | Non-CSG SD | Non-CSG Total | Weight | Mean Difference IV, Random, 95% CI |
|---|---|---|---|---|---|---|---|---|
| Al-Kandari, 2007 | 3.79 | 2.62 | 4009 | 3.41 | 2.28 | 3306 | 19.8% | 0.38 [0.27, 0.49] |
| Bener, 2006 | 5.35 | 2.67 | 818 | 5.03 | 2.73 | 697 | 14.7% | 0.32 [0.05, 0.59] |
| Blanco, 2006 | 4.35 | 2.77 | 474 | 3.89 | 2.84 | 2196 | 14.6% | 0.46 [0.18, 0.74] |
| Ciceklioglu, 2013 | 2.14 | 1.28 | 85 | 2.09 | 1.2 | 85 | 11.6% | 0.05 [-0.32, 0.42] |
| Devi, 1981 | 2.54 | 1.592 | 920 | 2.56 | 1.54 | 2301 | 19.6% | -0.02 [-0.14, 0.10] |
| Islam, 2013 | 4.8 | 3.6 | 1052 | 4.7 | 3.5 | 985 | 13.6% | 0.10 [-0.21, 0.41] |
| Luna and Fuster, 1990 | 4.43 | 2.71 | 75 | 3.91 | 2.2 | 572 | 6.2% | 0.52 [-0.12, 1.16] |
| **Total (95% CI)** | | | **7433** | | | **10142** | **100.0%** | **0.24 [0.05, 0.43]** |

Heterogeneity: Tau² = 0.04; Chi² = 28.63, df = 6 (P < 0.0001); I² = 79%
Test for overall effect: Z = 2.46 (P = 0.01)

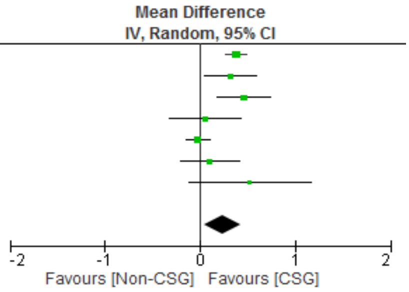


*Figure 28*. Mean Difference for 'Number of live births" in the CSG and non-CSG groups

| Study or Subgroup | CSG Mean | CSG SD | CSG Total | Non-CSG Mean | Non-CSG SD | Non-CSG Total | Weight | Mean Difference IV, Random, 95% CI |
|---|---|---|---|---|---|---|---|---|
| Al-Kandari, 2007 | 3.79 | 2.62 | 4009 | 3.41 | 2.28 | 3306 | 46.5% | 0.38 [0.27, 0.49] |
| Bener, 2006 | 5.35 | 2.67 | 818 | 5.03 | 2.73 | 697 | 14.9% | 0.32 [0.05, 0.59] |
| Blanco, 2006 | 4.35 | 2.77 | 474 | 3.89 | 2.84 | 2196 | 14.6% | 0.46 [0.18, 0.74] |
| Ciceklioglu, 2013 | 2.14 | 1.28 | 85 | 2.09 | 1.2 | 85 | 8.7% | 0.05 [-0.32, 0.42] |
| Devi, 1981 | 2.54 | 1.592 | 920 | 2.56 | 1.54 | 2301 | 0.0% | -0.02 [-0.14, 0.10] |
| Islam, 2013 | 4.8 | 3.6 | 1052 | 4.7 | 3.5 | 985 | 12.1% | 0.10 [-0.21, 0.41] |
| Luna and Fuster, 1990 | 4.43 | 2.71 | 75 | 3.91 | 2.2 | 572 | 3.2% | 0.52 [-0.12, 1.16] |
| **Total (95% CI)** | | | **6513** | | | **7841** | **100.0%** | **0.32 [0.21, 0.44]** |

Heterogeneity: Tau² = 0.00; Chi² = 6.18, df = 5 (P = 0.29); I² = 19%
Test for overall effect: Z = 5.47 (P < 0.00001)

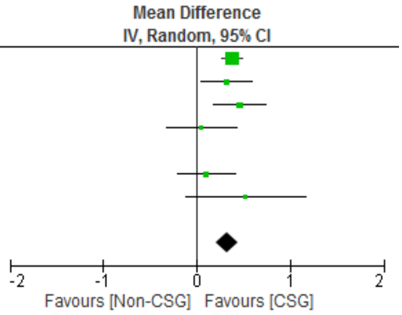


*Figure 29*. Sensitivity analysis for the Mean Difference for 'Number of live births" in the CSG and non-CSG groups (without DEVI, 1981)

| Study or Subgroup | CSG Events | CSG Total | Non-CSG Events | Non-CSG Total | Weight | Odds Ratio M-H, Random, 95% CI |
|---|---|---|---|---|---|---|
| Abdulrazzaq, 1997 | 320 | 1026 | 300 | 1007 | 27.7% | 1.07 [0.88, 1.29] |
| Al Husain, 1997 | 301 | 1022 | 250 | 979 | 26.9% | 1.22 [1.00, 1.48] |
| Asha, 1981 | 29 | 156 | 37 | 221 | 7.7% | 1.14 [0.66, 1.94] |
| Islam, 2013 | 385 | 1052 | 384 | 985 | 28.8% | 0.90 [0.76, 1.08] |
| Yuksel, 2009 | 34 | 116 | 59 | 293 | 8.9% | 1.64 [1.01, 2.69] |
| **Total (95% CI)** | | **3372** | | **3485** | **100.0%** | **1.10 [0.93, 1.30]** |
| Total events | 1069 | | 1030 | | | |

Heterogeneity: Tau² = 0.02; Chi² = 8.06, df = 4 (P = 0.09); I² = 50%
Test for overall effect: Z = 1.15 (P = 0.25)

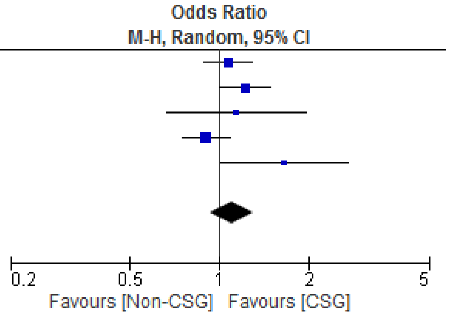


*Figure 30*. Odds ratio for proportion of 'miscarriages' in the CSG and non-CSG groups

| Study or Subgroup | CSG Events | CSG Total | Non-CSG Events | Non-CSG Total | Weight | Odds Ratio M-H, Random, 95% CI |
|---|---|---|---|---|---|---|
| Abdulrazzaq, 1997 | 75 | 1026 | 57 | 1007 | 29.5% | 1.31 [0.92, 1.88] |
| Al Husain, 1997 | 64 | 1022 | 50 | 979 | 26.1% | 1.24 [0.85, 1.82] |
| Asha, 1981 | 7 | 156 | 18 | 221 | 5.1% | 0.53 [0.22, 1.30] |
| Islam, 2013 | 79 | 1052 | 52 | 985 | 28.7% | 1.46 [1.01, 2.09] |
| Yuksel, 2009 | 18 | 116 | 34 | 293 | 10.6% | 1.40 [0.76, 2.59] |
| **Total (95% CI)** | | **3372** | | **3485** | **100.0%** | **1.28 [1.04, 1.57]** |
| Total events | 243 | | 211 | | | |

Heterogeneity: Tau² = 0.00; Chi² = 4.32, df = 4 (P = 0.36); I² = 7%
Test for overall effect: Z = 2.37 (P = 0.02)

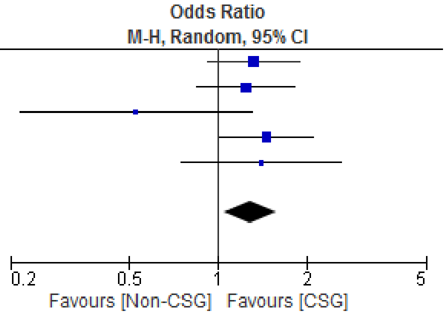


*Figure 31*. Odds ratio for proportion of 'stillbirths' in the CSG and non-CSG groups

| Study or Subgroup | CSG Events | CSG Total | Non-CSG Events | Non-CSG Total | Weight | Odds Ratio M-H, Random, 95% CI |
|---|---|---|---|---|---|---|
| Al Husain, 1997 | 66 | 1022 | 32 | 979 | 33.9% | 2.04 [1.33, 3.15] |
| Asha, 1981 | 19 | 156 | 17 | 221 | 13.3% | 1.66 [0.84, 3.32] |
| Saha, 1990 | 17 | 586 | 8 | 340 | 8.7% | 1.24 [0.53, 2.90] |
| Verma, 1992 | 49 | 308 | 87 | 692 | 44.0% | 1.32 [0.90, 1.92] |
| **Total (95% CI)** | | **2072** | | **2232** | **100.0%** | **1.57 [1.22, 2.02]** |
| Total events | 151 | | 144 | | | |

Heterogeneity: Tau² = 0.00; Chi² = 2.59, df = 3 (P = 0.46); I² = 0%
Test for overall effect: Z = 3.51 (P = 0.0005)

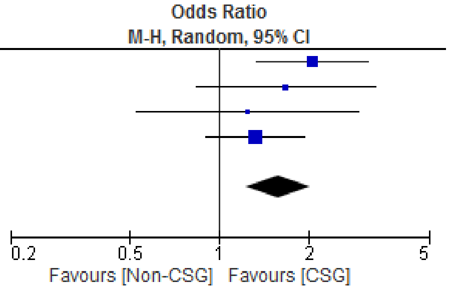


*Figure 32*. Odds ratio for proportion of ‘neonatal deaths’ in the CSG and non-CSG groups

**Publication bias assessment.**

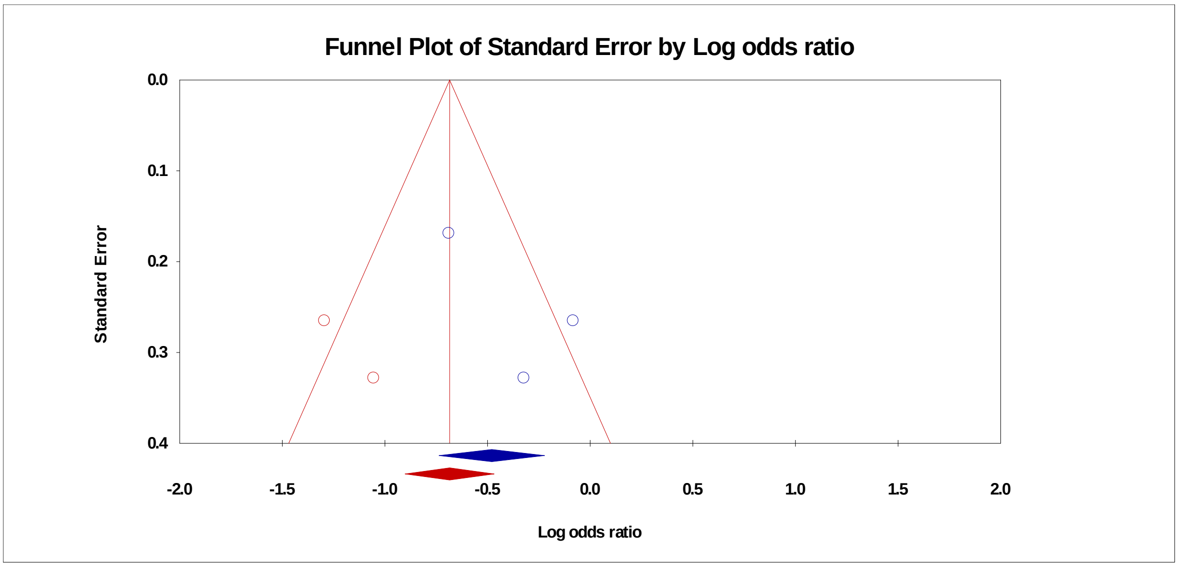


*Figure 33*. Funnel plot with trim and fill procedure to impute ‘missing’ studies (missing studies in red) for the percentage ‘never-pregnant’ analysis

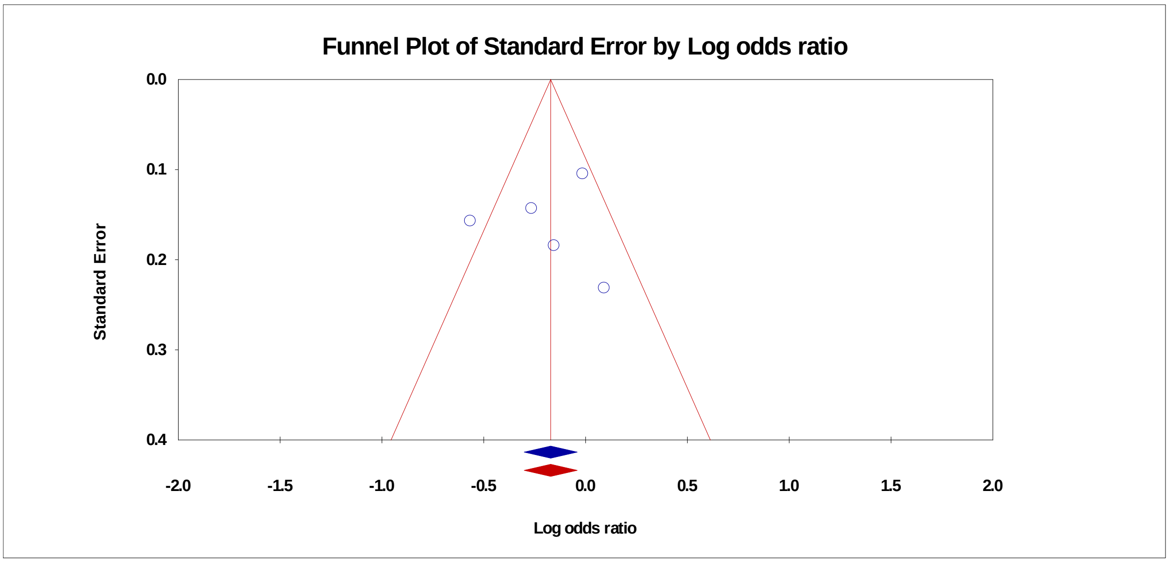


*Figure 34*. Funnel plot with trim and fill procedure to impute 'missing' studies (missing studies in red) for the percentage 'childless' analysis

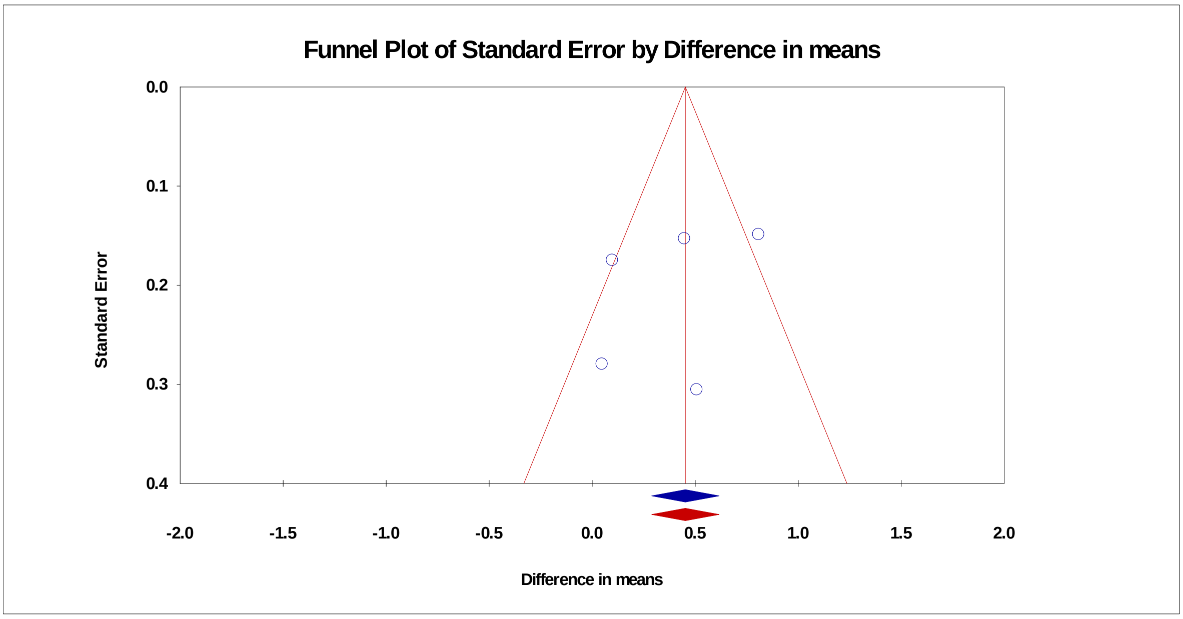


*Figure 35*. Funnel plot with trim and fill procedure to impute 'missing' studies (missing studies in red) for the Mean Difference for 'Number of Pregnancies' analysis

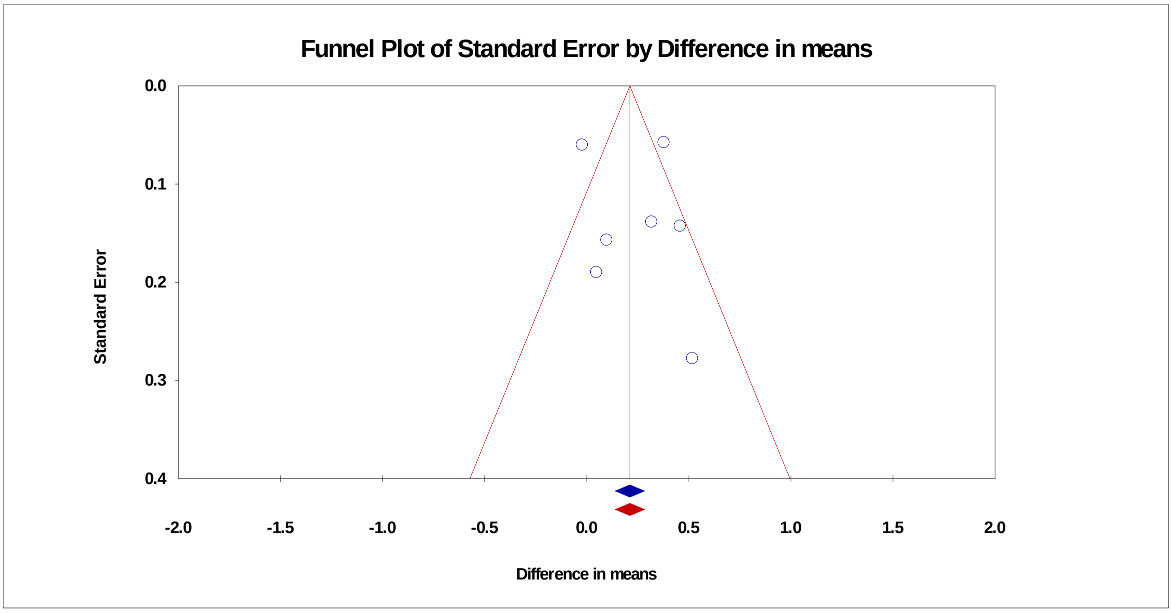


*Figure 36*. Funnel plot with trim and fill procedure to impute ‘missing’ studies (missing studies in red) for the Mean Difference for ‘Number of live births’ analysis

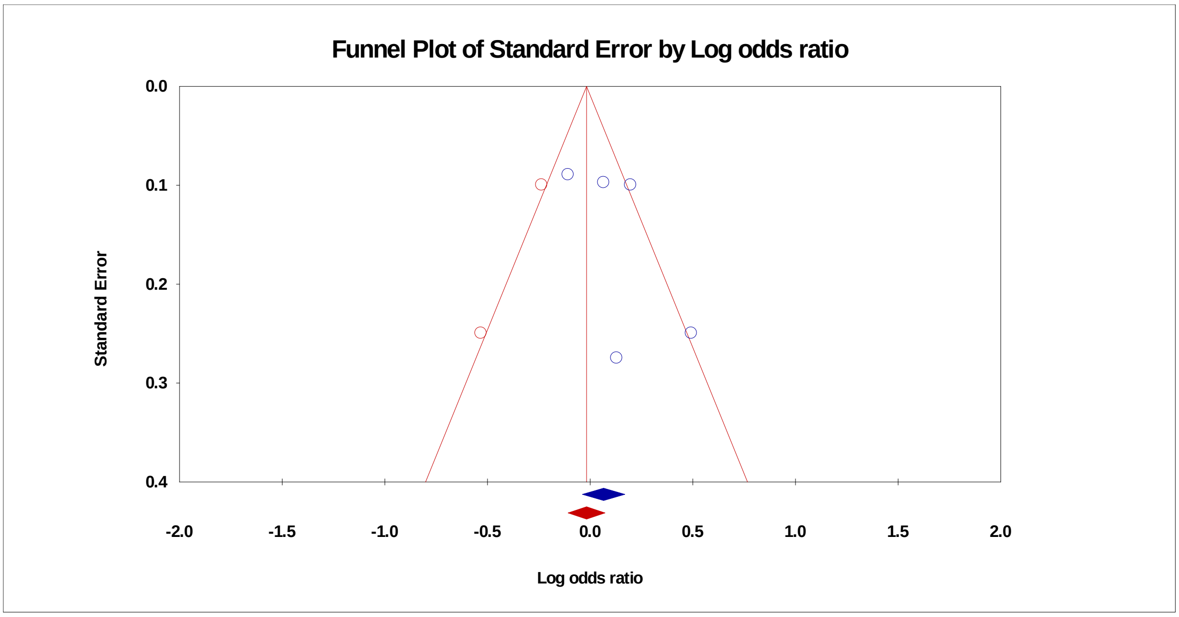


*Figure 37*. Funnel plot with trim and fill procedure to impute ‘missing’ studies (missing studies in red) for the percentage ‘Miscarriage’ analysis

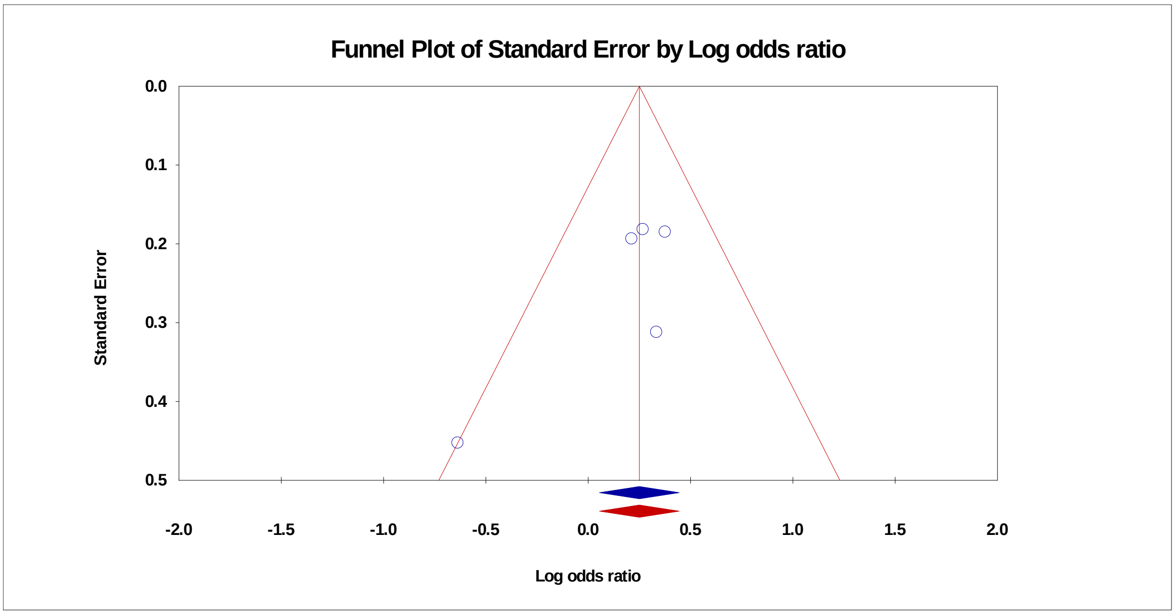


*Figure 38*. Funnel plot with trim and fill procedure to impute ‘missing’ studies (missing studies in red) for the percentage ‘Stillbirth’ analysis

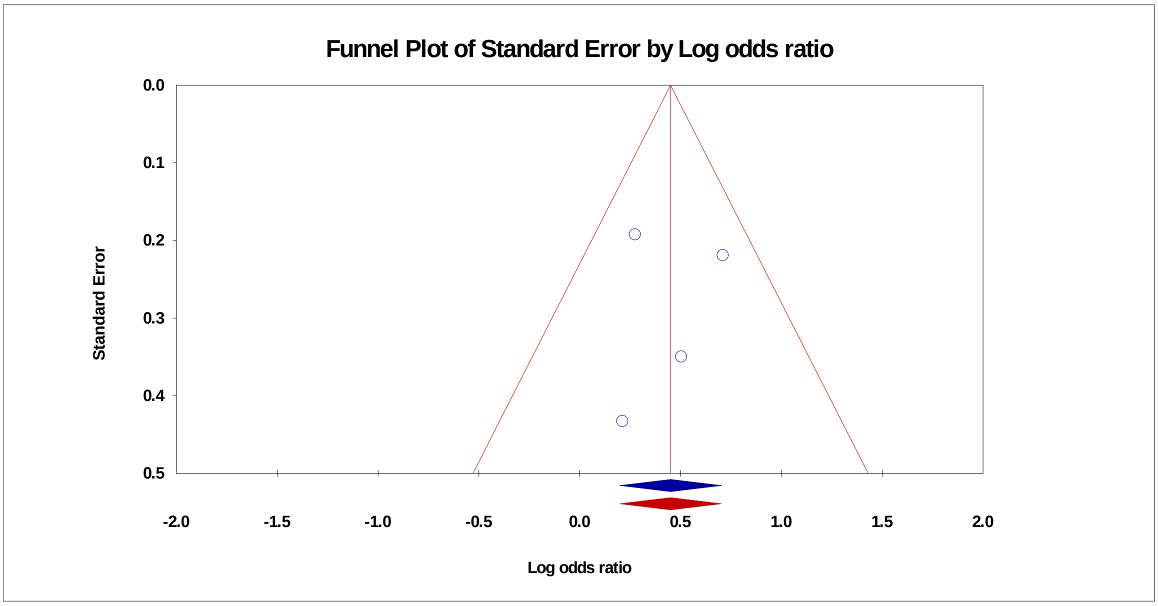


*Figure 39*. Funnel plot with trim and fill procedure to impute ‘missing’ studies (missing studies in red) for the percentage ‘Neonatal Death’ analysis

### Female Genital Mutilation/Cutting

Table 27.

WHO classification of FGM/C

| Type | Definition |
|---|---|
| Type I | Clitoridectomy; partial or total removal of the clitoris (a small sensitive and erectile part of the female genitals) or, in rare cases, only the prepuce (the fold of skin surrounding the clitoris) |
| Type II | Excision; partial or total removal of the clitoris and labia minora with or without removal or the labia majora (the labia are “the lips” that surround the vagina) |
| Type III | Infibulation; narrowing of the vaginal opening through the creation of a covering seal. The seal is formed by cutting and repositioning the labia minora or majora with or without removal of the clitoris |
| Type IV | Other; all other harmful procedures to the genital for non-medical reasons e.g. pricking, piercing, incision, scraping and cauterising the genital area |

*Note*. WHO = World Health Organization; FGM/C = Female Genital Mutilation/Cutting

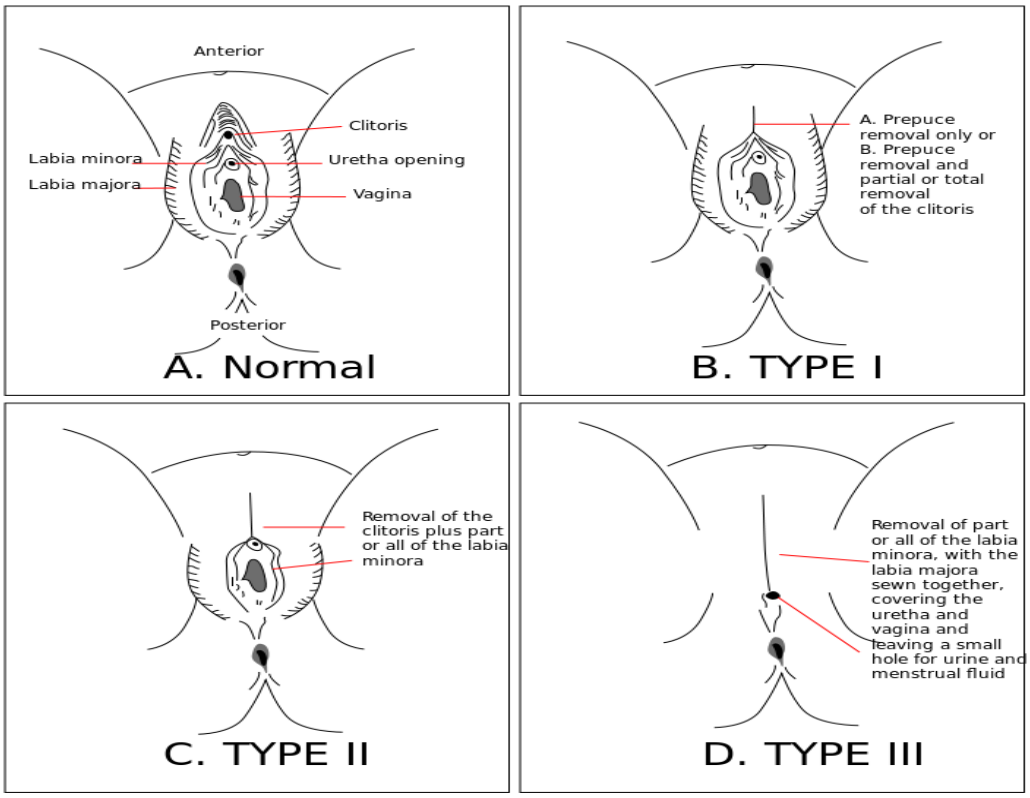


*Figure 40*. FGM/C procedure as classified by WHO. Figure from https://en.wikipedia.org/wiki/File:FGC_Types.svg Copyright by WHO. Reprinted with permission

Plausible Mechanisms to explain how FGM/C could be associated with fertility problems

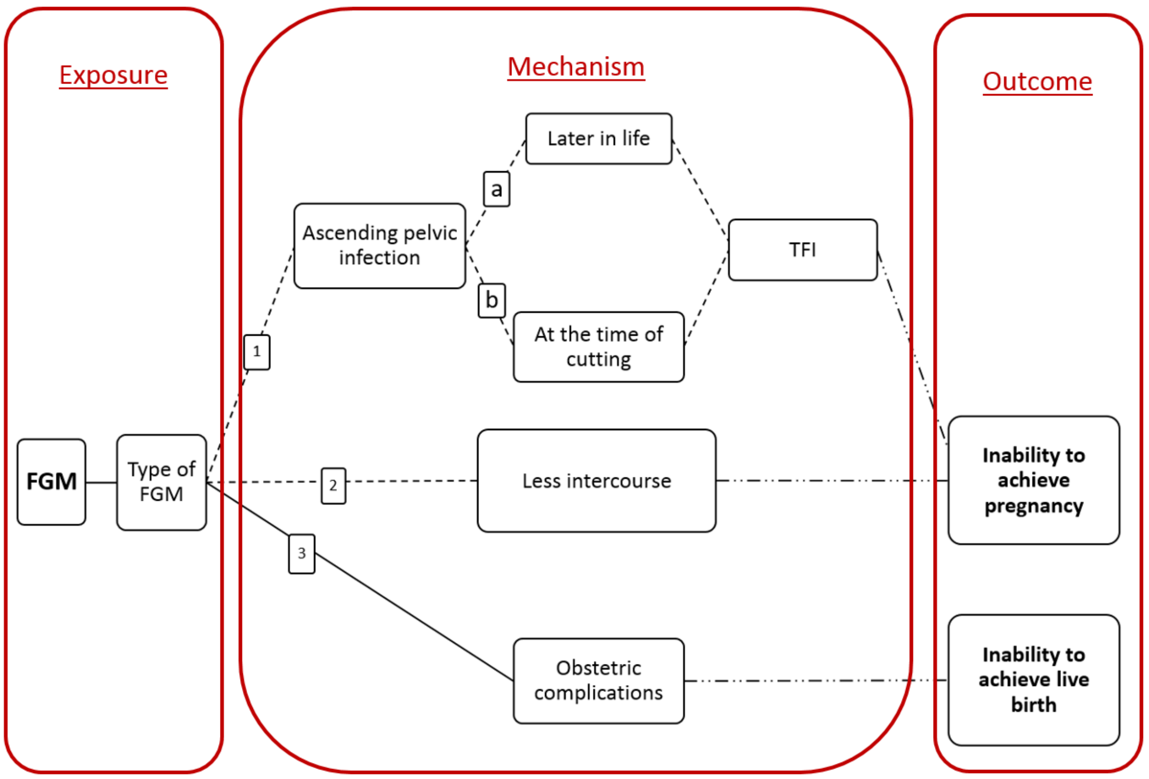


*Figure 41*. Proposed pathways for the impact of FGM/C on fertility. Solid line = Recent evidence; Dashed line = Proposd pathway/historic evidence; Dashed-Dotted line = Well established; FGM/C = female genital mutilation/cutting; TFI = tubal factor infertility

Table 28.

Summary of Reproductive Health Consequences of FGM/C Reported in the Literature

| Reproductive outcome | Effect of FGM/C | Statistics reported (where available) | | Review |
|---|---|---|---|---|
| | | **Percentage** | **Odds ratio** | |
| Short-term | | | | |
| | Traumatic bleeding, infection, damage to other adjacent organs, incomplete healing and death | NR | | Reisel & Creighton, 2015 |
| Long-term | | | | |
| Infertility | Childless for more than seven years<br>Primary infertility<br>Secondary infertility | 2-7 vs 2-6<br>1.4-3.3 vs 1.7<br>12.7-17.3 vs 15.5 | | Obermeyer, 2005 |

| Reproductive outcome | Effect of FGM/C | Statistics reported (where available) | | Review |
|---|---|---|---|---|
| | | Percentage | Odds ratio | |
| Gynaecological (Infection) | Bacterial vaginosis | | 1.7 | RCOG, 2015; Obermeyer, 2005; Morison et al., 2001 |
| | Herpes | | 4.7 | |
| | Urinary infections | 11 vs 6 | | De Silva, 1989; Jones, 1999 |
| | Genital infections | | 1.7 | |
| | Chronic genital abscesses, vaginal infections, Hepatitis B and HIV | NR | | Reisel & Creighton, 2015 |
| | Discharge | | 1.7-2.8 | Obermeyer, 2005 |
| | Genital ulcers | | 4.4 | |
| | Lesions | 7 vs 5 | | |
| | Damaged perineum | 62 vs 56 | | |
| | Cysts | 3 vs 2 | | |
| | Chronic pelvic infection | 13 vs 6 | | El Dareer, 1982 |
| | Abdominal pain | | 1.5 | Okonofua, 2002; Obermeyer, 2005 |
| | STIs | NR | | Elmusharaf, 2006 |
| Sexual | No sexual desire | 42 vs 16 | | Obermeyer, 2005 |
| | no orgasm | 43 vs 18 | | |
| | Reduced arousal, lubrication, orgasm, satisfaction, sexual quality of life, and dyspareunia and absence of sexual desire | NR | | Reisel and Creighton (2015) |
| Obstetric | Prolonged labour | | 1.69 | WHO, 2000; Reisel & Creighton, 2015; Berg & Underland, 2013 |
| | Obstetric/post-partum haemorrhage (PPH) | | 2.04 | |
| | | | RR: Type I (1.03), Type II (1.21), Type III (1.69) | WHO, 2006 |
| | Emergency C-section | 15.4 vs 6.5 | | Obermeyer, 2005; Reisel & Creighton, 2015 |
| | | | RR: Type I (1.03), Type II (1.29), Type III (1.31) | WHO, 2006 |
| | Difficulty in delivery | | 2.28-2.57 | Obermeyer, 2005; Berg & Underland, 2013 |
| | Foetal distress | | 2.6 | WHO, 2000; Obermeyer, 2005 |
| | Still birth | 15 vs 11 | | |
| | | | RR: Type I (1.15), Type II (1.32), Type III (1.55) | WHO, 2006 |
| | Pre-labour foetal death | | 2.5 | WHO, 2000; Obermeyer, 2005 |

| Reproductive outcome | Effect of FGM/C | Statistics reported (where available) | | Review |
|---|---|---|---|---|
| | | **Percentage** | **Odds ratio** | |
| | Early neonatal death | NR | | |
| | Obstetric lacerations | | 1.38 | Berg & Underland, 2013 |
| | Instrumental delivery | | 1.65 | |
| | Pain during and after deinfibulation (anterior episiotomy), maternal death postpartum, postnatal genital wound infection and fistulae formation | NR | | WHO, 2000 |
| | Episiotomies and perineal trauma | NR | | WHO, 2000; Reisel & Creighton, 2015 |
| | Obstetric complications | NR | | RCOG, 2015 |

*Note*. NR= data not reported

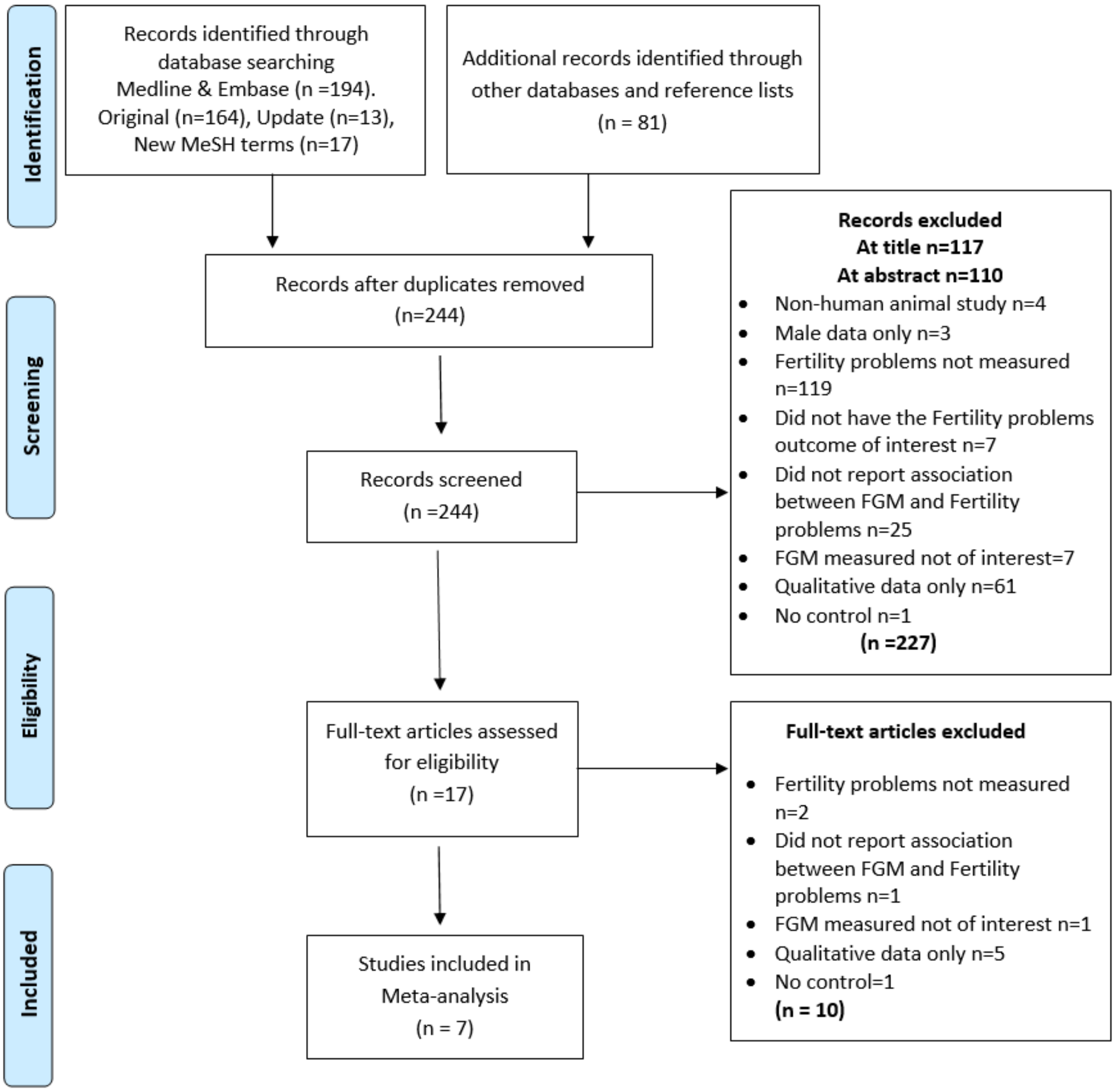


*Figure 42.* PRISMA Flow Diagram for FGM/C. Figure shows the exclusion of articles at the different stages and the reasons for exclusion. Records identified through datbase searching of Medline and Embase includes original search, an update from the time of original search and a search using new MeSH terms. FGM/C = Female Genital Mutilation/Cutting

Table 29.

Sample Characteristics Reported in the Seven Included Studies

| | Location | Sample (n) | N | N | | Age [a] Women | |
|---|---|---|---|---|---|---|---|
| | **Cross-sectional Studies** | | **FGM/C** | **No-FGM/C (control)** | | **FGM/C** | **No-FGM/C** |
| Klouman, 2005 | Tanzania | 969 women | 670 | 299 | Mean age (SD) | 27 (8) | |
| Larsen, 2000 | Central African Republic, Cote d'Ivoire, and Tanzania | 16361 women | 6124 | 10237 | NR | NR | NR |
| Larsen, 2002 | Sudan | 4218 women | 3747 | 471 | NR | NR | NR |
| Morrison, 2001 | Gambia | 776 women | 420 | 356 | NR | NR | NR |
| Yount, 2006 | Egypt | 1729 women | 1700 | 29 | Range | Percentage (n) | |
| | | | | | < 25 | 9.2 (156) | |
| | | | | | 25-34 | 39.1 (664) | |
| | | | | | 35-44 | 34.9 (593) | |
| | | | | | 45 + | 16.9 (287) | |
| | **Case-control studies** | | **Infertile [b]** | **Fertile (control)** | | **Infertile [b]** | **Fertile** |
| Almroth, 2005 | Sudan | 279 women | 99 | 180 | Mean age (SD) | 27.2 (3.9) | 24.7 (4.4) |
| | | | | | Range | Percentage (n) | Percentage (n) |
| Inhorn, 1993 | Egypt | 125 women | 39 | 86 | 0-19 | 2.2 (2) | 2 (2) |
| | | | | | 20-29 | 41.4 (37) | 47 (47) |
| | | | | | 30-39 | 49.5 (44) | 40 (40) |
| | | | | | 40+ | 7.1 (6) | 11 (11) |

*Note*: [a] Age for women at the beginning of the study; [b] Unable to become pregnant after 12 months of unprotected intercourse; FGM/C=women who have undergone Female Genital Mutilation. SD=Standard deviation NR= data not reported

Table 3.2.4.

Characteristics of the Design of the Seven Included Studies

| | Study design | Data collection | Study period | FGM/C assessment | FGM/C self-report or clinical examination | Fertility Problems outcome measure (and duration, where relevant) |
|---|---|---|---|---|---|---|

| | | | | | | |
|---|---|---|---|---|---|---|
| Klouman, 2005 | Cross-sectional | Community-based survey in rural area | 1991-1992 | Type I and II | Self-report & Clinical examination | Not able to become pregnant after 1 year living together (primary)<br>Subsequent infertile (secondary) not being able to become pregnant after 1 year from last birth<br>In the analysis combined |
| Larsen, 2000 | Cross-sectional | Demographic and Health Survey (Household interviews) | 1995, 1995, 1997 | Type I, II and III for Tanzania only. For others only cut v uncut | Self-report | Childless after more than 7 years of marriage, and subsequent infertile defined as still childless 5 years from last birth |
| Larsen, 2002 | Cross-sectional | Demographic and Health Survey (Household interviews) | 1989-1990 | Type I, II and III | Self-report | Childless after more than 7 years of marriage and subsequent infertile (5 years from last birth) |
| Morrison, 2001 | Cross-sectional | Community based survey in 17 villages (3 tribes) | Jan-July 1999 | Type I, II and III | Self-report & Clinical examination | 1 year trying to conceive |
| Yount, 2006 | Cross-sectional | Household interviews in rural area | 1995-1997 | Type I, II and IV | Self-report | Never had live birth after 5 years of marriage |
| Almroth, 2005 | Case-control | Hospital based (urban) | 2003-2004 | Anatomical extent and Type I, II and III | Clinical examination | 2 years trying to become pregnant (TFI subcategory) |
| Inhorn, 1993 | Case-control | Hospital based (urban and rural) | 1988-1989 | Type I, II and III | Self-report & medical records | 1 year trying to become pregnant, (TFI only) |

*Note***.** FGM/C = female genital mutilation/cutting; TFI = tubal factor infertility

Table 30.

Quality Ratings for the Seven Included Studies on the Basis of an Adapted Newcastle-Ottawa Quality Assessment Scale

| Study | Quality Criterion | | | | | | Overall rating [g] |
|---|---|---|---|---|---|---|---|
| | Adequacy of FGM/C (exposed) assessment[a] Max 2 points | Adequacy of control (non-exposed), definition and selection [b] Max 2 points | Comparability of control [c] Max 2 points | Confounders adequately assessed Max 2 points [d] | Adequacy of outcome Fertility Problems measure [e] Max 1 point | None response rate or loss to follow-up [f] Max 1 point | |
| Klouman, 2005 | 2 | 2 | 0 | 2 | 1 | | High |
| Larsen, 2000 | 1 | 2 | 1 | 2 | 1 | | High |
| Larsen, 2002 | 1 | 2 | 1 | 1 | 1 | | Average |
| Morrison, 2001 | 2 | 2 | 1 | 1 | 1 | | High |
| Yount, 2006 | 1 | 2 | 2 | 2 | 1 | | High |
| Almroth, 2005 | 2 | 2 | 2 | 1 | 1 | | High |
| Inhorn, 1993 | 1 | 2 | 2 | 2 | 1 | | High |

*Note*. a FGM/C was adequately assessed when independent validation of the degree of cutting was assessed (e.g. clinical examination and/or hospital/medical records) and it was representative of the cohort i.e. drawn from the same population (up to 2 points)
b Controls were adequately assessed when selection was comparable to cases, and FGM/C was excluded properly in the control population (up to 2 points)
c Comparability of controls was achieved if exposed/non-exposed were matched or adjustment during analysis conducted. One point for circumciser and one point for any other confounder (up to 2 points)
d Confounders were adequately assessed if they were obtained from records or a blind interview, and one point was given if the same method was used for both groups (up to 2 points)
e Fertility problems outcome was adequately assessed if independent or blind assessment was stated in the paper, or confirmation of the outcome by reference to secure records (medical records, etc.) (up to 1 point)
f Point given if same rate for both groups and <20% loss to follow up reported
g The overall quality rating was low (0 to 3 points), average (4 to 6 points), or high (7 to 10 points).

Table 31.

Number and Percentage of Women with Infertility Childlessness and TFI (n) in the FGM/C and No-FGM/C groups in the included studies (k=7)

| **Outcomes** | **Number of women (%)** | |
|---|---|---|
| | **FGM/C** | **Non-FGM/C** |
| Infertile (>12 months no pregnancy) | 117 of 1090 (10.7) | 61 of 655 (9.3) |
| Childlessness | 352 of 9903 (35.5) | 251 of 7760 (32.3) |
| TFI (infertile, >12 months no pregnancy) | Type II and III<br>72 of 276 (26.1) | Non-FGM/C and Type I<br>15 of 76 (19.7) |

*Note*. FGM/C = Female Genital Mutilation/Cutting; TFI = Tubal Factor Infertility

### Results of Meta-analyses

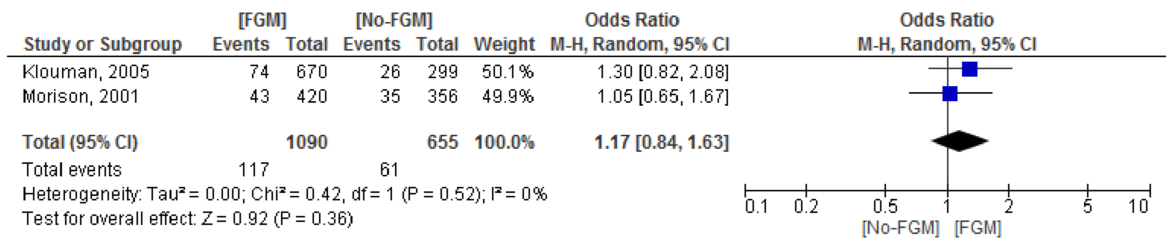


*Figure 43*. Odds ratio for proportion of 'infertile > 12 months' in the FGM/C and non-FGM/C groups

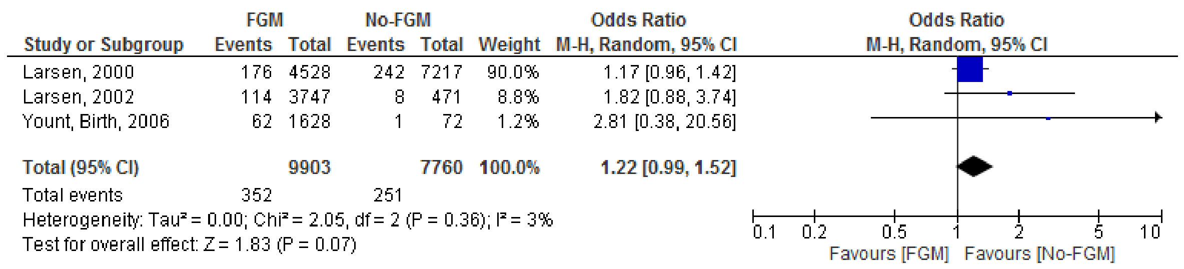


*Figure 44*. Odds ratio for proportion of 'childless' women in the FGM/C and non-FGM/C groups

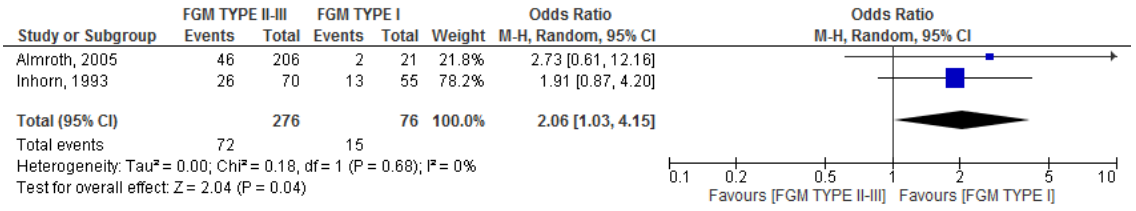


*Figure 45*. Odds ratio for proportion of women with TFI in the severe FGM/C and mild FGM/C groups

**Publication bias assessment.**

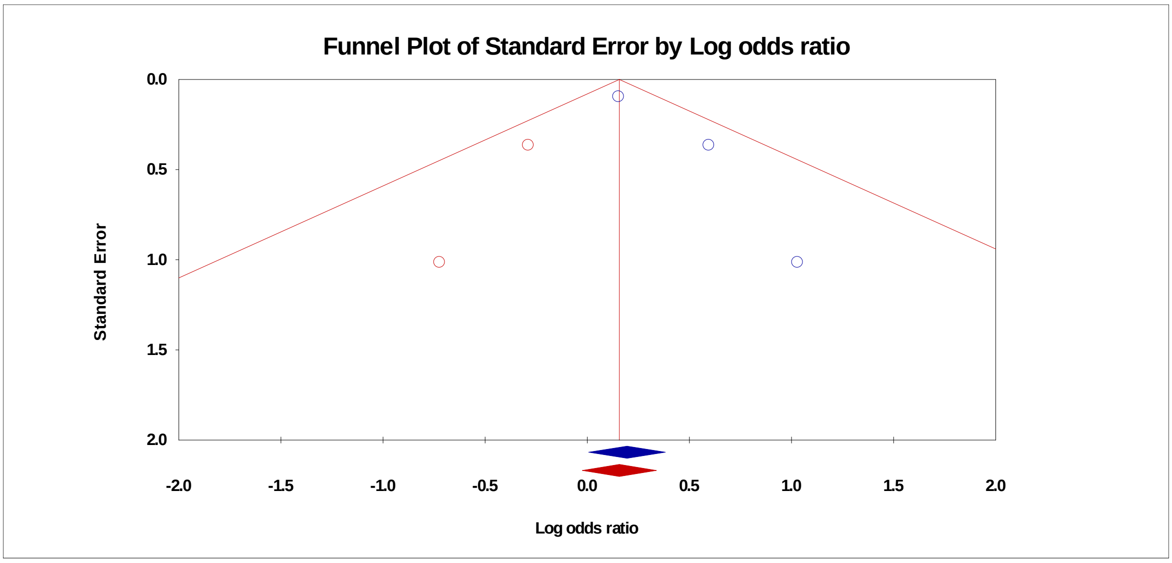


*Figure 46*. Funnel plot with trim and fill procedure to impute ‘missing’ studies (missing studies in red) for the proportion ‘childless’ analysis

### Dilatation and Curettage

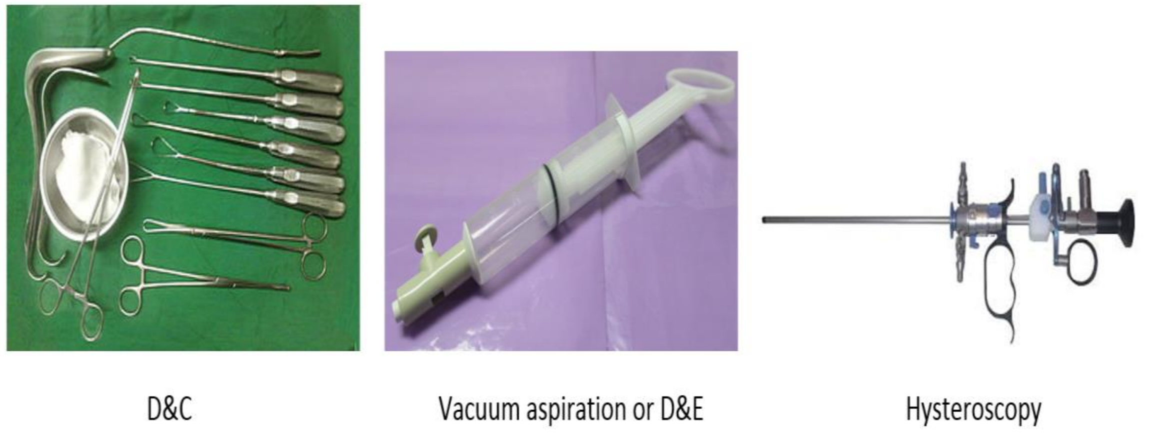


*Figure 47*. Instruments used in D&C, vacuum aspiration/D&E and hysteroscopy. D&C=dilatation and curettage, D&E=dilatation and evacuation

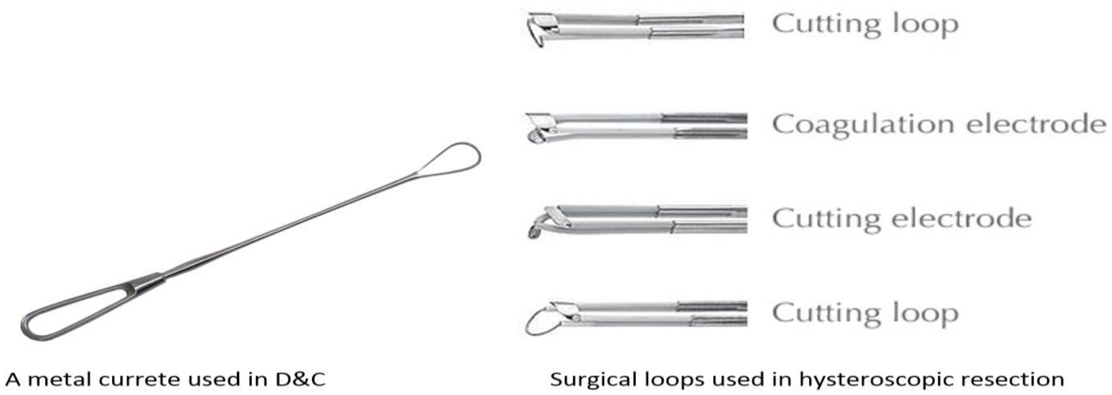


*Figure 48*. Instruments used to remove tissue in curettage as compared to hysteroscopy. D&C=dilatation and curettage, the surgical loops are found at the end of the hysteroscope

Table 32.

Summary of Long-term Negative Reproductive Outcomes Reported as a Consequence of D&C in the Literature

| Reproductive outcome | Long-term negative reproductive outcome | Primary study or review |
|---|---|---|
| **Historical literature (up to 2000)** | | |
| Single D&C | Intrauterine adhesions (IUAs), Asherman's syndrome (30.9% of women who had D&C after miscarriage) | Schenker & Margalioth, 1982; Schenker, 1996 |
| | Secondary infertility (after spontaneous miscarriage as a complication of the intrauterine surgery) | Schenker & Margalioth, 1982; Schenker, 1996 |
| | Recurrent miscarriages (after spontaneous miscarriage as a complication of the intrauterine surgery) | Schenker & Margalioth, 1982; Schenker, 1996 |
| | Negative pregnancy outcomes* after D&C (e.g. higher rates of spontaneous abortion), incompetent cervix**, preterm labour, preterm rupture of membranes, early neonatal death, and ectopic pregnancy) | Madore, Hawes, Many & Hexter, 1981; Linn et al., 1983; Kalish, Chasen, Rosenzweig, Rashbaum & Chervenak, 2002 |
| Repeated D&C | Negative pregnancy outcomes after repeated D&C (e.g. first trimester bleeding, abnormal presentations, placenta abruption, foetal distress, low birth weight, short gestation, and major malformations) | Linn, 1983 |
| | Primigravida abortion was only associated with infertility in cases where infection was present and consequently PID occurred | Hogue et al., 1983 (review) |
| | D&C as compared to vacuum aspiration was associated with negative reproductive outcomes (ectopic pregnancy, mid-trimester spontaneous abortion and low birth weight) | Hogue, 1986 (review) |
| **Current literature (2000-present)** | | |
| Single D&C | Significantly more IUAs were found after D&C compared with hysteroscopic resection*** (30% vs. 13%) | Hooker et al., 2016 (review) |
| | More postpartum haemorrhage in pregnancy following D&C (as compared to the literature) | Lohmann-Bigelow et al., 2007 |
| Repeated D&C | Odds of developing IUAs after repeated (>1) D&C were greater than after one D&C (OR 2.05, 95% CI 1.35–3.12, P=0.0008) | Hooker et al., 2014 (review) |

*Note*: D&C= dilatation and curettage, IUAs= intrauterine adhesions, PID=pelvic inflammatory disease, *Negative pregnancy outcomes are all the outcomes of a pregnancy that do not lead to a live birth (e.g. gestational problems, stillbirth) **incompetent cervix = cervical insufficiency i.e. weak cervical tissue contributes to premature birth. ***hysteroscopic resection is the removal of tissue from the uterus using a hysteroscope.

Plausible mechanisms to explain how D&/C could be associated with fertility problems

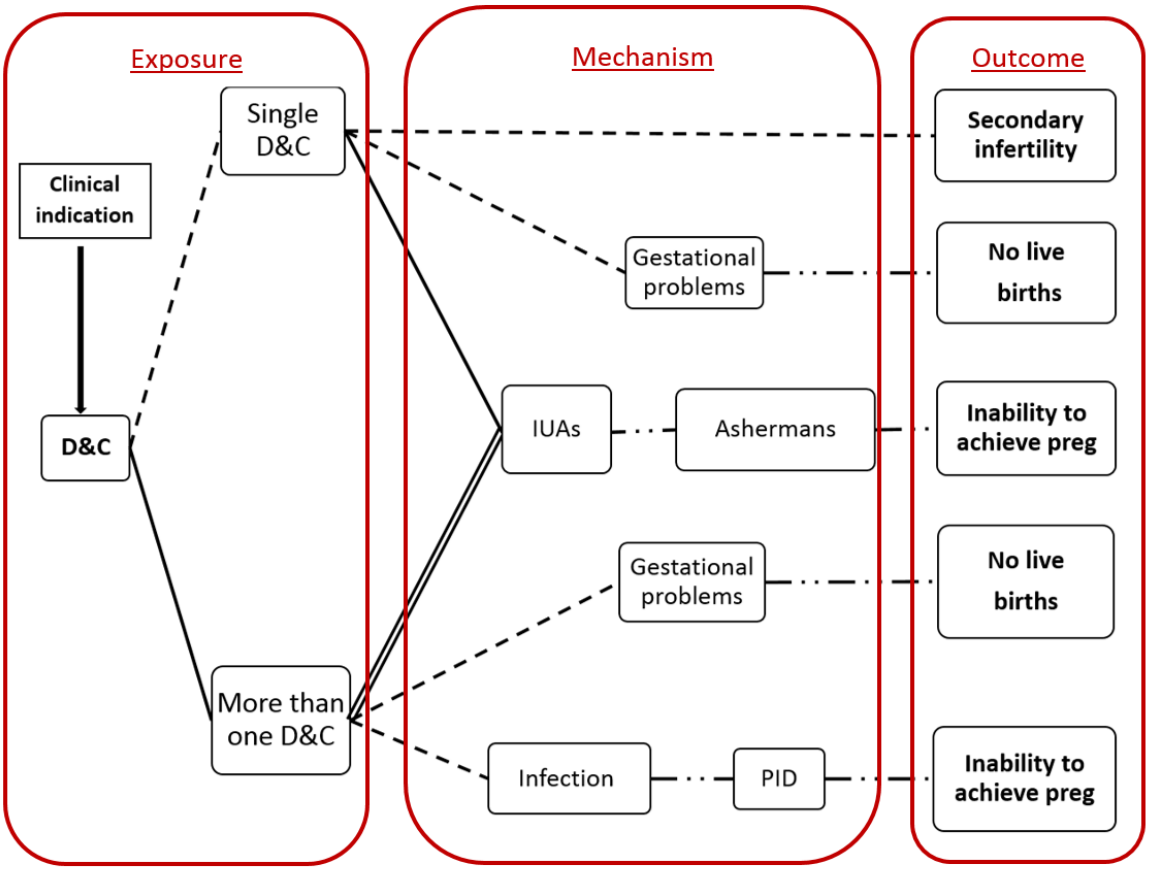


*Figure 49*. Proposed pathways for the impact of dilatation and curettage (D&C) on fertility. Solid line = Recent evidence; Double solid line= Recent Meta-analysis; Dashed line = Proposd pathway/ historic evidence; Dashed-Dotted line = Well established; Solid arrow = Moderator; D&C = dilatation and curettage; IUAs = intrauterine adhesion; gestational problems = any problem that occurs during pregancy that does not lead to a healthy live birth

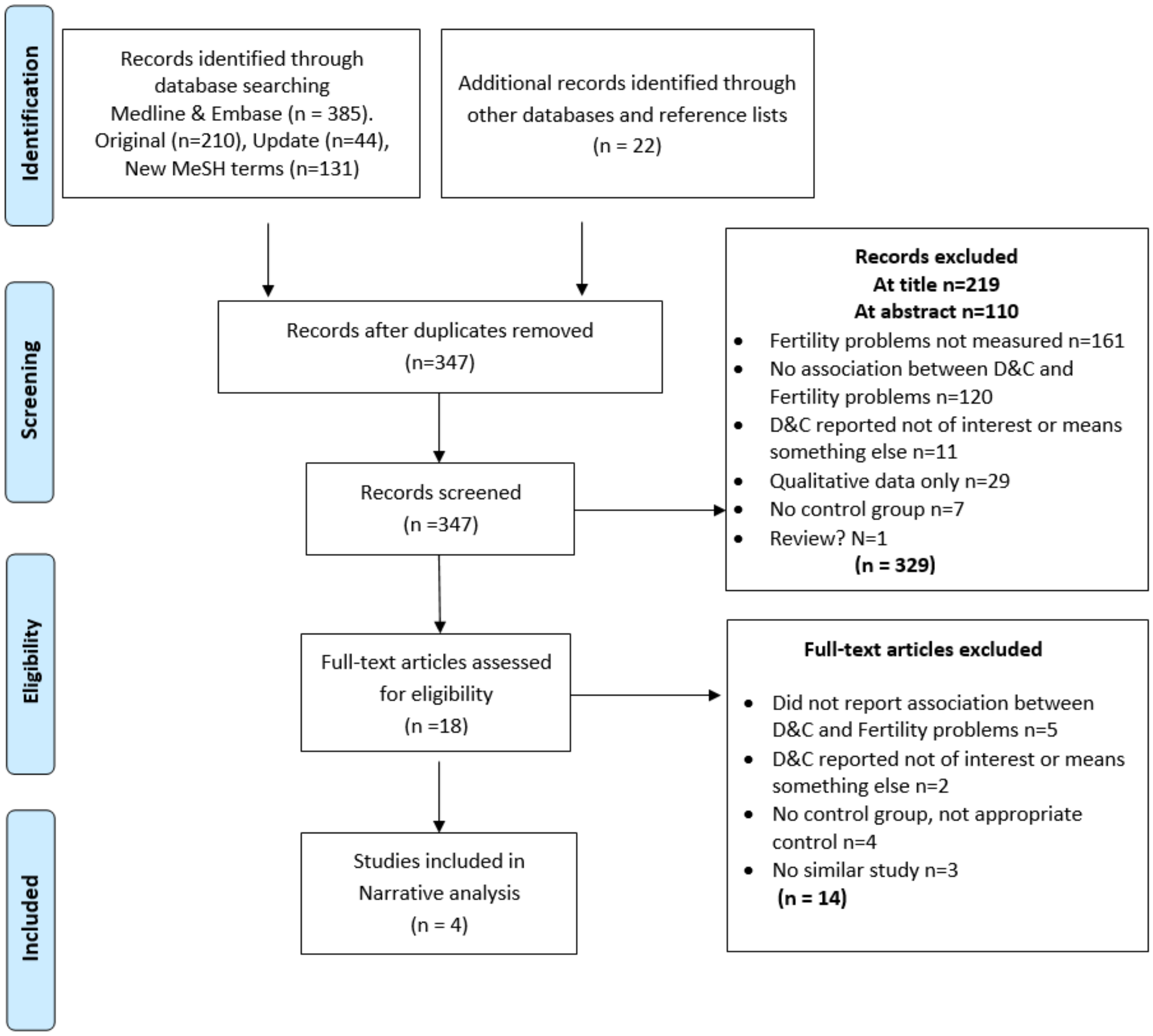


*Figure 50.* PRISMA Flow Diagram for D&C. Figure shows the exclusion of articles at the different stages and the reasons for exclusion. Records identified through datbase searching of Medline and Embase includes original search, an update from the time of original search and a search using new MeSH terms. D&C = Dilatation and Curettage

Table 33.

Sample Characteristics Reported in the Four Included Studies

| Study | Country | Sample (n) | N | N | | Age [a] Women | |
|---|---|---|---|---|---|---|---|
| | | | D&C | No-D&C[x] | | D&C | No-D&C |
| Ben-Ami, 2014 | Israel | 177 women | 94 women | 83 women | Mean (SD) | 30.4 (6.3) | 30.5 (5.9) |
| Sotnikova, 1986 | Moscow | 650 women | 350 women | 300 women | NR | NR | NR |
| Taylor, 1982 | N/A | 195 women | 53 women | 142 women | NR | NR | NR |
| Ben-Baruch, 1991 | Israel | 86 women | 52 women | 35 women | Mean (SD) | 28.6 (6.1) | 29.2 (5.0) |

*Note*. [x] type of control group described in Table 3. **[a]** Age for women at the beginning of the study; **[b]** Unable to become pregnant after at least 12 months of unprotected intercourse; D&C= dilatation and curettage; NR= data not reported; SD=Standard deviation; Shaded study from search of reference list

Table 34.

Characteristics of the Design of the Four Included Studies

| **Study** | **Study design** | **Data collection** | **Study period** | **Control Group (no-D&C)** | **Indication for procedure** | **Fertility Problems: outcomes reported in primary studies** |
|---|---|---|---|---|---|---|
| Ben-Ami, 2014 | Retrospective cohort study | Hospital based | 2000-2010 | Hysteroscopic resection | RPOC | Infertility, time to conception in months, conception rate |
| Sotnikova, 1986 | Retrospective cohort study | NR | NR | PG & vacuum suction | Induced abortion | Gynaecological diseases (e.g. salphingophoitis, endometriosis), menstrual dysfunction (e.g. biphasic menstrual cycle, insufficient luteal phase) |
| Taylor, 1982 | Cross-sectional study | Hospital based | NR | Did not undergo D&C | Routine investigation for infertility | PID, endometriosis and fibroid |
| Ben-Baruch, 1991 | Prospective cohort study | Hospital based | 19983-1988 | Expectant management | Spontaneous abortion (miscarriage) | Infertility (attempted conception > 12) months after abortion or stopping contraception. Future pregnancy, miscarriage and normal delivery. |

*Note*: D&C= dilatation and curettage; NR= data not reported; RPOC = retained products of conception; PG = prostaglandins; PID = pelvic inflammatory disease. Shaded study from search of reference list

Table 35.

Quality Ratings for the Four Included Studies on the Basis of an Adapted Newcastle-Ottawa Quality Assessment Scale

| Study | Quality Criterion | | | | | | Overall rating [g] |
|---|---|---|---|---|---|---|---|
| | **Adequacy of D&C (exposed) measure [a] Max 2 points** | **Adequacy of control (non-exposed), definition and selection [b] Max 2 points** | **Comparability of control [c] Max 2 points** | **Confounders adequately assessed Max 2 points [d]** | **Adequacy of outcome Fertility Problems measure [e] Max 1 point** | **None response rate or loss to follow-up [f] Max 1 point** | |
| Ben-Ami, 2014 | 2 | 2 | 2 | 2 | 0 | 0 | High |
| Sotnikova, 1986 | 0 | 0 | 0 | 0 | 1 | 0 | Low |
| Taylor, 1982 | 1 | 2 | 1 | 1 | 2 | NA | High |
| Ben-Baruch, 1991 | 2 | 2 | 1 | 0 | 0 | 0 | Average |

*Note*. [a] D&C was adequately assessed when hospital/medical records were available and sample was drawn from the same population (up to 2 points); [b] Controls were adequately assessed when selection was comparable to cases, and D&C was excluded properly in the control population (up to 2 points); [c] Comparability of controls was achieved if exposed/non-exposed were matched or adjustment during analysis conducted. One point for *'obstetric history'* and one point for any other confounder (up to 2 points); [d] Confounders were adequately assessed if they were obtained from records or a blind interview, and one point was given if the same method was used for both groups (up to 2 points); [e] Fertility problems outcome was adequately assessed if independent or blind assessment was stated in the paper, or confirmation of the outcome by reference to secure records (medical records, etc.) (up to 1 point); [f] Point given if same rate for both groups and <20% loss to follow up reported, NA: not applicable; [g] The overall quality rating was low (0 to 3 points), average (4 to 6 points), or high (7 to 10 points). Shaded from search of ref list

Table 36.

Summary of Methodological Considerations and Results of the Four Included Studies

| Study | Indication for procedure | Control Group (no-D&C) | Other factors | Follow up period | Results: Outcomes reported in primary studies | |
|---|---|---|---|---|---|---|
| | | | | | Significant difference | No significant difference |
| Ben-Ami, 2014 | RPOC after birth, spontaneous or induced abortion | Hysteroscopic resection (HR) | More HR after birth and more D&C after abortion<br><br>D&C group more abdominal pain (before procedure), HR group longer time from birth/abortion to RPOC | NR | More infertility in the D&C group<br><br>Longer time to pregnancy (months) in the D&C group | Desire for pregnancy<br><br>Achieve pregnancy |
| Ben-Baruch, 1991 | Spontaneous abortion (miscarriage) | Conservative management (waiting) | Which treatment would be performed was decided by treating physician | 28 months (range 12-68) in the D&C group 26 months (range 12-72) in the control group | | Achieve pregnancy, miscarriage and normal delivery.<br><br>Infertility (including existing and new cases) |
| Sotnikova, 1986 | Induced abortion | Group 1- PG OR vacuum suction | Gynaecological history (e.g. age at menarche, genital inflammation) was reported | One year | More gynaecological diseases (e.g. inflammation of fallopian tubes, endometriosis) in the D&C group | |
| | | Group 2 - PG | | 5 years | More menstrual dysfunction (e.g. anovulation, oligomenorrhea, insufficient luteal phase) in the D&C group | |
| Taylor, 1982 | Routine investigation for infertility | Did not undergo D&C | Excluded women with history of PID, pelvic surgery abnormal menstruation | History of D&C or no-D&C | More PID in the D&C group | Endometriosis and fibroid |

*Note*: D&C = dilatation and curettage; RPOC = retained products of conception; HR = Hysteroscopic resection; PG = prostaglandins; PID = pelvic inflammatory disease; NR = not reported.

### Vitamin D deficiency

Plausible mechanisms to explain how vitamin D deficiency could be associated with fertility problems

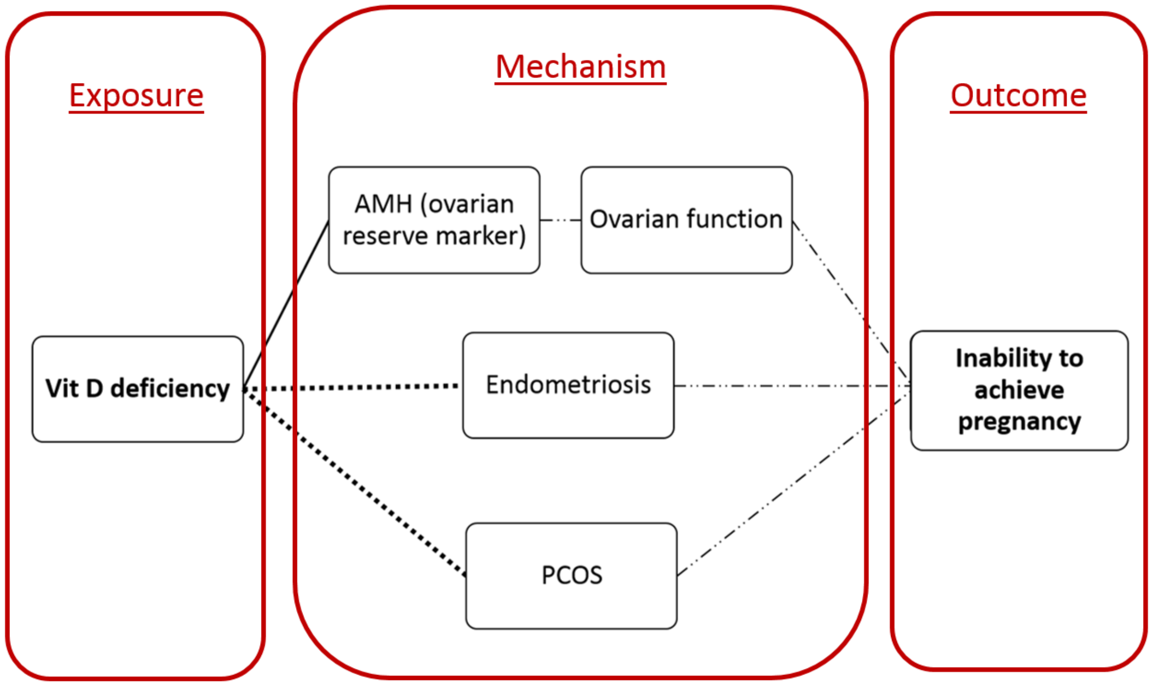


*Figure 51*. Proposed pathways for the impact of Vitamin D deficiency on fertility. Solid line = Recent evidence (primary molecular studies); Double solid line = meta-analytic evidence; Dotted line = inconsistency in results of primary studies and/or meta-analyses; Dashed line =Proposed pathway/historic evidence; Dashed-Dotted line = Well established

### Applicable to all RFs

Table 37.
Summary of evidence reviewed, outcomes reported, number of studies in each meta-analysis and pooled effects estimate.

| RF | Evidence reviewed | Outcome reported | Number of studies included in MA | Pooled effect estimates OR (95% CI)/ Mean Difference (95% CI) |
|---|---|---|---|---|
| **CSG** | 451 records retrieved, 24 studies included in MA | Time to first birth | 2 | MD 0.24 (-0.39-0.87) p=0.46 |
| | | Miscarriage | 5 | 1.1 (0.93-1.30) p=0.25 |
| | | Never-pregnant | 3 | 0.66 (0.45-0.98) p=0.04 |
| | | Childlessness | 5 | 0.83 (0.67-1.03) p=0.09 |
| | | Mean # pregnancies | 5 | MD 0.40 (0.10-0.71) p=0.009 |
| | | Mean # live-births | 7 | MD 0.24 (0.05-0.43) p=0.01 |
| | | Stillbirth | 5 | 1.28 (1.04-1.57) p=0.02 |
| | | Neonatal Death | 4 | 1.57 (1.22-2.02) p=0.0005 |
| **FGM/C** | 244 records retrieved, 7 studies included in MA | Infertile > 12 months no pregnancy | 2 | 1.17 (0.84-1.63) p=0.36 |
| | | Childlessness | 3 | 1.22 (0.99-1.52) p=0.07 |
| | | Infertile 2 yrs (TFI)* | 2 | 2.06 (1.03-4.15) p=0.04 |
| **HIV** | 741 records retrieved, 9 included in MA | Cumulative Pregnancy rate | 2 | 0.36 (0.15-0.89) p=0.03 |
| | | Miscarriage | 2 | 0.03 (-0.03-0.09) p=0.35 |
| | | Amenorrhea | 3 | 2.44 (1.56-3.81) p<0.00001 |
| | | FSH >25 IU/l | 2 | 1.51 (0.77-2.94) p=0.23 |
| | | Infertile > 12 months no pregnancy* | 2 | 2.93 (1.95-4.42) p<0.00001 |
| **GTB** | 451 records retrieved, 5 included in MA | Infertile >12 months no pregnant | 2 | 8.91 (1.89-42.12) p=0.006 |
| | | Amenorrhea | 2 | 4.24 (0.23-78.14) p=0.33 |
| | | Primary infertility | 2 | 2.94 (1.89-4.37) p<0.00001 |
| **BV** | 184 records retrieved, 11 included in MA | Infertile > 12 months no pregnancy* | 11 | 2.81 (1.85-4.27) p<0.00001 |
| | | **Narrative reviews** | | |

| RF | Evidence reviewed | Outcome reported | Number of studies included in MA | Pooled effect estimates |
|---|---|---|---|---|
| | | | | OR (95% CI)/ Mean Difference (95% CI) |
| **D&C** | 347 records retrieved, 4 included in narrative review | Infertile > 12 months no pregnancy | 1 | Significantly more than hysteroscopy group |
| | | Time to pregnancy | 1 | Significantly longer than hysteroscopy group |
| | | Gynaecological diseases (e.g. inflammation of fallopian tubes, endometriosis) | 1 | More in the D&C than vacuum aspiration of prostaglandins. |
| | | PID | | More in the D&C than no treatment group |
| **Vitamin D Deficiency** | No review necessary | NA | 0 | NA |
| **Water-pipe** | No review necessary | NA | 0 | NA |

*Note*. * = data converted from case-control studies. RF = risk factor; OR = odds ratio; NA = not applicable; MA = meta-analysis; CSG = consanguinity; FGM/C = female genital mutilation/cutting; GTB = genital tuberculosis; BV = bacterial vaginosis; D&C = dilatation and curettage; PID = pelvic inflammatory disease.

Table 38.

Summary of which Bradford-Hill Criteria were met for each of the six Risk Factors included in Systematic Review

| **Criteria** | **Risk Factor** | | | | | |
|---|---|---|---|---|---|---|
| | CSG | FGM/C | HIV | GTB | BV | D&C |
| Strength | | | | **X** | **X** | |
| Consistency | | | | **X** | **X** | **X** |
| Specificity | | **X** | | | | **X** |
| Temporality | **X** | **X** | | | | |
| Biological gradient | **X** | **X** | | | **X** | **X** |
| Plausibility | **X** | **X** | | **X** | **X** | |
| Coherence | | | | **X** | **X** | |
| Experiment | | | | | | |
| Analogy | | | | | | |

*Note*. Bradford-Hill Criteria from Hill, 1965. CSG = consanguinity; FGM/C = female genital mutilation/cutting; GTB = genital tuberculosis; BV = bacterial vaginosis; D&C = dilatation and curettage
